# Pharmacological Management of Gambling Disorder: A Systematic Review and Network Meta-Analysis

**DOI:** 10.1101/2023.10.20.23297314

**Authors:** Konstantinos Ioannidis, Cinzia Del Giovane, Charidimos Tzagarakis, Jeremy E Solly, Samuel J. Westwood, Valeria Parlatini, Henrietta Bowden-Jones, Jon E Grant, Samuele Cortese, Samuel R Chamberlain

**Author notes:** **Corresponding author**; Dr Konstantinos Ioannidis PhD, Consultant Psychiatrist, Clinical Lead, Southern Gambling Service, College Keep, 4-12 Terminus Terrace, Southampton. SO14 3DT. equally contributed (Samuel J. Westwood, Valeria Parlatini).

## Abstract

**Background:** Clinical guidelines remain unclear on which medications for gambling disorder are to be preferred in terms of efficacy and tolerability. We aimed to compare pharmacological treatments for gambling disorder in terms of efficacy and tolerability, using network meta-analysis (NMA).

**Methods:** We searched, up to 19 February 2024, a broad range of databases, including MEDLINE, EMBASE, PsycINFO, PubMed, CINAHL, AMED, and the Cochrane Database of Systematic Reviews, ERIC and Web of Science (including Science Citation Index Expanded (SCI-EXPANDED), Social Science Citation Index (SSCI), Conference Proceedings Citation Index-Science (CPCI-S) and Conference Proceedings Citation Index-Social Science and Humanities (CPCI-SSH)) via Web of Knowledge and the WHO International Trials Registry Platform (including ClinicalTrials.gov), for double-blind randomised controlled trials (RCTs) of medications for gambling disorder. Outcomes were gambling symptom severity and quality of life (for efficacy), and tolerability. Confidence in the network estimates was assessed using the CINeMA framework. We followed the PRISMA-NMA guidelines and pre-registered the protocol [CRD42022329520].

**Outcomes:** We included 22 RCTs in the systematic review and 16 RCTs (n = 977 participants) in the NMA. Compared with placebo, moderate confidence evidence indicated that nalmefene [Standardized Mean Difference (SMD): −0·86; 95% confidence interval (CI: −1·32,-0·41)] reduced gambling severity, followed by naltrexone [-0·42; −0·85, 0·01)]. Nalmefene [Odds Ratio (OR): 7·55; 95%CI: 2·24-25·41] and naltrexone (7·82; 1·26-48·70) had significantly higher dropout due to side effects (lower tolerability) compared with placebo. Naltrexone (SMD: −0·50; 95%CI: −0·85,-0·14) and nalmefene (−0·36; −0·72,-0·01) were more beneficial than placebo in terms of quality of life. Olanzapine and topiramate were not more efficacious than placebo.

**Interpretation:** Nalmefene and naltrexone currently have the most supportive evidence for the pharmacological treatment of gambling disorder. Further clinical trials of novel compounds, and analysis of individual participant data are needed, to strengthen the evidence base, and help tailor treatments at the individual patient level.

**Funding:** This study was supported by unrestricted grant funds to Professor Chamberlain held at the University of Southampton, originating from the NHS.

**Research in Context:** *Evidence before this study:* Over the past few decades, there has been a substantial need for evidence-based pharmacological treatments of gambling disorder. However, the benefits and safety of medications trialled to treat gambling disorder remains debateable. Before planning this study, we searched PubMed on 01.06.2022 (and again on 16.03.2024 for any new evidence) for meta-analyses of randomised controlled trials (RCTs) of pharmacological treatments for gambling disorder, using the following syntax/search terms: gambling [tiab] AND meta-analy* [tiab]. We found two recent pairwise meta-analyses assessing the efficacy and tolerability of individual medications. However, we could not find any network meta-analysis (NMA) providing evidence on the comparative efficacy and tolerability of medications used for the treatment of gambling disorder. As NMAs have been successfully used to inform treatment approaches for other conditions, the lack of NMA of pharmacological treatments for gambling disorder is an important gap.

*Added value of this study:* We conducted the first NMA of pharmacological treatments for gambling, based on state-of-the-art methodology for NMA. Our NMA represents the most comprehensive synthesis to date on the comparative efficacy and tolerability of pharmacological options to treat gambling disorder. Unlike previous systematic reviews and pairwise meta-analyses of head-to-head trials, we considered each treatment option separately (i.e., not as “class of medication”) and focused on clinically relevant outcomes – namely, the efficacy on gambling severity symptoms, tolerability and effects on the quality of life. We found that nalmefene and naltrexone currently have the most supportive evidence, in terms of clinical efficacy (reduction of gambling severity and improvement in quality of life), for the pharmacological treatment of gambling disorder, whereas olanzapine or topiramate had less or no supportive evidence. We also found that nalmefene and naltrexone were less well tolerated than placebo, which highlights the need for future clinical trials to broaden the evidence base.

*Implications of all the available evidence:* Evidence from our NMA supports the use of nalmefene and naltrexone in adults with gambling disorder as the preferred first pharmacological choice for the management of gambling disorder. Our NMA should inform future guidelines and supplement clinical decision-making on the choice of treatment for adults with gambling disorder, along with available evidence on psychological options, evidence on cost-effectiveness, and patients’ preferences. Future studies should evaluate a broader range of pharmacological agents for the treatment of gambling disorder .Future research should also include individual patient data in NMA of gambling disorder medications, which will allow a wider and more reliable estimation of predictors of individual response.

## INTRODUCTION

Gambling disorder is a complex behavioural addiction, which affects individuals and those around them and has substantial public health implications worldwide.^1^ It is characterized by persistent and recurrent gambling leading to negative consequences, including e.g. interpersonal conflict, serious financial problems, homelessness, mortgage foreclosure, and elevated risk of suicide.^2,3^ Gambling disorder is currently classified as a behavioural addiction in the International classification of Disease 11^th^ Edition (ICD-11)^4^ and as a Substance-Related and Addictive Disorder in the Diagnostic and Statistical Manual 5^th^ Edition (DSM-5-TR).^5^

Psychological interventions (i.e., gambling focused cognitive-behavioural therapy –CBT–, in its many variants) are widely used to treat gambling disorder.^3^ However, there is no consensus about the most effective treatment strategies. Some international guidelines suggest, further to CBT, pharmacological options (specifically, naltrexone) for treatment-resistant gambling disorder^6^ or adjunct pharmacology to talking therapies or as monotherapy.^7^ Also, those guidelines recommend against the use of antidepressants as monotherapy for gambling disorder, unless there is comorbid depression or anxiety.

A growing body of studies of pharmacological treatments for gambling is emerging. There have been previous attempts to pool evidence from these studies. A pairwise meta-analysis including 34 studies (open label, non-randomised and randomised control trials [RCTs], with or without concomitant psychological interventions), showed large effects for pharmacological treatments overall (Hedge’s g=1·35 in terms of global severity of gambling; medium effect size, g=0·41 when including RCTs only).^8^ Moreover, that review was not designed to compare different compounds, and moreover, did not find differences in effects between classes of medication.^8^

A recent Cochrane systematic review and pairwise meta-analysis of the pharmacological interventions of disordered and problem gambling^9^ used a major-category examination approach (e.g. “antidepressants”, “opioid-antagonists”) across 17 RCTs (n=1193). The meta-analysis found evidence that antidepressants and mood stabilizers were no significantly better than placebo in treating gambling symptoms. Opioid antagonists (SMD −0.46, 95% CI −0.74 to −0.19) and atypical antipsychotics (SMD −0.59, 95% CI −1.10 to −0.08) were found to be beneficial in treating gambling symptoms. However, this meta-analysis only considered direct (head-to-head) comparisons, and this limited the number of comparisons included due to the paucity of head-to-head studies for gambling disorder.^9^ Moreover, the Cochrane review and previous meta-analyses did not consider any quality of life outcomes, which are highly relevant outcome in addition to gambling symptoms severity.

Thus, many important questions on the pharmacological treatment of gambling remain unanswered, including: 1) which individual compounds (even from those within the same class, e.g. opioid receptor antagonists) are the most efficacious for gambling disorder? 2) which are best tolerated medications when compared to each other? and 3) which medications have the strongest impact on quality of life?

Network meta-analysis (NMA) can address these crucial gaps in the field, by providing, under certain assumptions, comparative evidence on the efficacy and tolerability of two or more treatments, even when they have not been directly compared in the individual trials included in the NMA.^10^ This evidence can then be used to rank or compare the effects of several interventions simultaneously.^11^ One of the advantages of NMAs is that by adding indirect evidence, they increase the precision of an effect size estimate, even when there is direct evidence for that specific comparison.^10^ Notably, while the useful insights produced by NMAs are well established, many NMAs are commissioned by industry, are not pre-registered, and never get published to allow for an equitable dissemination of knowledge and public health benefits.^12^ At a time when the UK National Institute for Health and Care Excellence (NICE) is developing initial clinical guidelines for gambling disorder,^13^ an NMA is urgently needed to allow for a thorough and up-to-date examination of evidence supporting pharmacological treatment(s) for gambling disorder. Such work is also likely to directly inform other international guidelines (whether new or updated) in the future. Therefore, we conducted the first NMA to compare the effects of pharmacological treatments on symptoms and quality of life, in the management of gambling disorder, as well as the relative tolerability of such treatments.

## METHODS

The study protocol was pre-registered on the PROSPERO International prospective register of systematic reviews [Registration number: CRD42022329520 Available from: https://www.crd.york.ac.uk/prospero/display_record.php?RecordID=329520]. This study reporting followed the PRISMA-NMA guidelines^14^ Additional methodological details are presented in the appendix (pp 3-4). The PRISMA-NMA checklist is shown in the appendix (pp 5-7).

### Search strategy

We searched for published and unpublished data. The search strategy and syntax were determined by consensus amongst the co-authors, with further expert refinement from Systematic Review Solutions Ltd. (SRS), an independent professional company specialising in meta-research. The search strings used and full list of electronic databases and clinical trial registries in which the search was conducted are available in the appendix (pp 8-16). The initial search was conducted on the 13^th^ of July 2022 and then updated on the 19^th^ of February 2024.

### Eligibility criteria

We included RCTs comparing an active medication vs. placebo, or active medications with each other, for the treatment of Gambling Disorder/Pathological Gambling. Trials with a cross-over design were included if data from the pre cross-over phase were available, to avoid carry-over effects.^15^ We included only studies of adults (>18yrs) with a primary DSM (III onwards) or ICD (9 onwards) diagnosis of Gambling Disorder/Pathological Gambling. For the NMA, we excluded studies which had insufficient data, or those that specifically included a primary psychiatric condition in the whole sample, other than gambling as part of inclusion criteria.

### Data extraction and outcomes

Details on data extraction can be found in the appendix (p 3).

### Choice of primary measure

The primary efficacy outcome was gambling symptom severity measured by well-established and validated instruments, namely the Yale-Brown Obsessive Compulsive Scale adapted for Pathological Gambling (PG-YBOCS),^16^ the Gambling Symptom Assessment Scale (G-SAS),^17^ and the Clinical Global Impression-Improvement scale (CGI-I).^18^ If a study reported results from multiple scales, we used the following hierarchy in the choice of the scale: PG-YBOCS as first preference; G-SAS as next preference, CGI-I as third preference. This was done to prioritize structured clinical instruments against unstructured or self-report instruments. The primary tolerability outcome was defined as the proportion of patients who exited a given study due to side-effects. The secondary efficacy outcome was the improvement in quality of life and functioning as measured by validated instruments including but not limited to the Sheehan Disability Scale (SDS)^19^ and other Quality of life metrics. Where outcome data or any relevant information were missing, the corresponding authors of the individual papers were contacted with a request to provide unpublished data/information.

### Data synthesis

We calculated the standardized mean differences (SMD) using Hedges’ g to measure the efficacy outcomes, because different scales were used to assess the same outcome. The measure of effect for tolerability was the dropout rate due to medication side effects, expressed as odds ratio (OR). Study arms randomizing the same compound at different dose were merged into a single arm. First, we conducted conventional pairwise meta-analyses with a random-effects model for all outcomes and treatment comparisons with at least two studies,^20^ followed by frequentist NMA for all outcomes using random-effects models. We evaluated the assumption of transitivity within NMA by assessing the similarities of the distributions of the potential effect modifiers, such as study and patient-level covariates, across pairwise comparisons.^21^ All results were reported as treatment effects with their 95% confidence intervals (95%CIs). In standard pairwise meta-analysis, we assessed the heterogeneity within each comparison visually by considering the forest plot, and quantitatively with the I² statistic and the τ^2^.^20^ In NMA, we assumed a common parameter for heterogeneity across comparisons in each network and we estimated the heterogeneity standard deviation τ for each outcome.^22^ Statistical incoherence was evaluated globally by using the design-by-treatment approach and locally by using both the node-splitting approach and the loop-specific approach.^23^ Surface Under the Cumulative RAnking (SUCRA)^24^ curves were used to measure, for any outcome, the probability for each treatment to provide the best/worst outcome among all treatments included in the network. Additionally, we calculated other available ranking metrics, i.e. mean rank and the probability to provide best outcome.^22,24^ We also performed a sensitivity analysis by conducting NMA for gambling severity, by analysing each scale of gambling severity separately, and restricting the analysis to mean difference (MD) only. We used STATA®/IC 18.1 (StataCorp LLC, College Station, TX, USA) to perform all analyses.

### Study risk of bias assessment and confidence in Network Meta-Analysis estimates (CINeMA)

We used the Confidence in Network Meta-Analysis (CINeMA) framework to assess the confidence in the estimates obtained from the NMA; this was done separately for gambling severity, tolerability and quality of life. The framework considers six domains: within-study bias, reporting bias, indirectness, imprecision, heterogeneity, and incoherence.^25^ We evaluated the reporting bias using RoB-MEN framework as part of CINeMA, which includes a variety of statistical (e.g. funnel plots, meta-regressions) and non-statistical methods (using the Outcome Reporting Bias in Trials [ORBIT] classifications).^26^ For within-study bias, each individual RCT included in our Network Meta-Analysis was assessed using the Cochrane Risk of Bias tool version 2 (RoB-2).^27^ More information about how CINeMA was implemented is presented in the appendix (pp 17-18).

## RESULTS

The search yielded 4261 references from electronic databases and 71 hits from clinical trial registries. A final set of 22 eligible RCTs were selected for inclusion in the systematic review. Six of these RCTs were excluded from the NMA either due to insufficient data (n=4), or the inclusion of a primary psychiatric co-concurrent condition in the whole sample (n=2), which we deemed violated the NMA transitivity assumption. Randomized participants were ∼49% males (674/1371), and their ages ranged from 29·7 to 51·5 years (Mean=43·56; SD=5·81). Each of the 16 RCTs included in the NMA (total participants: 977) contributed to one pairwise comparison, totalling 16 pairwise comparisons across studies (16 for gambling severity, 12 for tolerability, nine for quality of life). Full details about the search results are presented in the PRISMA flowchart (Figure 1). Full characteristics of the studies, RoB 2 assessment and ORBIT classifications are presented in the appendix (pp 19-23).

**Figure 1.**
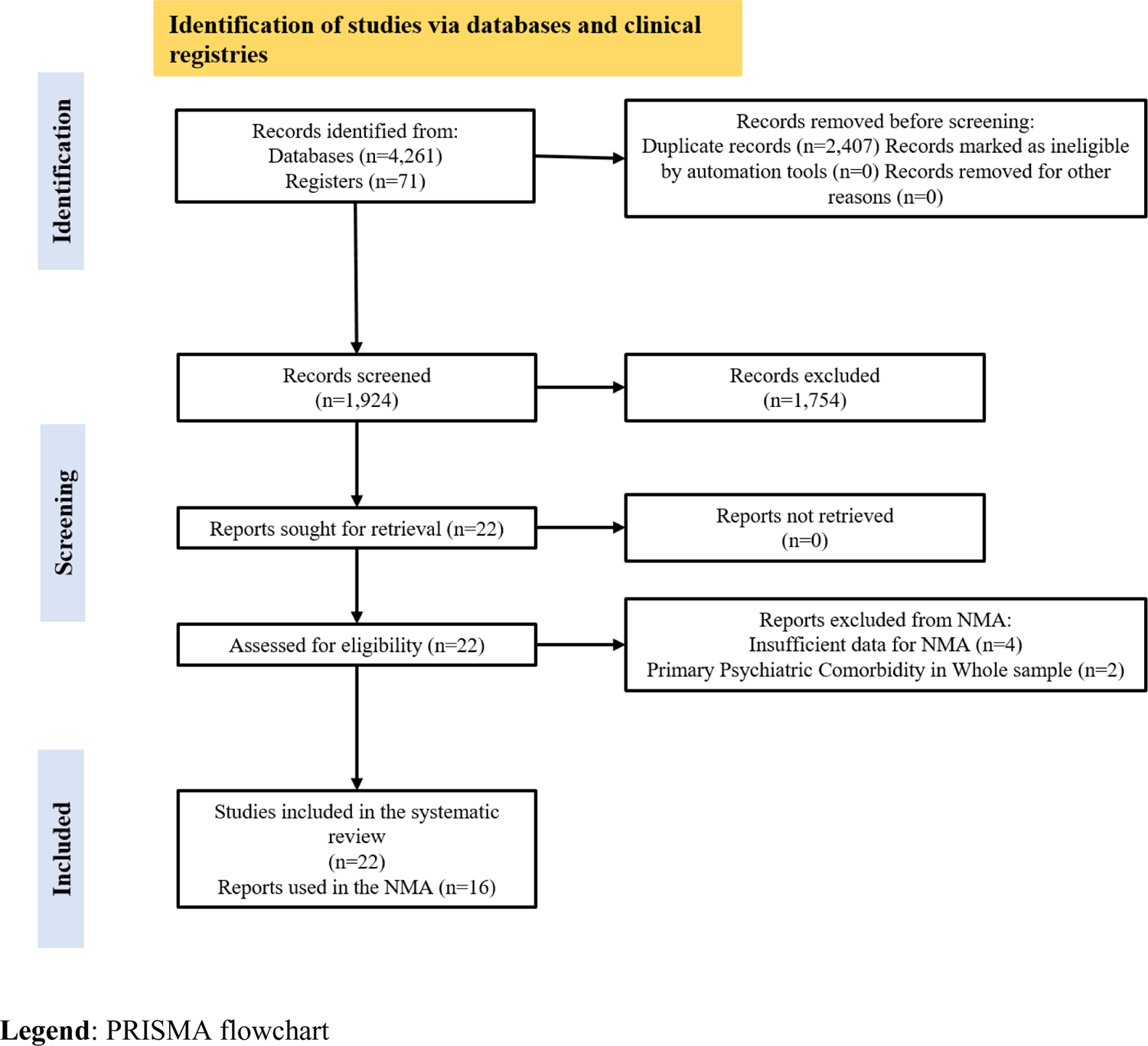
PRISMA Flowchart

Comparisons included in the NMA comprised nine different medications: three opioid receptor antagonists, (naltrexone, nalmefene, naloxone); two selective serotonin reuptake inhibitors (SSRIs - paroxetine and fluvoxamine); one mood stabilizer/antiepileptic (topiramate); one norepinephrine–dopamine reuptake inhibitor (NDRI, bupropion); one antipsychotic (olanzapine); and one plant-based antioxidant (silymarin). Six more medications were included in the studies retained in the systematic review only: lithium, sertraline, clomipramine, baclofen, acamprosate and n-acetyl-cysteine (NAC). We did not identify any clear evidence that the transitivity assumption did not hold.

Results from the conventional pairwise meta-analysis for each outcome and within each treatment comparison are showed in the forest plots in the appendix (pp 25-30). Figure 2 shows the network plots for efficacy on gambling severity, tolerability and efficacy on quality of life.

**Figure 2.**
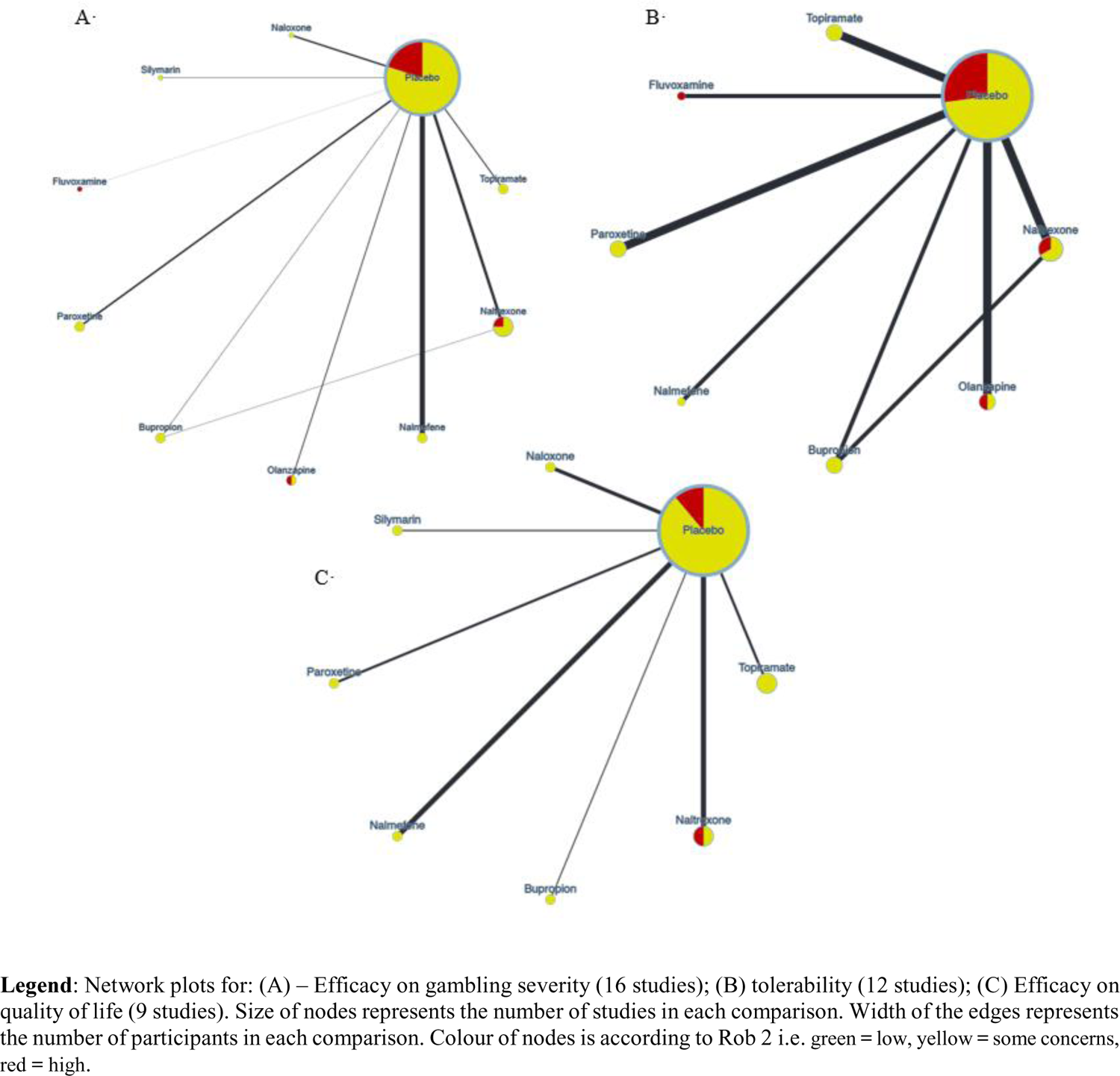
Network plots

Forest plots of NMA results for gambling severity, tolerability, and quality of life showing the network estimates for each treatment versus placebo are presented in Figure 3. The results of the NMA for efficacy on gambling severity and tolerability for all possible comparisons are shown in Table 1. The results of the NMA for quality of life are presented in Table 2. The NMA showed that, in terms of gambling severity, among nine active drugs and placebo, nalmefene [SMD: −0·86; 95% confidence interval (CI: −1·32, −0·41)] was associated with higher efficacy compared with placebo, followed by naltrexone [SMD: −0·42; 95%CI: −0·85, 0·01)]. Across medications, we identified a superiority of nalmefene over naloxone (SMD 1·01 (95%CI: 0·20, 1·82)). In terms of tolerability, nalmefene (OR 0·13 (95%CI: 0·04, 0·45)) and naltrexone (OR 0·13 (95%CI: 0·02, 0·80)) were found to be the least tolerated (i.e., having the highest dropout risk due to side effects) as compared with placebo. We did not find any significant difference between treatments. Naltrexone (SMD −0·50 (95%CI: −0·85, −0·14)) and nalmefene (SMD −0·36 (−0·01, −0·72) were associated with higher quality of life outcomes compared with placebo. Across medication treatments, we identified a superiority of nalmefene over naloxone (SMD −1·01 (95%CI: −1·82, −0·20) on gambling severity and naltrexone over naloxone (SMD −0·61 (95%CI: −1·13, −0·09)) on quality of life. Sensitivity analyses did not indicate any material changes the main NMA results. Main NMA results from sensitivity analyses of gambling severity are presented in the appendix (pp 31-37).

**Figure 3.**
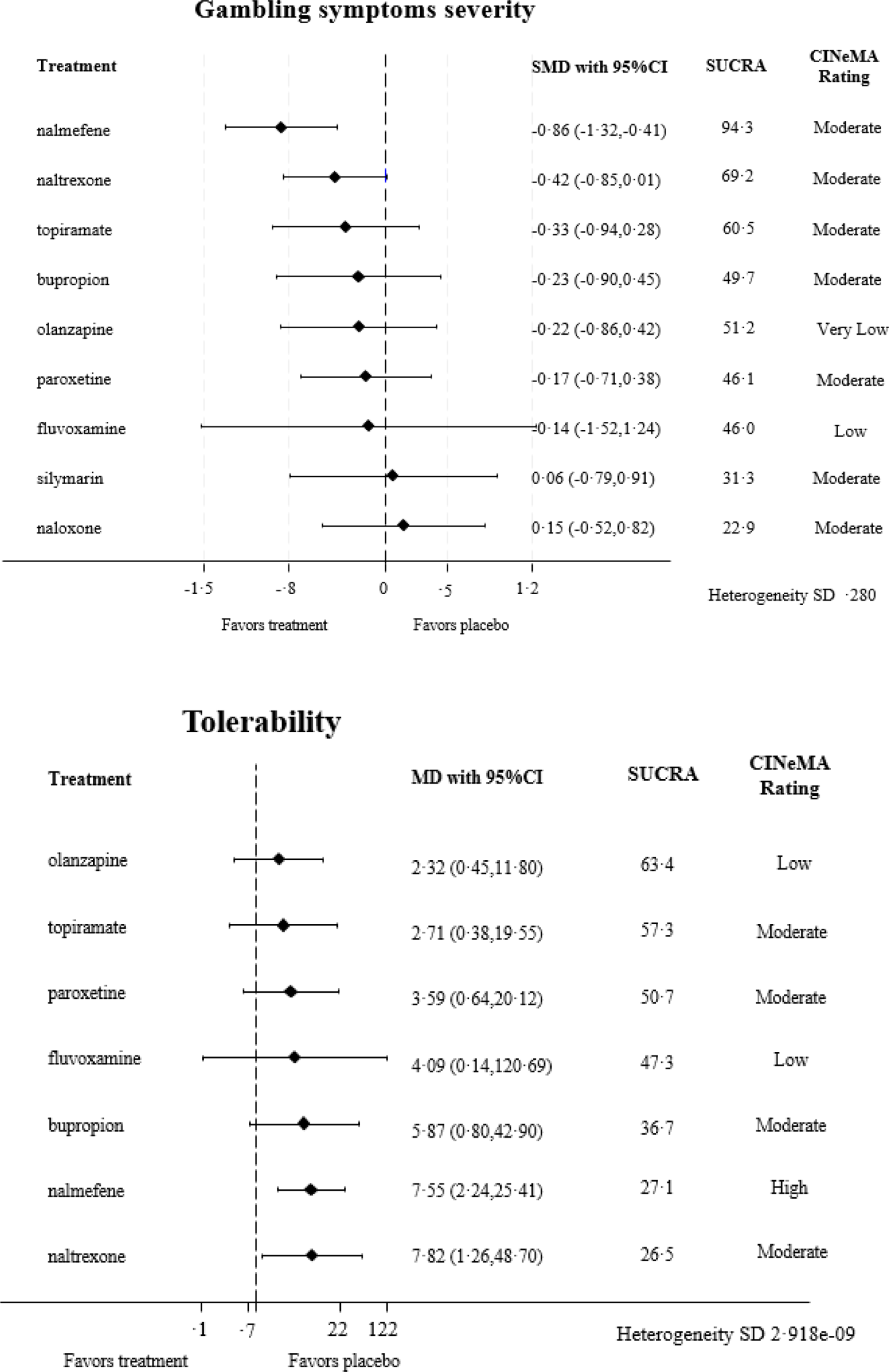
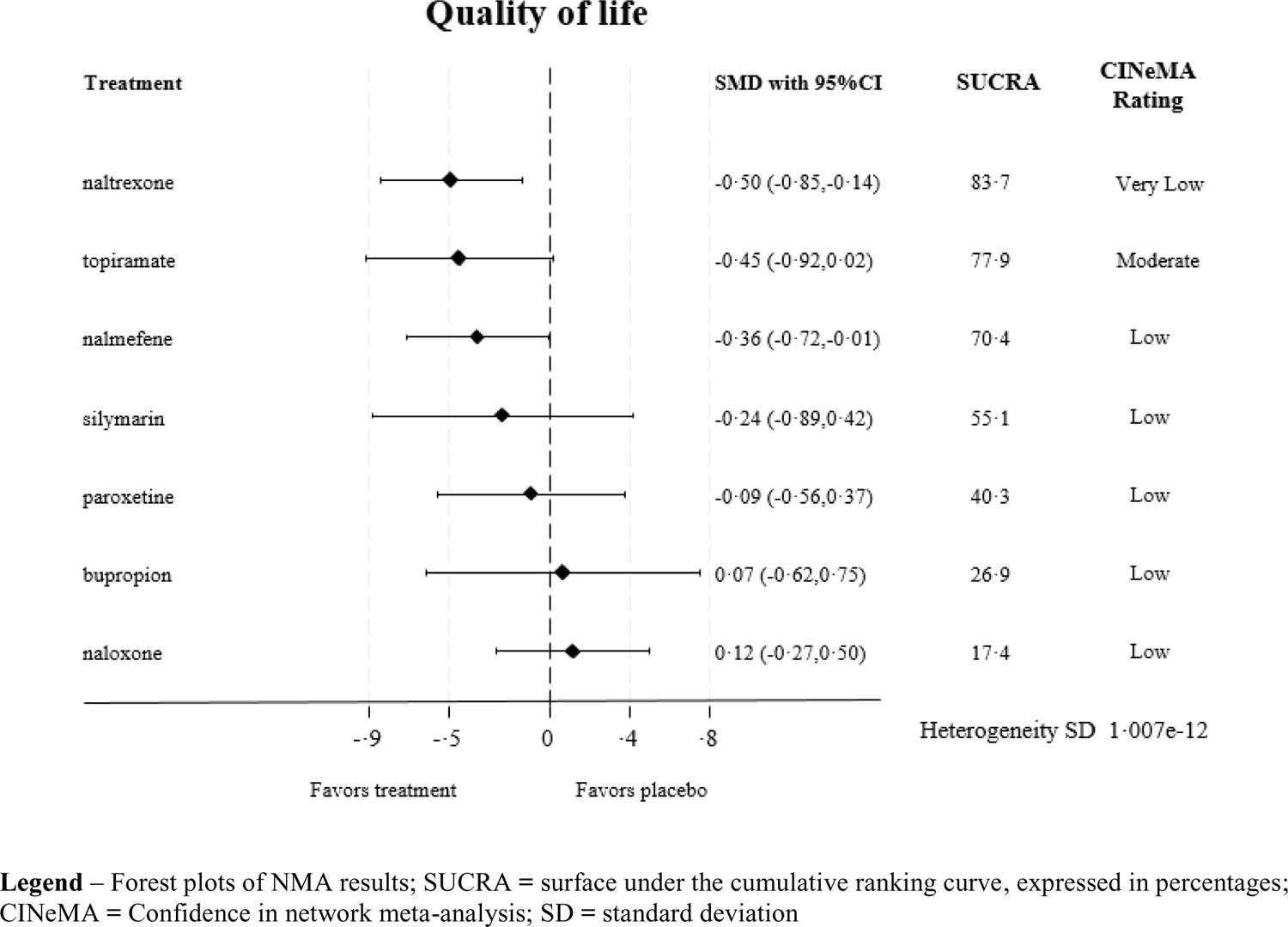
Forest plots of Network Meta-Analysis results for gambling severity, tolerability and quality of life

**Table 1.**
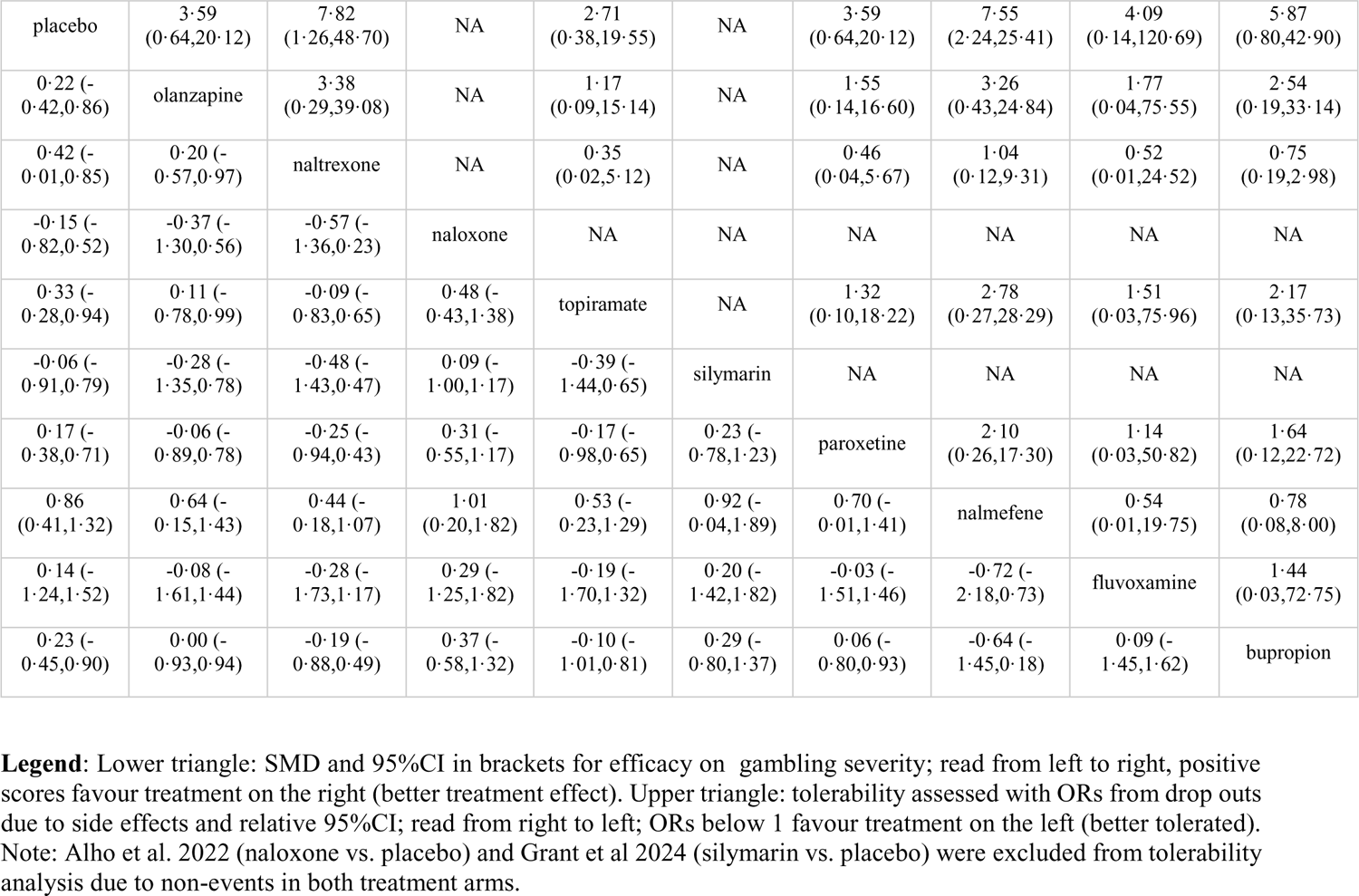
NMA results for all possible comparisons for gambling severity (lower triangle) and tolerability (upper triangle)

**Table 2.** NMA results for all possible comparisons for quality of life.

|  |  |  |  |  |  |  |  |
| --- | --- | --- | --- | --- | --- | --- | --- |
| topiramate |  |  |  |  |  |  |  |
| -0.45 (-<br>0.92,0.02) | placebo |  |  |  |  |  |  |
| -0.36 (-<br>1.02,0.30) | 0.09 (-<br>0.37,0.56) | paroxetine |  |  |  |  |  |
| 0.04 (-<br>0.55,0.63) | 0.50<br>(0.14,0.85) | 0.40 (-<br>0.18,0.99) | naltrexone |  |  |  |  |
| -0.57 (-<br>1.18,0.03) | -0.12 (-<br>0.50,0.27) | -0.21 (-<br>0.81,0.39) | -0.61 (-1.13,-<br>0.09) | naloxone |  |  |  |
| -0.09 (-<br>0.68,0.50) | 0.36<br>(0.01,0.72) | 0.27 (-<br>0.32,0.86) | -0.13 (-<br>0.64,0.37) | 0.48 (-<br>0.04,1.00) | nalmefene |  |  |
| -0.22 (-<br>1.02,0.59) | 0.24 (-<br>0.42,0.89) | 0.15 (-<br>0.66,0.95) | -0.26 (-<br>1.00,0.48) | 0.35 (-<br>0.40,1.11) | -0.12 (-<br>0.87,0.62) | silymarin |  |
| -0.52 (-<br>1.35,0.31) | -0.07 (-<br>0.75,0.62) | -0.16 (-<br>0.99,0.67) | -0.56 (-<br>1.33,0.21) | 0.05 (-<br>0.73,0.84) | -0.43 (-<br>1.20,0.34) | -0.30 (-<br>1.25,0.64) | bupropion |

**Legend:** Lower triangle: SMD and 95%CI in brackets for quality of life; read from left to right, positive scores favour treatment on the right (better treatment effect).

Heterogeneity measures (i.e., common standard deviation heterogeneity estimates) and results from incoherence assessment for all outcomes are presented in the appendix (pp 34-37). We found low heterogeneity within each network and we did not find evidence of incoherence. In ROB-MEN, since all comparisons had fewer than 10 studies, the construction of funnel plots and testing for small-study effects was not possible.

Treatment ranking measures based on NMA results are presented in detail in the appendix (p 38) for primary outcomes. Sensitivity analysis for low risk of bias studies was not possible due to all available studies having scored “some concerns” or above.

### Confidence in network meta-analysis estimates based on CINeMA

The confidence in the NMA estimates ranged between very low to high. Moderate confidence was assigned to most of the comparisons versus placebo. In particular, we were moderately confident that nalmefene and naltrexone were the most effective when compared to placebo. We were highly confident that nalmefene was the least tolerated treatment when compared to placebo. The main reasons for downgrading confidence were within-study-bias, imprecision and heterogeneity. Full CINeMA assessment for gambling severity, tolerability and quality of life are presented in the appendix (pp 39-47).

## DISCUSSION

This is the first NMA of RCTs for the pharmacological management of gambling disorder. We found moderate confidence evidence indicating that the opioid antagonist nalmefene was the most efficacious treatment in reducing gambling symptom severity. The other opioid antagonist often used in clinical practice, naltrexone, had moderate confidence evidence of being the second most efficacious treatment and the highest probability of improving quality of life, but with very low confidence in the evidence. However, both these treatments were associated with significantly higher dropout, due to side effects, than placebo, with confidence of the evidence from moderate (naltrexone) to high (nalmefene). Both nalmefene (in gambling severity, low confidence) and naltrexone (in quality of life, low confidence) showed superiority against naloxone in indirect comparisons. We did not find any other significant differences among medications. Naloxone did not differentiate from placebo in the one available study.^30^ Notably, naloxone is an opioid antagonist but has a very short duration of effect (short half-life),^31^ so lack of efficacy is perhaps unsurprising. Overall, these results provide useful insights that can support clinical decision-making around the pharmacological management of gambling. There are differences between opioid receptor antagonists in how they bind to brain receptors. For example, naltrexone has a preference for mu opioid receptors (MOR) and binds to a lesser extend to kappa opioid receptors (KOR), while nalmefene binds with similar strength to MOR and KOR.^31^ Such differences could theoretically influence treatment effects and tolerability profiles, but ultimately, both medications are thought to dampen dopamine neurotransmission in the nucleus accumbens and associated motivational neurocircuitry, reducing gambling excitement and craving.^32^

Contrary to a previous Cochrane review,^9^ this NMA showed that olanzapine was not statistically better than placebo in terms of primary or secondary efficacy outcomes. Due to combining direct and indirect evidence, as well as head-to-head comparisons, an NMA analysis may provide more accurate and complete results as compared to a conventional pairwise meta-analysis. Our NMA indicated that olanzapine is a less suitable treatment option for gambling disorder than previously considered, due to lack of efficacy. We found low confidence evidence indicating that olanzapine was the most well-tolerated among treatments. This is in contrast with the side effect profile reported more widely in the psychiatric literature.^33^ However, considering the lack of evidence of its efficacy, a preferable tolerability profile becomes less relevant.

Furthermore, we observed moderate confidence evidence that topiramate was not different from placebo in NMA on any of the primary or secondary outcomes, however it achieved relatively good rankings in all outcomes. Topiramate has a complex pharmacology and has been used in other areas of addiction psychiatry. Therefore, given the relatively good rankings, topiramate might merit further evaluation in clinical trials before being dismissed as a treatment option for gambling disorder.

### Consideration on studies identified in the systematic review but not included in NMA

Other treatments that have been used in addiction psychiatry have attracted interest for RCTs in gambling disorder, such as baclofen and acamprosate.^34^ Those studies were negative and their reports did not provide enough data for them to be included in the current NMA. This was also the case for sertraline;^35^ interestingly the sertraline RCT was characterized by >70% response in both active treatment and placebo groups.

Two other studies (on lithium^28^ and n-acetyl cysteine [NAC],^36^ respectively) were excluded from our NMA, as their studies involved recruitment of patients required to have a particular single named disorder in addition to gambling disorder, which violated the transitivity assumption necessary for inclusion. In an RCT examining lithium monotherapy for pathological gamblers with bipolar spectrum disorders,^28^ lithium was beneficial in reducing both gambling symptoms and mania scores, versus placebo. Lithium would thus appear to be a potentially useful option for gambling, which is comorbid with bipolar spectrum disorders during a mood episode. However it remains to be seen whether this would also apply to gambling disorder without this comorbidity. NAC was used successfully in one open label study^37^ and then one placebo-controlled RCT^38^ which included participants with a primary nicotine dependence diagnosis and gambling disorder (i.e. both were mandatory). Nicotine dependence has been identified as amongst the most common mental health comorbidity in gamblers.^39^ While the trial at the end of treatment (12 weeks) did not show any benefit for NAC for gambling (though it did, interestingly, in terms of reducing symptoms of nicotine dependence), the 24 week follow-up indicated that those previously treated with NAC had lower gambling severity than those previously treated with placebo. NAC is very well tolerated treatment and has been successfully used in impulse control disorders,^40^ which share many pathophysiology characteristics with gambling disorder. Further studies are warranted to assess the efficacy of NAC in gambling disorder.

### Limitations

Several limitations should be considered, reflecting both limitations of the included RCTs and of our NMA. In terms of limitations of the included RCTs, 81·25% [13/16] of them were judged of moderate quality and 18·75% [3/16] of low quality at the RoB2, mainly due to concerns over bias in the selection of reported result, but also missing outcome data. This reflects the state of the field and the fact that gambling disorder has been relatively neglected as compared to other mental health conditions, with most studies having been completed more than a decade ago. High quality RCTs are scarce and, in our view, this highlights further the importance of this NMA analysis in guiding the design of future RCTs. Moreover, the vast majority of the samples included both genders of middle aged participants. Differential gender effects could not be examined and the results cannot be extrapolated to other age groups e.g. olden people or children and adolescents. Furthermore, while we did not detect significant heterogeneity across studies overall, there were some methodological differences among studies that need to be considered. Different dosing schemes could have accounted for some heterogeneity – for example, Kovanen et al.^41^ used as required (PRN) dosing of naltrexone, as opposed to previous studies which used daily schemes with titration to a maximum tolerated dose using clinical judgement.^17,42^ Similarly, the two nalmefene studies used different dosing schemes (20mg/40mg vs. 25mg/50mg/100mg),^43,44^ and dose can potentially influence efficiency and tolerability outcomes, including dropout rates. In terms of limitations of our NMA, due to the presence of few closed loops in our network, we were not able to assess incoherence in all areas of our network. In the loops assessed we did not find evidence of incoherence. Moreover, due to the limited number of available studies, we were not able to meaningfully perform meta-regression to assess whether duration of treatment or mode of administration or dosing moderated effects in our network. However, most of the included studies had a similar duration of treatment between 12-20 weeks and used oral treatments. Furthermore, we did not have enough data to test effect modifiers like study sponsorship, comorbid psychiatric conditions, and mean baseline severity. Similarly, due to the small overall number of studies, we were not able to produce funnel plots to graphically identify reporting bias in ROB-MEN; while we could not detect such biases, this does not mean that those do not exist. Finally, while NMAs, in general, do not generate randomized evidence, they do provide observational evidence and helpful insights into the clinical dilemma of choosing between pharmacological options. Therefore, while NMA allows for indirect head-to-head comparisons, those could theoretically be better estimated from real-life RCTs. In practice, though, this can be prohibitively expensive and time consuming.

### Implications of the results for practice, policy, and future research

The current NMA provides the highest level of evidence synthesis to inform clinical practice as well as national and international treatment guidelines in terms of pharmacological options that should be recommended for gambling disorder. Based on the NMA findings, nalmefene and naltrexone should currently be regarded as having the best available evidence for efficacy in the treatment of gambling disorder. In the context of other disorders, it has been argued that nalmefene may have advantages over naltrexone in terms of its bioavailability, ability to bind differentially to particular brain opioid receptors (stronger affinity to kappa opioid receptors, mechanistically relevant in exerting antidepressant effects, but can also alter the side effect profile),^45,46^ and its apparent absence of dose-dependent liver toxicity (for discussion see Soyka).^47^ However, in the absence of direct head-to-head comparisons of these two medications in gambling disorder, and rigorous health-economic evaluations, we would suggest that both are retained as key first-line pharmacological treatment options. Retaining several options may also reduce the likelihood of patients having no feasible pharmacological treatment option – such as if one medication is not available in a particular geographical area/country, or in the case of supply disruptions.

Given the limited number of treatment options identified in the current NMA, and the high public health priority of gambling disorder,^3,48^ further large-scale clinical trials are urgently needed in relation to these and other medications for gambling disorder. A major reason for the low yearly rate (<1/year) of pharmacological RCTs for gambling disorder over the past decade, and relatively small number of studies accrued over time, is the lack of independent research funding being made available. Therefore, we urgently call on national governments and funding bodies to support independent clinical trials into gambling disorder. A summary of recommendations for future clinical trials for gambling disorder are made in Box 1, based on the findings of this review.

#### BOX 1

##### RECOMMENDATIONS FOR FUTURE CLINICAL TRIALS OF PHARMACOLOGICAL INTERVENTIONS FOR GAMBLING

- Full data reporting and adherence to intention to treat principles.
- Fostering wider collaborations and research design and dissemination practices to support data synthesis using individual participant data (PID) is critical
- Standardization of treatment duration and follow up
- Nalmefene and naltrexone show the best efficacy profile, however they coupled with relatively lower tolerability versus other compounds or placebo. To address tolerability issues, clinicians should initially consider doses found to be effective but not in the high range e.g. 50mg-100mg for naltrexone, 50mg or less for nalmefene. At the same time, it should be appreciated that higher doses may be needed in particular cases, such as in treatment non-response or partial response.
- Topiramate ranked relatively highly in terms of efficacy and tolerability profile, but it was not statistically better than placebo. Further studies are warranted to determine if this is an issue with statistical power.

Due to its good tolerability profile, future studies should further examine NAC to assess its efficacy.

### Analytic code availability

Example STATA code and package information for NMA can be found in the appendix (p 50).

## Data Availability

All data produced in the present study are available upon reasonable request to the authors

## Acknowledgement

We would like to thank authors of published papers included in this network meta-analysis who responded to requests for additional information to enable the meta-analysis.

## Author contributions

KI, CT, JES, SC, SRC contributed to the design of the study; CT led the initial pre-registration; CT and JES led the search and screening process; KI, JES, CDG, SRC contributed to data collection. KI, JES, CDG and SRC had access to the data. KI and CDG conducted the NMA analysis and take responsibility for the integrity and accuracy of the data analysis itself. SHW and VP led and equally contributed on the RoB2 scoring. All authors have intellectually contributed and reviewed the final submitted manuscript. All authors accept responsibility for the conduct of the study and its integrity.

## APPENDIX - Ioannidis et al. (2024)

### §S1 Methods details

Below we provide additional clarifications on the methods. Such details were not deviations from the original protocol published in PROSPERO, but rather a more comprehensive presentation of the full methodology used here, for reasons of clarity and transparency.

#### Inclusion/exclusion criteria

The co-existence of an intervention e.g. psychosocial support, cognitive restructuring, motivational interviewing, imaginal desensitization, attendance to Gamblers Anonymous (GA) meetings or equivalent was not an exclusion criterion, as long as those interventions existed equally in all respective study arms. There was no study in the sample breaching these criteria.

The existence of comorbidities in the sample was not an exclusion criterion, however, if the study specifically included a sample with a primary psychiatric condition in the whole sample (main inclusion criterion), then these studies were to be excluded, as this would violate the transitivity assumption. This applied to two studies in the final set, which were excluded from NMA.^1,2^

The need to have outcomes measures at two time points was not an inclusion criterion for NMA, as NMA can be performed without baseline outcome measures. This applies universally to all NMAs but was not explicitly stated in the original protocol publication.

When there were no events in both arms in respect to the tolerability analysis, we opted to exclude those studies from NMA as per Cochrane Handbook 16.9.3 (studies with no events. ^3^

#### Data collection

Where total sample size (N) from Intention-To-Treat (ITT) analyses was provided then this was used; if this was not given, then N was taken from the number of completers of the study.

Data were collected from manuscript figures where this was deemed possible and appropriate.

We included multi-arm studies, however, we used the approach provided by Cochrane Handbook (5.1 - 7.7 Extracting study results and converting to the desired format) ^4^ Therefore, study arms randomizing the same compound at different doses were merged into a single arm for inclusion in the NMA.

We used the Cochrane Handbook approach (5.1 - 7.7 Extracting study results and converting to the desired format) ^4^ to reverse calculate standard deviations or standard errors from 95%CIs or p-values where appropriate.

#### Data extraction

Data extraction started on the 14th of Jan 2023. Two authors (KI, JES) extracted the following data from each included study: demographic (mean age, % male), study design (sample size, number of arms, parallel or cross-over design, pragmatic design, single or multicentre, placebo control, the role of sponsorship), medicinal (dose used, dosing scheme and route of administration), clinical (diagnostic criteria for gambling disorder, presence of co-morbidities) characteristics. For the NMA we also extracted primary and secondary efficacy outcomes (mean, standard deviation) and tolerability (number of drop outs from medication side-effects).

#### Main analysis

If a study reported results from multiple scales, we analysed the results from one scale by using the following hierarchy: PG-YBOCS; G-SAS, CGI-I. CGI reported as % of responders was not further considered in analyses, as this format was not accompanied by a measure of variability.

#### Meta-regression and subgroup analyses

We did not use duration of treatment for meta-regression analyses. Our approach was to collect the last reported observation as end-of-treatment and use that as the final end point. While theoretically differences in treatment duration can violate the transitivity assumption, the vast majority of studies in this sample used similar shorts durations therefore we assumed that the transitivity assumption was met (e.g. 12-16 weeks).

We did not use mode of administration (e.g. oral tabs, caps, inhalation/nasal spray, injection etc) of treatment for subgroup analyses because all studies used oral preparations, apart from one ^5^ which used Naloxone Nasal spray.

### §S2 PRISMA-NMA checklist

#### PRISMA NMA^6^ Checklist of Items to Include When Reporting A Systematic Review Involving a Network Meta-analysis

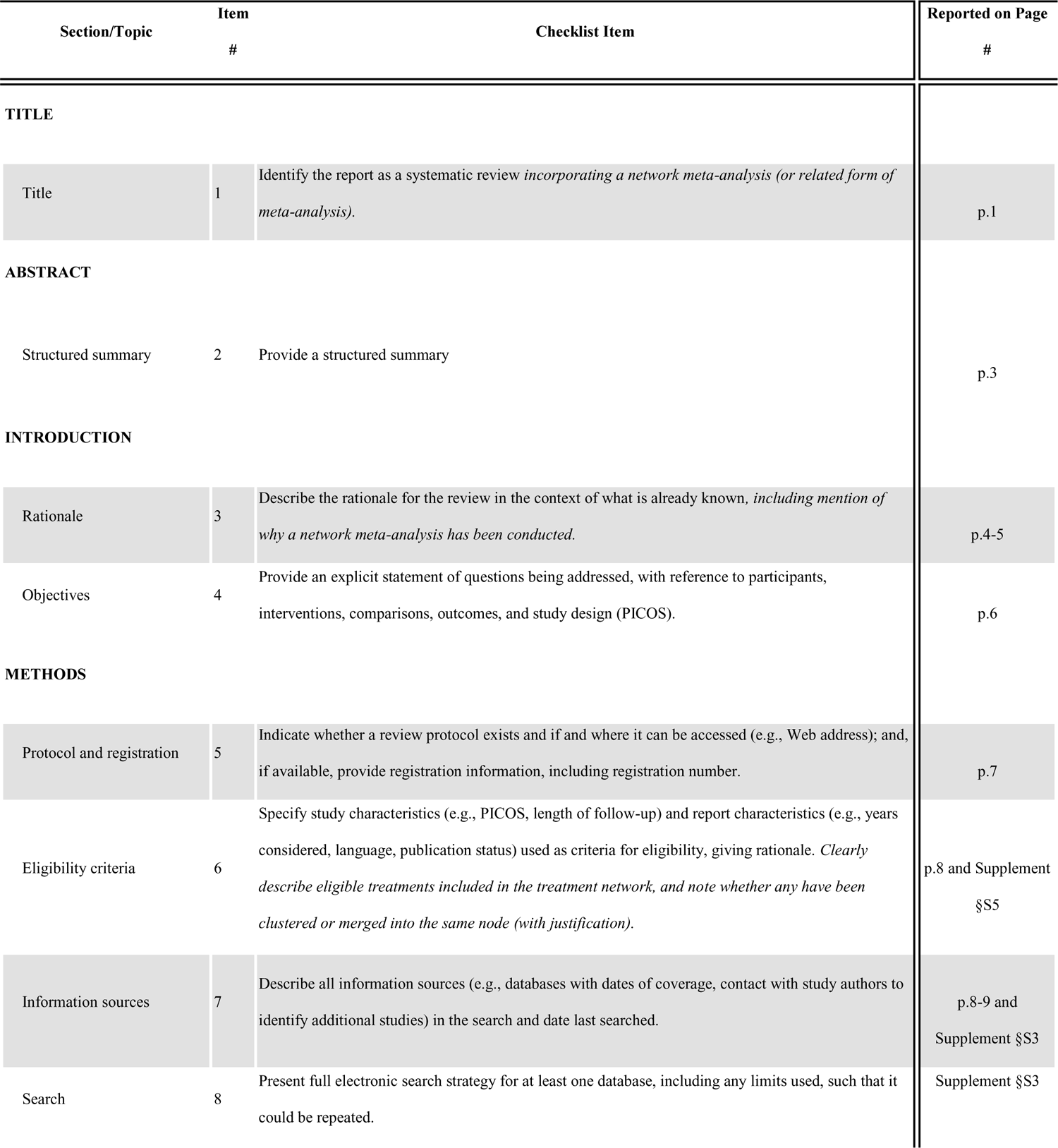
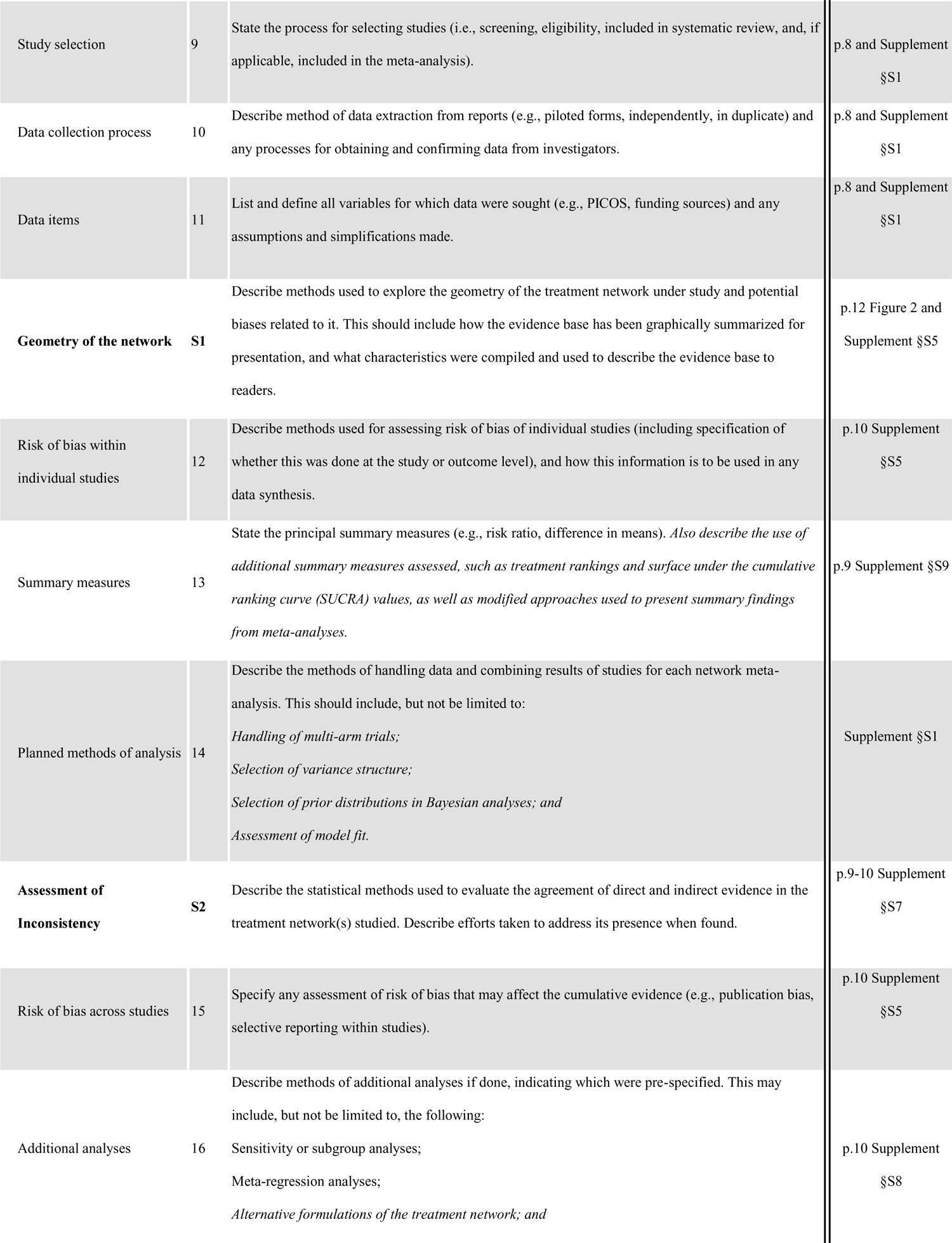
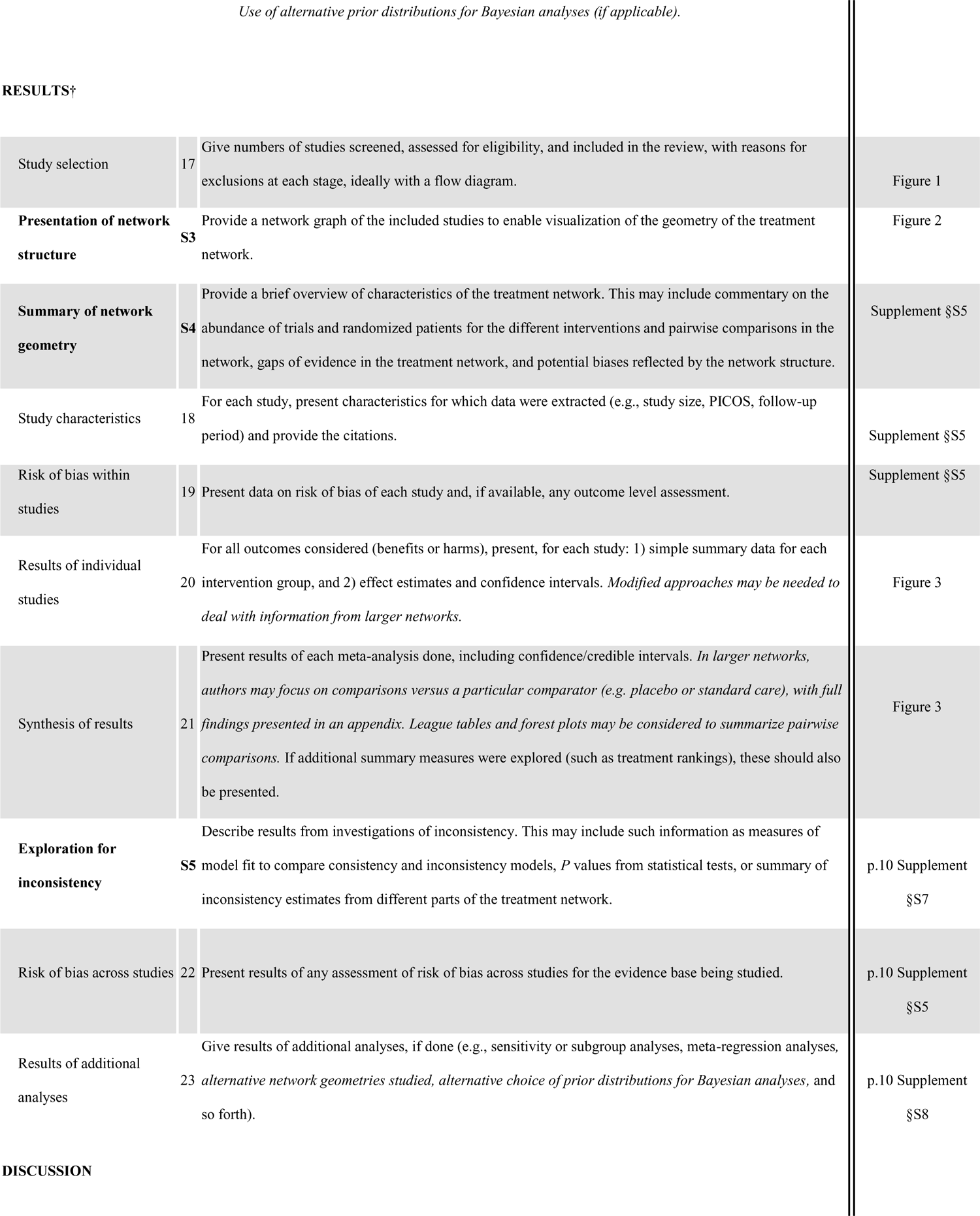
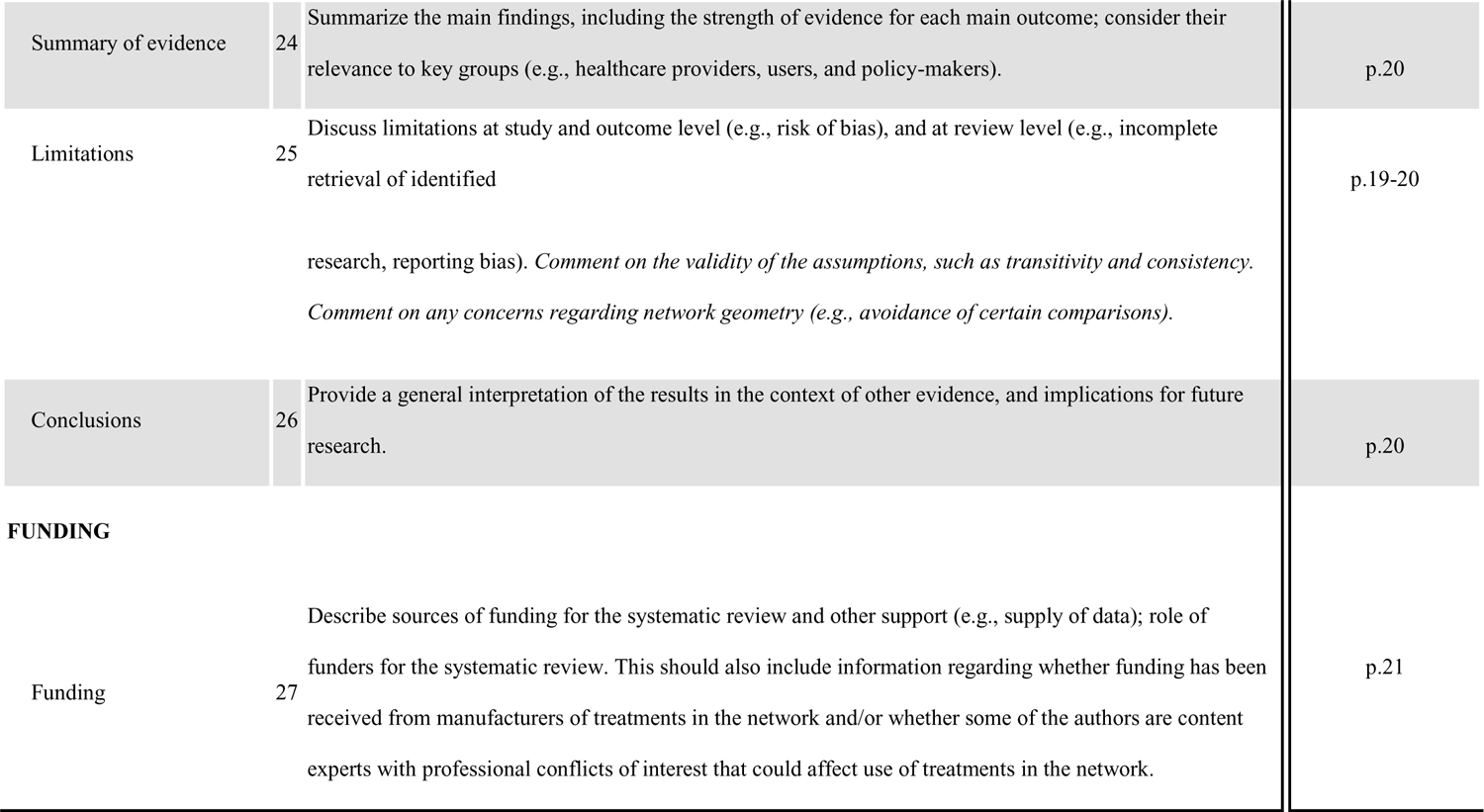

### §S3 Search strategy including online databases and clinical registers

Sources were searched included the following:

MEDLINE, EMBASE, PsycINFO, PubMed, CINAHL, AMED, and the Cochrane Database of Systematic Reviews, ERIC and Web of Science (including Science Citation Index Expanded (SCI-EXPANDED), Social Science Citation Index (SSCI), Conference Proceedings Citation Index-Science (CPCI-S) and Conference Proceedings Citation Index-Social Science and Humanities (CPCI-SSH)) via Web of Knowledge.

#### PubMed

((“Gambling”[Mesh] OR “Gambling”[tw] OR “Gambling Disorder”[tw] OR “Gambling Disorders”[tw] OR “Pathological Gambling”[tw] OR “Gambling Addiction”[tw] OR “Disordered Gambling”[tw] OR “Pathological Gambler”[tw] OR “Disordered Gambler”[tw] OR “Pathological Gamblers”[tw] OR “Disordered Gamblers”[tw] OR “ludomania”[tw]) AND (“Gambling/drug therapy”[Mesh] OR “Drug Therapy”[mesh] OR “Drug therapy”[tiab] OR “Drug therap*”[tiab] OR “Pharmacological Treatments”[tw] OR “Pharmacological Treatment”[tw] OR “Pharmacologic Treatments”[tw] OR “Pharmacologic Treatment”[tw] OR “Pharmacological Therapy”[tw] OR “Pharmacologic Therapy”[tw] OR “Pharmacotherapy”[tw] OR “Pharmacotherap*”[tw] OR “Psychopharmacotherapy”[tw] OR “Psychopharmacotherap*”[tw] OR “Drug treatment”[tw] OR “Drug treat*”[tw] OR “Medication”[tw] OR “Medications”[tw] OR “Medicat*”[tw] OR “Pharmaceutical Preparations”[Mesh] OR “pharmaco*”[tw] OR “Narcotic Antagonists”[Pharmacological Action] OR “Narcotic Antagonists”[mesh] OR “Dopamine Agents”[Mesh] OR “Dopamine Agents”[Pharmacological Action] OR “Hypnotics and Sedatives”[Pharmacological Action] OR “Hypnotics and Sedatives”[mesh] OR “Antidepressive Agents”[Pharmacological Action] OR “Antidepressive Agents”[mesh] OR “Antipsychotic Agents”[Mesh] OR “Antipsychotic Agents”[Pharmacological Action] OR “Catechol O-Methyltransferase Inhibitors”[mesh] OR “Catechol O-Methyltransferase Inhibitors”[PharmacologicalAction] OR “Psychotropic Drugs”[Mesh] OR “Psychotropic Drugs”[Pharmacological Action] OR “Alcohol Deterrents”[Mesh] OR “Alcohol Deterrents”[pharmacological action] OR “Anticonvulsants”[Pharmacological Action] OR “Anticonvulsants”[mesh] OR “Antiparkinson Agents”[Mesh] OR “Antiparkinson Agents”[Pharmacological Action] OR “Naloxone”[Mesh] OR “Naltrexone”[Mesh] OR “Topiramate”[Mesh] OR “Acetylcysteine”[Mesh] OR “Tolcapone”[Mesh] OR “nalmefene”[Supplementary Concept] OR “Narcotic Antagonists”[mesh] OR “Narcotic Antagonists”[Pharmacological Action] OR “Amantadine”[Mesh] OR “Memantine”[Mesh] OR “Modafinil”[Mesh] OR “Olanzapine”[Mesh] OR “Bupropion”[Mesh] OR “Escitalopram”[Mesh] OR “Serotonin Uptake Inhibitors”[Pharmacological Action] OR “Serotonin Uptake Inhibitors”[mesh] OR “Paroxetine”[Mesh] OR “Fluvoxamine”[Mesh] OR “Sertraline”[Mesh] OR “Lithium Carbonate”[Mesh] OR “Valproic Acid”[Mesh] OR “Clomipramine”[Mesh] OR “Antidepressive Agents, Tricyclic”[Pharmacological Action] OR “Antidepressive Agents, Tricyclic”[mesh] OR naloxone[tw] OR naltrexone[tw] OR topiramate[tw] OR n-acetylcysteine[tw] OR tolcapone[tw] OR nalmefene[tw] OR amantadine[tw] OR memantine[tw] OR modafinil[tw] OR olanzapine[tw] OR bupropion[tw] OR escitalopram[tw] OR paroxetine[tw] OR fluvoxamine[tw] OR sertraline[tw] OR lithium[tw] OR valproate[tw] OR clomipramine[tw] OR “Placebos”[Mesh] OR “Placebo Effect”[Mesh] OR “Placebos”[tw] OR “Placebo”[tw]) AND ((randomized controlled trial[pt] OR controlled clinical trial[pt] OR randomized[tiab] OR placebo[tiab] OR drug therapy[sh] OR randomly[tiab] OR trial[tiab] OR groups[tiab] NOT (animals [mh] NOT humans [mh])) OR (“Cross-Over Studies”[Mesh] OR “Crossover”[tw] OR “Cross over”[tw])))

#### MEDLINE (OVID)

((“Gambling“/ OR “Gambling“.mp OR “Gambling Disorder“.mp OR “Gambling Disorders“.mp OR“Pathological Gambling“.mp OR “Gambling Addiction“.mp OR “Disordered Gambling“.mp OR“Pathological Gambler“.mp OR “Disordered Gambler“.mp OR “Pathological Gamblers“.mp OR “Disordered Gamblers“.mp OR “ludomania“.mp) AND (“Gambling“/dt OR exp “Drug Therapy“/ OR “Drug therapy“.ti,ab OR “Drug therap*“.ti,ab OR “Pharmacological Treatments“.mp OR “Pharmacological Treatment“.mp OR “Pharmacologic Treatments“.mp OR “Pharmacologic Treatment“.mp OR “Pharmacological Therapy“.mp OR “Pharmacologic Therapy“.mp “Pharmacotherapy“.mp OR “Pharmacotherap*“.mp OR “Psychopharmacotherapy“.mp OR “Psychopharmacotherap*“.mp OR “Drug treatment“.mp OR “Drug treat*“.mp OR “Medication“.mp OR “Medications“.mp OR “Medicat*“.mp OR exp “Pharmaceutical Preparations“/ OR “pharmaco*“.mp OR exp “Naloxone“/ OR exp “Naltrexone“/ OR exp “Topiramate“/ OR exp “Acetylcysteine“/ OR exp “Tolcapone“/ OR exp “nalmefene“/ OR exp “Narcotic Antagonists“/ OR exp “Amantadine“/ OR exp “Memantine“/ OR exp “Modafinil“/ OR exp “Olanzapine“/ OR exp “Bupropion“/ OR exp “Escitalopram“/ OR exp “Serotonin Uptake Inhibitors“/ OR exp “Paroxetine“/ OR exp “Fluvoxamine“/ OR exp “Sertraline“/ OR exp “Lithium Carbonate“/ OR exp “Valproic Acid“/ OR exp “Clomipramine“/ OR exp “Antidepressive Agents, Tricyclic“/ OR naloxone.mp OR naltrexone.mp OR topiramate.mp OR n-acetylcysteine.mp OR tolcapone.mp OR nalmefene.mp OR amantadine.mp OR memantine.mp OR modafinil.mp OR olanzapine.mp OR bupropion.mp OR escitalopram.mp OR paroxetine.mp OR fluvoxamine.mp OR sertraline.mp OR lithium.mp OR valproate.mp OR clomipramine.mp OR exp “Narcotic Antagonists“/ OR exp “Dopamine Agents“/ OR exp “Hypnotics and Sedatives“/ OR exp “Antidepressive Agents“/ OR exp “Antipsychotic Agents“/ OR exp “Catechol O-Methyltransferase Inhibitors“/ OR exp “Psychotropic Drugs“/ OR exp “Alcohol Deterrents“/ OR exp “Anticonvulsants“/ OR exp “Antiparkinson Agents“/ OR exp “Placebos“/ OR exp “Placebo Effect“/ OR “Placebos“.mp OR “Placebo“.mp) AND ((exp randomized controlled trial/ OR exp controlled clinical trial/ OR randomized.ti,ab OR placebo.ti,ab OR exp drug therapy/ OR randomly.ti,ab OR trial.ti,ab OR groups.ti,ab NOT (exp animals/ NOT exp humans/)) OR (exp “Cross-Over Studies“/ OR “Crossover“.mp OR “Cross over“.mp)))

#### Embase (OVID)

((“Pathological Gambling“/ OR “Gambling“.mp OR “Gambling Disorder“.mp OR “Gambling Disorders“.mp OR “Pathological Gambling“.mp OR “Gambling Addiction“.mp OR “Disordered Gambling“.mp OR “Pathological Gambler“.mp OR “Disordered Gambler“.mp OR “Pathological Gamblers“.mp OR “Disordered Gamblers“.mp OR “ludomania“.mp) AND (“Pathological Gambling“/dt OR exp “Drug Therapy“/ OR “Drug therapy“.ti,ab OR “Drug therap*“.ti,ab OR “Pharmacological Treatments“.mp OR “Pharmacological Treatment“.mp OR “Pharmacologic Treatments“.mp OR “Pharmacologic Treatment“.mp OR “Pharmacological Therapy“.mp OR “Pharmacologic Therapy“.mp OR “Pharmacotherapy“.mp OR “Pharmacotherap*“.mp OR “Psychopharmacotherapy“.mp OR “Psychopharmacotherap*“.mp OR “Drug treatment“.mp OR “Drug treat*“.mp OR “Medication“.mp OR “Medications“.mp OR “Medicat*“.mp OR exp “Drug“/ OR “pharmaco*“.mp OR exp “Naloxone“/ OR exp “Naltrexone“/ OR exp “Topiramate“/ OR exp “Acetylcysteine“/ OR exp “Tolcapone“/ OR exp “nalmefene“/ OR exp “Narcotic Antagonist“/ OR exp “Amantadine“/ OR exp “Memantine“/ OR exp “Modafinil“/ OR exp “Olanzapine“/ OR exp “Bupropion“/ OR exp “Escitalopram“/ OR exp “Serotonin Uptake Inhibitor“/ OR exp “Paroxetine“/ OR exp “Fluvoxamine“/ OR exp “Sertraline“/ OR exp “Lithium Carbonate“/ OR exp “Valproic Acid“/ OR exp “Clomipramine“/ OR exp “Tricyclic Antidepressant Agent“/ OR naloxone.mp OR naltrexone.mp OR topiramate.mp OR n-acetylcysteine.mp OR tolcapone.mp OR nalmefene.mp OR amantadine.mp OR memantine.mp OR modafinil.mp OR olanzapine.mp OR bupropion.mp OR escitalopram.mp OR paroxetine.mp OR fluvoxamine.mp OR sertraline.mp OR lithium.mp OR valproate.mp OR clomipramine.mp OR exp “Narcotic Antagonist“/ OR exp “dopamine receptor stimulating agent“/ OR exp “Hypnotic Sedative Agent“/ OR exp “Antidepressant Agent“/ OR exp “Neuroleptic Agent“/ OR exp “Catechol OMethyltransferase Inhibitor“/ OR exp “Psychotropic Agent“/ OR exp “drugs used in the treatment of addiction“/ OR exp “Anticonvulsive Agent“/ OR exp “Antiparkinson Agent“/ OR exp “Placebo“/ OR exp “Placebo Effect“/ OR “Placebos“.mp OR “Placebo“.mp) AND ((Randomized controlled trial/ OR Controlled clinical study/ OR random$.ti,ab. OR randomization/ OR intermethod comparison/ OR placebo.ti,ab. OR (compare OR compared OR comparison).ti. OR ((evaluated OR evaluate OR evaluating OR assessed OR assess) and (compare OR compared OR comparing OR comparison)).ab. OR (open adj label).ti,ab. OR ((double OR single OR doubly OR singly) adj (blind OR blinded OR blindly)).ti,ab. OR double blind procedure/ OR parallel group$1.ti,ab. OR (crossover OR cross over).ti,ab. OR ((assign$ OR match OR matched OR allocation) adj5 (alternate OR group$1 OR intervention$1 OR patient$1 OR subject$1 OR participant$1)).ti,ab. OR (assigned OR allocated).ti,ab. OR (controlled adj7 (study OR design OR trial)).ti,ab. OR (volunteer OR volunteers).ti,ab. OR human experiment/ OR trial.ti.) not (((random$ adj sampl$ adj7 (“cross section $“ OR questionnaire$1 OR survey$ OR database$1)).ti,ab. not (comparative study/ OR controlled study/ OR randomi?ed controlled.ti,ab. OR randomly assigned.ti,ab.)) OR (Cross-sectional study/ not (randomized controlled trial/ OR controlled clinical study/ OR controlled study/ OR randomi?ed controlled.ti,ab. OR control group$1.ti,ab.)) OR (((case adj control$) and random$) not randomi?ed controlled).ti,ab. OR (Systematic review not (trial OR study)).ti. OR (nonrandom$ not random$).ti,ab. OR “Random field$“.ti,ab. OR (random cluster adj3 sampl$).ti,ab. OR ((review.ab. and review.pt.) not trial.ti.) OR (“we searched“.ab. and (review.ti. OR review.pt.)) OR “update review“.ab. OR (databases adj4 searched).ab. OR ((rat OR rats OR mouse OR mice OR swine OR porcine OR murine OR sheep OR lambs OR pigs OR piglets OR rabbit OR rabbits OR cat OR cats OR dog OR dogs OR cattle OR bovine OR monkey OR monkeys OR trout OR marmoset$1).ti. and animal experiment/) OR (Animal experiment/ not (human experiment/ OR human/))) OR (exp “Crossover Procedure“/ OR “Crossover“.mp OR “Cross over“.mp)))

#### PsyclNFO (EbscoHOST)

((TI(“Pathological Gambling” OR “Gambling” OR “Gambling Disorder” OR “Gambling Disorders” OR “Pathological Gambling” OR “Gambling Addiction” OR “Disordered Gambling” OR “Pathological Gambler” OR “Disordered Gambler” OR “Pathological Gamblers” OR “Disordered Gamblers” OR “ludomania “) OR MA(“Pathological Gambling” OR “Gambling” OR “Gambling Disorder” OR “Gambling Disorders” OR “Pathological Gambling” OR “Gambling Addiction” OR “Disordered Gambling” OR “Pathological Gambler” OR “Disordered Gambler” OR “Pathological Gamblers” OR “Disordered Gamblers “) OR SU(“Pathological Gambling” OR “Gambling” OR “Gambling Disorder” OR “Gambling Disorders” OR “Pathological Gambling” OR “Gambling Addiction” OR “Disordered Gambling” OR “Pathological Gambler” OR “Disordered Gambler” OR “Pathological Gamblers” OR “Disordered Gamblers” OR “ludomania “) OR AB(“Pathological Gambling” OR “Gambling” OR “Gambling Disorder” OR “Gambling Disorders” OR “Pathological Gambling” OR “Gambling Addiction” OR “Disordered Gambling” OR “Pathological Gambler” OR “Disordered Gambler” OR “Pathological Gamblers” OR “Disordered Gamblers” OR “ludomania “)) AND (TI(“Drug Therapy” OR “Drug therapy” OR “Drug therap*” OR “Pharmacological Treatments” OR “Pharmacological Treatment” OR “Pharmacologic Treatments” OR “Pharmacologic Treatment” OR “Pharmacological Therapy” OR “Pharmacologic Therapy” OR “Pharmacotherapy” OR “Pharmacotherap*” OR “Psychopharmacotherapy” OR “Psychopharmacotherap*” OR “Drug treatment” OR “Drug treat*” OR “Medication” OR “Medications” OR “Medicat*” OR “pharmaco*” OR “Naloxone” OR “Naltrexone” OR “Topiramate” OR “Acetylcysteine” OR “Tolcapone” OR “nalmefene” OR “Narcotic Antagonist” OR “Amantadine” OR “Memantine” OR “Modafinil” OR “Olanzapine” OR “Bupropion” OR “Escitalopram” OR “Serotonin Uptake Inhibitor” OR “Paroxetine” OR “Fluvoxamine” OR “Sertraline” OR “Lithium Carbonate” OR “Valproic Acid” OR “Clomipramine” OR “Tricyclic Antidepressant Agent” OR naloxone OR naltrexone OR topiramate OR n-acetylcysteine OR tolcapone OR nalmefene OR amantadine OR memantine OR modafinil OR olanzapine OR bupropion OR escitalopram OR paroxetine OR fluvoxamine OR sertraline OR lithium OR valproate OR clomipramine OR “Narcotic Antagonist” OR “dopamine receptor stimulating agent” OR “Hypnotic Sedative Agent” OR “Antidepressant Agent” OR “Neuroleptic Agent” OR “Catechol OMethyltransferase Inhibitor” OR “Psychotropic Agent” OR “drugs used in the treatment of addiction” OR “Anticonvulsive Agent” OR “Antiparkinson Agent” OR “Placebo” OR “Placebo Effect” OR “Placebos” OR “Placebo “) OR MA(“Drug Therapy” OR “Drug therapy” OR “Drug therap*” OR “Pharmacological Treatments” OR “Pharmacological Treatment” OR “Pharmacologic Treatments” OR “Pharmacologic Treatment” OR “Pharmacological Therapy” OR “Pharmacologic Therapy” OR “Pharmacotherapy” OR “Pharmacotherap*” OR “Psychopharmacotherapy” OR “Psychopharmacotherap*” OR “Drug treatment” OR “Drug treat*” OR “Medication” OR “Medications” OR “Medicat*” OR “pharmaco*” OR “Naloxone” OR “Naltrexone” OR “Topiramate” OR “Acetylcysteine” OR “Tolcapone” OR “nalmefene” OR “Narcotic Antagonist” OR “Amantadine” OR “Memantine” OR “Modafinil” OR “Olanzapine” OR “Bupropion” OR “Escitalopram” OR “Serotonin Uptake Inhibitor” OR “Paroxetine” OR “Fluvoxamine” OR “Sertraline” OR “Lithium Carbonate” OR “Valproic Acid” OR “Clomipramine” OR “Tricyclic Antidepressant Agent” OR naloxone OR naltrexone OR topiramate OR n-acetylcysteine OR tolcapone OR nalmefene OR amantadine OR memantine OR modafinil OR olanzapine OR bupropion OR escitalopram OR paroxetine OR fluvoxamine OR sertraline OR lithium OR valproate OR clomipramine OR “Narcotic Antagonist” OR “dopamine receptor stimulating agent” OR “Hypnotic Sedative Agent” OR “Antidepressant Agent” OR “Neuroleptic Agent” OR “Catechol OMethyltransferase Inhibitor” OR “Psychotropic Agent” OR “drugs used in the treatment of addiction” OR “Anticonvulsive Agent” OR “Antiparkinson Agent” OR “Placebo” OR “Placebo Effect” OR “Placebos” OR “Placebo “) OR SU(“Drug Therapy” OR “Drug therapy” OR “Drug therap*” OR “Pharmacological Treatments” OR “Pharmacological Treatment” OR “Pharmacologic Treatments” OR “Pharmacologic Treatment” OR “Pharmacological Therapy” OR “Pharmacologic Therapy” OR “Pharmacotherapy” OR “Pharmacotherap*” OR “Psychopharmacotherapy” OR “Psychopharmacotherap*” OR “Drug treatment” OR “Drug treat*” OR “Medication” OR “Medications” OR “Medicat*” OR “pharmaco*” OR “Narcotic Antagonist” OR “dopamine receptor stimulating agent” OR “Hypnotic Sedative Agent” OR “Antidepressant Agent” OR “Neuroleptic Agent” OR “Catechol OMethyltransferase Inhibitor” OR “Psychotropic Agent” OR “drugs used in the treatment of addiction” OR “Anticonvulsive Agent” OR “Antiparkinson Agent” OR “Naloxone” OR “Naltrexone” OR “Topiramate” OR “Acetylcysteine” OR “Tolcapone” OR “nalmefene” OR “Narcotic Antagonist” OR “Amantadine” OR “Memantine” OR “Modafinil” OR “Olanzapine” OR “Bupropion” OR “Escitalopram” OR “Serotonin Uptake Inhibitor” OR “Paroxetine” OR “Fluvoxamine” OR “Sertraline” OR “Lithium Carbonate” OR “Valproic Acid” OR “Clomipramine” OR “Tricyclic Antidepressant Agent” OR naloxone OR naltrexone OR topiramate OR n-acetylcysteine OR tolcapone OR nalmefene OR amantadine OR memantine OR modafinil OR olanzapine OR bupropion OR escitalopram OR paroxetine OR fluvoxamine OR sertraline OR lithium OR valproate OR clomipramine OR “Placebo” OR “Placebo Effect” OR “Placebos” OR “Placebo “) OR AB(“Drug Therapy” OR “Drug therapy” OR “Drug therap*” OR “Pharmacological Treatments” OR “Pharmacological Treatment” OR “Pharmacologic Treatments” OR “Pharmacologic Treatment” OR “Pharmacological Therapy” OR “Pharmacologic Therapy” OR “Pharmacotherapy” OR “Pharmacotherap*” OR “Psychopharmacotherapy” OR “Psychopharmacotherap*” OR “Drug treatment” OR “Drug treat*” OR “Medication” OR “Medications” OR “Medicat*” OR “pharmaco*” OR “Narcotic Antagonist” OR “dopamine receptor stimulating agent” OR “Hypnotic Sedative Agent” OR “Antidepressant Agent” OR “Neuroleptic Agent” OR “Catechol OMethyltransferase Inhibitor” OR “Psychotropic Agent” OR “drugs used in the treatment of addiction” OR “Anticonvulsive Agent” OR “Antiparkinson Agent” OR “Naloxone” OR “Naltrexone” OR “Topiramate” OR “Acetylcysteine” OR “Tolcapone” OR “nalmefene” OR “Narcotic Antagonist” OR “Amantadine” OR “Memantine” OR “Modafinil” OR “Olanzapine” OR “Bupropion” OR “Escitalopram” OR “Serotonin Uptake Inhibitor” OR “Paroxetine” OR “Fluvoxamine” OR “Sertraline” OR “Lithium Carbonate” OR “Valproic Acid” OR “Clomipramine” OR “Tricyclic Antidepressant Agent” OR naloxone OR naltrexone OR topiramate OR n-acetylcysteine OR tolcapone OR nalmefene OR amantadine OR memantine OR modafinil OR olanzapine OR bupropion OR escitalopram OR paroxetine OR fluvoxamine OR sertraline OR lithium OR valproate OR clomipramine OR “Placebo” OR “Placebo Effect” OR “Placebos” OR “Placebo “)) AND (TI(randomized controlled trial OR controlled clinical trial OR randomized OR placebo OR “drug therapy” OR randomly OR trial OR groups OR “Cross-Over Studies” OR “Crossover” OR “Cross over “) OR MA(randomized controlled trial OR controlled clinical trial OR randomized OR placebo OR “drug therapy” OR randomly OR trial OR groups OR “Cross-Over Studies” OR “Crossover” OR “Cross over “) OR SU(randomized controlled trial OR controlled clinical trial OR randomized OR placebo OR “drug therapy” OR randomly OR trial OR groups OR “Cross-Over Studies” OR “Crossover” OR “Cross over “) OR AB(randomized controlled trial OR controlled clinical trial OR randomized OR placebo OR “drug therapy” OR randomly OR trial OR groups OR “Cross-Over Studies” OR “Crossover” OR “Cross over “)))

#### Emcare (OVID)

((“Pathological Gambling “/ OR “Gambling “.mp OR “Gambling Disorder “.mp OR “Gambling Disorders “.mp OR “Pathological Gambling “.mp OR “Gambling Addiction “.mp OR “Disordered Gambling “.mp OR “Pathological Gambler “.mp OR “Disordered Gambler “.mp OR “Pathological Gamblers “.mp OR “Disordered Gamblers “.mp OR “ludomania “.mp) AND (exp “Drug Therapy “/ OR “Drug therapy “.ti,ab OR “Drug therap* “.ti,ab OR “Pharmacological Treatments “.mp OR “Pharmacological Treatment “.mp OR “Pharmacologic Treatments “.mp OR “Pharmacologic Treatment “.mp OR “Pharmacological Therapy “.mp OR “Pharmacologic Therapy “.mp OR “Pharmacotherapy “.mp OR “Pharmacotherap* “.mp OR “Psychopharmacotherapy “.mp OR “Psychopharmacotherap* “.mp OR “Drug treatment “.mp OR “Drug treat* “.mp OR “Medication “.mp OR “Medications “.mp OR “Medicat* “.mp OR exp “Drug “/ OR “pharmaco* “.mp OR exp “Narcotic Antagonist “/ OR exp “dopamine receptor stimulating agent “/ OR exp “Hypnotic Sedative Agent “/ OR exp “Antidepressant Agent “/ OR exp “Neuroleptic Agent “/ OR exp “Catechol OMethyltransferase Inhibitor “/ OR exp “Psychotropic Agent “/ OR exp “drugs used in the treatment of addiction “/ OR exp “Anticonvulsive Agent “/ OR exp “Antiparkinson Agent “/ OR exp “Naloxone “/ OR exp “Naltrexone “/ OR exp “Topiramate “/ OR exp “Acetylcysteine “/ OR exp “Tolcapone “/ OR exp “nalmefene “/ OR exp “Narcotic Antagonist “/ OR exp “Amantadine “/ OR exp “Memantine “/ OR exp “Modafinil “/ OR exp “Olanzapine “/ OR exp “Bupropion “/ OR exp “Escitalopram “/ OR exp “Serotonin Uptake Inhibitor “/ OR exp “Paroxetine “/ OR exp “Fluvoxamine “/ OR exp “Sertraline “/ OR exp “Lithium Carbonate “/ OR,exp “Valproic Acid “/ OR exp “Clomipramine “/ OR exp “Tricyclic Antidepressant Agent “/ OR naloxone.mp OR naltrexone.mp OR topiramate.mp OR n-acetylcysteine.mp OR tolcapone.mp OR nalmefene.mp OR amantadine.mp OR memantine.mp OR modafinil.mp OR olanzapine.mp OR bupropion.mp OR escitalopram.mp OR paroxetine.mp OR fluvoxamine.mp OR sertraline.mp OR lithium.mp OR valproate.mp OR clomipramine.mp OR exp “Placebo “/ OR exp “Placebo Effect “/ OR “Placebos “.mp OR “Placebo “.mp) AND ((Randomized controlled trial/ OR Controlled clinical study/ OR random$.ti,ab. OR randomization/ OR intermethod comparison/ OR placebo.ti,ab. OR (compare OR compared OR comparison).ti. OR ((evaluated OR evaluate OR evaluating OR assessed OR assess) and (compare OR compared OR comparing OR comparison)).ab. OR (open adj label).ti,ab. OR ((double OR single OR doubly OR singly) adj (blind OR blinded OR blindly)).ti,ab. OR double blind procedure/ OR parallel group$1.ti,ab. OR (crossover OR cross over).ti,ab. OR ((assign$ OR match OR matched OR allocation) adj5 (alternate OR group$1 OR intervention$1 OR patient$1 OR subject$1 OR participant$1)).ti,ab. OR (assigned OR allocated).ti,ab. OR (controlled adj7 (study OR design OR trial)).ti,ab. OR (volunteer OR volunteers).ti,ab. OR human experiment/ OR trial.ti.) not (((random$ adj sampl$ adj7 (“cross section$” OR questionnaire$1 OR survey$ OR database$1)).ti,ab. Not (comparative study/ OR controlled study/ OR randomi?ed controlled.ti,ab. OR randomly assigned.ti,ab.)) OR (Cross-sectional study/ not (randomized controlled trial/ OR controlled clinical study/ OR controlled study/ OR randomi?ed controlled.ti,ab. OR control group$1.ti,ab.)) OR (((case adj control$) and random$) not randomi?ed controlled).ti,ab. OR (Systematic review not (trial OR study)).ti. OR (nonrandom$ not random$).ti,ab. OR “Random field$ “.ti,ab. OR (random cluster adj3 sampl$).ti,ab. OR ((review.ab. and review.pt.) not trial.ti.) OR (“we searched “.ab. and (review.ti. OR review.pt.)) OR “update review “.ab. OR (databases adj4 searched).ab. OR ((rat OR rats OR mouse OR mice OR swine OR porcine OR murine OR sheep OR lambs OR pigs OR piglets OR rabbit OR rabbits OR cat OR cats OR dog OR dogs OR cattle OR bovine OR monkey OR monkeys OR trout OR marmoset$1).ti. and animal experiment/) OR (Animal experiment/ not (human experiment/ OR human/))) OR (exp “Crossover Procedure “/ OR “Crossover “.mp OR “Cross over “.mp)))

#### Cochrane Library

((“Pathological Gambling” OR “Gambling” OR “Gambling Disorder” OR “Gambling Disorders” OR “Pathological Gambling” OR “Gambling Addiction” OR “Disordered Gambling” OR “Pathological Gambler” OR “Disordered Gambler” OR “Pathological Gamblers” OR “Disordered Gamblers” OR “ludomania “) AND (“Drug Therapy” OR “Drug therap*” OR “Pharmacological Treatments” OR “Pharmacological Treatment” OR “Pharmacologic Treatments” OR “Pharmacologic Treatment” OR “Pharmacological Therapy” OR “Pharmacologic Therapy” OR “Pharmacotherapy” OR “Pharmacotherap*” OR “Psychopharmacotherapy” OR “Psychopharmacotherap*” OR “Drug treatment” OR “Drug treat*” OR “Medication” OR “Medications” OR “Medicat*” OR “Pharmaceutical Preparations” OR “pharmaco*” OR “Narcotic Antagonists” OR “Narcotic Antagonists” OR “Dopamine Agents” OR “Dopamine Agents” OR “Hypnotics and Sedatives” OR “Hypnotics and Sedatives” OR “Antidepressive Agents” OR “Antidepressive Agents” OR “Antipsychotic Agents” OR “Antipsychotic Agents” OR “Catechol O-Methyltransferase Inhibitors” OR “Catechol O-Methyltransferase Inhibitors” OR “Psychotropic Drugs” OR “Psychotropic Drugs” OR “Alcohol Deterrents” OR “Alcohol Deterrents” OR “Anticonvulsants” OR “Anticonvulsants” OR “Antiparkinson Agents” OR “Antiparkinson Agents” OR “Naloxone” OR “Naltrexone” OR “Topiramate” OR “Acetylcysteine” OR “Tolcapone” OR “nalmefene” OR “Narcotic Antagonist” OR “Amantadine” OR “Memantine” OR “Modafinil” OR “Olanzapine” OR “Bupropion” OR “Escitalopram” OR “Serotonin Uptake Inhibitor” OR “Paroxetine” OR “Fluvoxamine” OR “Sertraline” OR “Lithium Carbonate” OR “Valproic Acid” OR “Clomipramine” OR “Tricyclic Antidepressant Agent” OR naloxone OR naltrexone OR topiramate OR n-acetylcysteine OR tolcapone OR nalmefene OR amantadine OR memantine OR modafinil OR olanzapine OR bupropion OR escitalopram OR paroxetine OR fluvoxamine OR sertraline OR lithium OR valproate OR clomipramine OR “Placebos” OR “Placebo Effect” OR “Placebos” OR “Placebo “))

#### ERIC (EbscoHOST)

((TI(“Pathological Gambling” OR “Gambling” OR “Gambling Disorder” OR “Gambling Disorders” OR “Pathological Gambling” OR “Gambling Addiction” OR “Disordered Gambling” OR “Pathological Gambler” OR “Disordered Gambler” OR “Pathological Gamblers” OR “Disordered Gamblers” OR “ludomania “) OR SU(“Pathological Gambling” OR “Gambling” OR “Gambling Disorder” OR “Gambling Disorders” OR “Pathological Gambling” OR “Gambling Addiction” OR “Disordered Gambling” OR “Pathological Gambler” OR “Disordered Gambler” OR “Pathological Gamblers” OR “Disordered Gamblers” OR “ludomania “) OR AB(“Pathological Gambling” OR “Gambling” OR “Gambling Disorder” OR “Gambling Disorders” OR “Pathological Gambling” OR “Gambling Addiction” OR “Disordered Gambling” OR “Pathological Gambler” OR “Disordered Gambler” OR “Pathological Gamblers” OR “Disordered Gamblers” OR “ludomania “)) AND (TI(“Drug Therapy” OR “Drug therapy” OR “Drug therap*” OR “Pharmacological Treatments” OR “Pharmacological Treatment” OR “Pharmacologic Treatments” OR “Pharmacologic Treatment” OR “Pharmacological Therapy” OR “Pharmacologic Therapy” OR “Pharmacotherapy” OR “Pharmacotherap*” OR “Psychopharmacotherapy” OR “Psychopharmacotherap*” OR “Drug treatment” OR “Drug treat*” OR “Medication” OR “Medications” OR “Medicat*” OR “pharmaco*” OR “Narcotic Antagonist” OR “dopamine receptor stimulating agent” OR “Hypnotic Sedative Agent” OR “Antidepressant Agent” OR “Neuroleptic Agent” OR “Catechol OMethyltransferase Inhibitor” OR “Psychotropic Agent” OR “drugs used in the treatment of addiction” OR “Anticonvulsive Agent” OR “Antiparkinson Agent” OR “Naloxone” OR “Naltrexone” OR “Topiramate” OR “Acetylcysteine” OR “Tolcapone” OR “nalmefene” OR “Narcotic Antagonist” OR “Amantadine” OR “Memantine” OR “Modafinil” OR “Olanzapine” OR “Bupropion” OR “Escitalopram” OR “Serotonin Uptake Inhibitor” OR “Paroxetine” OR “Fluvoxamine” OR “Sertraline” OR “Lithium Carbonate” OR “Valproic Acid” OR “Clomipramine” OR “Tricyclic Antidepressant Agent” OR naloxone OR naltrexone OR topiramate OR n-acetylcysteine OR tolcapone OR nalmefene OR amantadine OR memantine OR modafinil OR olanzapine OR bupropion OR escitalopram OR paroxetine OR fluvoxamine OR sertraline OR lithium OR valproate OR clomipramine OR “Placebo” OR “Placebo Effect” OR “Placebos” OR “Placebo “) OR SU(“Drug Therapy” OR “Drug therapy” OR “Drug therap*” OR “Pharmacological Treatments” OR “Pharmacological Treatment” OR “Pharmacologic Treatments” OR “Pharmacologic Treatment” OR “Pharmacological Therapy” OR “Pharmacologic Therapy” OR “Pharmacotherapy” OR “Pharmacotherap*” OR “Psychopharmacotherapy” OR “Psychopharmacotherap*” OR “Drug treatment” OR “Drug treat*” OR “Medication” OR “Medications” OR “Medicat*” OR “pharmaco*” OR “Narcotic Antagonist” OR “dopamine receptor stimulating agent” OR “Hypnotic Sedative Agent” OR “Antidepressant Agent” OR “Neuroleptic Agent” OR “Catechol OMethyltransferase Inhibitor” OR “Psychotropic Agent” OR “drugs used in the treatment of addiction” OR “Anticonvulsive Agent” OR “Antiparkinson Agent” OR “Naloxone” OR “Naltrexone” OR “Topiramate” OR “Acetylcysteine” OR “Tolcapone” OR “nalmefene” OR “Narcotic Antagonist” OR “Amantadine” OR “Memantine” OR “Modafinil” OR “Olanzapine” OR “Bupropion” OR “Escitalopram” OR “Serotonin Uptake Inhibitor” OR “Paroxetine” OR “Fluvoxamine” OR “Sertraline” OR “Lithium Carbonate” OR “Valproic Acid” OR “Clomipramine” OR “Tricyclic Antidepressant Agent” OR naloxone OR naltrexone OR topiramate OR n-acetylcysteine OR tolcapone OR nalmefene OR amantadine OR memantine OR modafinil OR olanzapine OR bupropion OR escitalopram OR paroxetine OR fluvoxamine OR sertraline OR lithium OR valproate OR clomipramine OR “Placebo” OR “Placebo Effect” OR “Placebos” OR “Placebo “) OR AB(“Drug Therapy” OR “Drug therapy” OR “Drug therap*” OR “Pharmacological Treatments” OR “Pharmacological Treatment” OR “Pharmacologic Treatments” OR “Pharmacologic Treatment” OR “Pharmacological Therapy” OR “Pharmacologic Therapy” OR “Pharmacotherapy” OR “Pharmacotherap*” OR “Psychopharmacotherapy” OR “Psychopharmacotherap*” OR “Drug treatment” OR “Drug treat*” OR “Medication” OR “Medications” OR “Medicat*” OR “pharmaco*” OR “Narcotic Antagonist” OR “dopamine receptor stimulating agent” OR “Hypnotic Sedative Agent” OR “Antidepressant Agent” OR “Neuroleptic Agent” OR “Catechol OMethyltransferase Inhibitor” OR “Psychotropic Agent” OR “drugs used in the treatment of addiction” OR “Anticonvulsive Agent” OR “Antiparkinson Agent” OR “Naloxone” OR “Naltrexone” OR “Topiramate” OR “Acetylcysteine” OR “Tolcapone” OR “nalmefene” OR “Narcotic Antagonist” OR “Amantadine” OR “Memantine” OR “Modafinil” OR “Olanzapine” OR “Bupropion” OR “Escitalopram” OR “Serotonin Uptake Inhibitor” OR “Paroxetine” OR “Fluvoxamine” OR “Sertraline” OR “Lithium Carbonate” OR “Valproic Acid” OR “Clomipramine” OR “Tricyclic Antidepressant Agent” OR naloxone OR naltrexone OR topiramate OR n-acetylcysteine OR tolcapone OR nalmefene OR amantadine OR memantine OR modafinil OR olanzapine OR bupropion OR escitalopram OR paroxetine OR fluvoxamine OR sertraline OR lithium OR valproate OR clomipramine OR “Placebo” OR “Placebo Effect” OR “Placebos” OR “Placebo “)) AND (TI(randomized controlled trial OR controlled clinical trial OR randomized OR placebo OR “drug therapy” OR randomly OR trial OR groups OR “Cross-Over Studies” OR “Crossover” OR “Cross over “) OR MA(randomized controlled trial OR controlled clinical trial OR randomized OR placebo OR “drug therapy” OR randomly OR trial OR groups OR “Cross-Over Studies” OR “Crossover” OR “Cross over “) OR AB(randomized controlled trial OR controlled clinical trial OR randomized OR placebo OR “drug therapy” OR randomly OR trial OR groups OR “Cross-Over Studies” OR “Crossover” OR “Cross over “)))

#### Web of Science

((TI=(“Pathological Gambling” OR “Gambling” OR “Gambling Disorder” OR “Gambling Disorders” OR “Pathological Gambling” OR “Gambling Addiction” OR “Disordered Gambling” OR “Pathological Gambler” OR “Disordered Gambler” OR “Pathological Gamblers” OR “Disordered Gamblers” OR “ludomania “) OR AK=(“Pathological Gambling” OR “Gambling” OR “Gambling Disorder” OR “Gambling Disorders” OR “Pathological Gambling” OR “Gambling Addiction” OR “Disordered Gambling” OR “Pathological Gambler” OR “Disordered Gambler” OR “Pathological Gamblers” OR “Disordered Gamblers” OR “ludomania “) OR AB=(“Pathological Gambling” OR “Gambling” OR “Gambling Disorder” OR “Gambling Disorders” OR “Pathological Gambling” OR “Gambling Addiction “OR “Disordered Gambling” OR “Pathological Gambler” OR “Disordered Gambler” OR “PathologicalGamblers” OR “Disordered Gamblers” OR “ludomania “)) AND (TI=(“Drug Therapy” OR “Drug therapy “OR “Drug therap*” OR “Pharmacological Treatments” OR “Pharmacological Treatment” OR “Pharmacologic Treatments” OR “Pharmacologic Treatment” OR “Pharmacological Therapy” OR “Pharmacologic Therapy” OR “Pharmacotherapy” OR “Pharmacotherap*” OR “Psychopharmacotherapy” OR “Psychopharmacotherap*” OR “Drug treatment” OR “Drug treat*” OR “Medication” OR “Medications” OR “Medicat*” OR “pharmaco*” OR “Narcotic Antagonist” OR “dopamine receptor stimulating agent” OR “Hypnotic Sedative Agent” OR “Antidepressant Agent “OR “Neuroleptic Agent” OR “Catechol OMethyltransferase Inhibitor” OR “Psychotropic Agent” OR “drugs used in the treatment of addiction” OR “Anticonvulsive Agent” OR “Antiparkinson Agent” OR “Placebo” OR “Placebo Effect” OR “Placebos” OR “Placebo” OR “Naloxone” OR “Naltrexone” OR “Topiramate” OR “Acetylcysteine” OR “Tolcapone” OR “nalmefene” OR “Narcotic Antagonist” OR “Amantadine” OR “Memantine” OR “Modafinil” OR “Olanzapine” OR “Bupropion” OR “Escitalopram “OR “Serotonin Uptake Inhibitor” OR “Paroxetine” OR “Fluvoxamine” OR “Sertraline” OR “LithiumCarbonate” OR “Valproic Acid” OR “Clomipramine” OR “Tricyclic Antidepressant Agent” OR naloxone OR naltrexone OR topiramate OR n-acetylcysteine OR tolcapone OR nalmefene OR amantadine ORmemantine OR modafinil OR olanzapine OR bupropion OR escitalopram OR paroxetine ORfluvoxamine OR sertraline OR lithium OR valproate OR clomipramine) OR AK=(“Drug Therapy” OR “Drug therapy” OR “Drug therap*” OR “Pharmacological Treatments” OR “Pharmacological Treatment” OR “Pharmacologic Treatments” OR “Pharmacologic Treatment” OR “Pharmacological Therapy” OR “Pharmacologic Therapy” OR “Pharmacotherapy” OR “Pharmacotherap*” OR “Psychopharmacotherapy” OR “Psychopharmacotherap*” OR “Drug treatment” OR “Drug treat*” OR “Medication” OR “Medications” OR “Medicat*” OR “pharmaco*” OR “Narcotic Antagonist” OR quot;dopamine receptor stimulating agent” OR “Hypnotic Sedative Agent” OR “Antidepressant Agent” OR “Neuroleptic Agent” OR “Catechol OMethyltransferase Inhibitor” OR “Psychotropic Agent” OR “drugs used in the treatment of addiction” OR “Anticonvulsive Agent” OR “Antiparkinson Agent” OR “Naloxone” OR “Naltrexone” OR “Topiramate” OR “Acetylcysteine” OR “Tolcapone” OR “nalmefene” OR “Narcotic Antagonist” OR “Amantadine” OR “Memantine” OR “Modafinil” OR “Olanzapine” OR “Bupropion” OR “Escitalopram” OR “Serotonin Uptake Inhibitor” OR “Paroxetine” OR “Fluvoxamine” OR “Sertraline” OR “Lithium Carbonate” OR “Valproic Acid” OR “Clomipramine” OR “Tricyclic Antidepressant Agent” OR naloxone OR naltrexone OR topiramate OR n-acetylcysteine OR tolcapone OR nalmefene OR amantadine OR memantine OR modafinil OR olanzapine OR bupropion OR escitalopram OR paroxetine OR fluvoxamine OR sertraline OR lithium OR valproate OR clomipramine OR “Placebo” OR “Placebo Effect” OR “Placebos” OR “Placebo “) OR AB=(“Drug Therapy” OR “Drug therapy” OR “Drug therap*” OR “Pharmacological Treatments” OR “Pharmacological Treatment” OR “Pharmacologic Treatments” OR “Pharmacologic Treatment” OR “Pharmacological Therapy” OR “Pharmacologic Therapy” OR “Pharmacotherapy” OR “Pharmacotherap*” OR “Psychopharmacotherapy” OR “Psychopharmacotherap*” OR “Drug treatment” OR “Drug treat*” OR “Medication” OR “Medications” OR “Medicat*” OR “pharmaco*” OR “Narcotic Antagonist” OR “dopamine receptor stimulating agent” OR “Hypnotic Sedative Agent” OR “Antidepressant Agent” OR “Neuroleptic Agent” OR “Catechol OMethyltransferase Inhibitor” OR “Psychotropic Agent” OR “drugs used in the treatment of addiction” OR “Anticonvulsive Agent” OR “Antiparkinson Agent” OR “Naloxone” OR “Naltrexone” OR “Topiramate” OR “Acetylcysteine” OR “Tolcapone” OR “nalmefene” OR “Narcotic Antagonist” OR “Amantadine” OR “Memantine” OR “Modafinil” OR “Olanzapine” OR “Bupropion” OR “Escitalopram” OR “Serotonin Uptake Inhibitor” OR “Paroxetine” OR “Fluvoxamine” OR “Sertraline” OR “Lithium Carbonate” OR “Valproic Acid” OR “Clomipramine” OR “Tricyclic Antidepressant Agent” OR naloxone OR naltrexone OR topiramate OR n-acetylcysteine OR tolcapone OR nalmefene OR amantadine OR memantine OR modafinil OR olanzapine OR bupropion OR escitalopram OR paroxetine OR fluvoxamine OR sertraline OR lithium OR valproate OR clomipramine OR “Placebo” OR “Placebo Effect” OR “Placebos” OR “Placebo “)) AND (TI=(randomized controlled trial OR controlled clinical trial OR randomized OR placebo OR “drug therapy” OR randomly OR trial OR groups OR “Cross-Over Studies” OR “Crossover” OR “Cross over “) OR AK=(randomized controlled trial OR controlled clinical trial OR randomized OR placebo OR “drug therapy” OR randomly OR trial OR groups OR “Cross-Over Studies” OR “Crossover” OR “Cross over “) OR AB=(randomized controlled trial OR controlled clinical trial OR randomized OR placebo OR “drug therapy” OR randomly OR trial OR groups OR “Cross-Over Studies” OR “Crossover” OR “Cross over “)))

#### WHO International Clinical Trials Registry Platform (ICTRP), including

Australian New Zealand Clinical Trials Registry (ANZCTR) (including clinical trials from Therapeutic Goods Administration (TGA)); Brazilian Clinical Trials Registry (ReBec); Chinese Clinical Trial Register (ChiCTR); Clinical Research Information Service (CRiS), Republic of Korea ClinicalTrials.gov (including clinical trials from FDA); Clinical Trials Registry—India (CTRI); Cuban Public Registry of Clinical Trials (RPCEC); EU Clinical Trials Register (EU-CTR) (including clinical trials from the European Medicines Agency (EMA)); German Clinical Trials Register (DRKS); Iranian Registry of Clinical Trials (IRCT); ISRCTN.org (including clinical trials from controlled-trials.com, The Wellcome Trust (UK), UK trials (UK), Action Medical Research (UK), the Medicines and Healthcare products Regulatory Agency (MHRA) and National Research Register); Japan Primary Registries Network (JPRN) (including clinical trials from UMIN-CTR, JapicCTI and JMACCT); Pan African Clinical Trial Registry (PACTR); Sri Lanka Clinical Trials Registry (SLCTR); The Netherlands National Trial Register (NTR); Thai Clinical Trials Register (TCTR); The International Federation of Pharmaceutical Manufacturers and Associations (IFPMA); UK Clinical Trials Gateway; BIOSIS Previews via Web of Knowledge; SEAGLE (OpenGrey); ProQuest Theses and Dissertations; ClinicalTrials.gov; http://www.fda.gov/; http://www.ema.europa.eu

All of the above will be searched for any type of Randomized Controlled Trial. Only English language records or available English language translations thereof will be sought. Searches will span a period from inception up to search date.

### §S4 Confidence in Network Meta-Analysis (CINeMA)

#### Within study bias

Within-study bias refers to shortcomings in the design or conduct of a study that can lead to an estimated relative treatment effect that systematically differs from the truth. ^7^ In our application of CINeMA we considered the per-study contribution matrix in conjunction with RoB 2 assessments to evaluate each relative treatment effect with respect to within-study bias. In CINeMA, a “Risk of bias contributions” section provides a bar chart with each bar corresponding to an estimate of relative effect - each study is represented by a colored area and is proportional to its contribution. We chose “Majority RoB 2” (a level of concern according to the RoB 2 with the greatest total percentage contribution) for all CINeMA assessments.

#### Reporting bias

Reporting bias occurs when the results included in the systematic review are not a representative sample of the results generated by studies undertaken. ^7^ We used the Risk Of Bias due to Missing Evidence in Network meta-analysis (ROB-MEN) for all assessments, ^8^ within the ROB-MEN approach, we utilized the Outcome Reporting Bias In Trials (ORBIT) to inform our reporting bias ratings ^9^. Within ROB-MEN, we did not have enough studies (>10) per comparator to conduct graphical assessment of across study bias. To evaluate small-study effects, we run a network meta-regression model with a measure of precision (e.g. variance or standard error) as the covariate via ROB-MEN ^8^. This model generates an adjusted relative effect by extrapolating the regression line to the smallest observed variance (the ‘largest’ study) independently for each comparison. To assess the presence of small-study effects, we compared the obtained adjusted estimates with the original (unadjusted) estimates by looking at the overlap of their corresponding confidence intervals. We regarded a lack of overlap between the two intervals (or between one estimate and the interval for the other estimate) as an indication of small-study effect. ROB-MEN scores were then loaded into CINeMA.

#### Indirectness

Indirectness refers to the relevance of the included studies to the research question. After assessing the relevance of the included studies in our NMA, we set all indirectness contributions to no concern. In CINeMA, indirectness contributions (matrix) were resolved by majority voting.

#### Imprecision

CINeMA compares the treatment effects included in the 95% confidence interval with the range of equivalence.^7^ This helps to understand if the treatment effect extends beyond the area of equivalence and assign the respective confidence ratings. To evaluate imprecision the relative treatment effect that represents a clinically important difference needs to be defined. We defined a clinically important size of effect for continuous outcomes (gambling severity and quality of life) as (−0.20, 0.20) and for dichotomous outcomes (tolerability) as (0.80, 1.25).

#### Heterogeneity

Variability in the results of studies influences our confidence in the point estimate of a relative treatment effect. If this variability reflects genuine differences between studies, rather than random variation, it is called heterogeneity. ^7^ The CINeMA approach to heterogeneity involves comparisons of results with the pre-specified range of clinical equivalence, therefore, the definition of clinically important difference (see above, imprecision) was used here as well. This evaluates the prediction intervals (which capture heterogeneity) in addition to confidence intervals in relation to the range of equivalence. Confidence ratings for heterogeneity followed the rules outlined in Nikolakopoulou et al. ^7^.

#### Incoherence

In network meta-analysis, there may be variation between direct and indirect sources of evidence. This is called incoherence. ^7^ Incoherence testing requires a definition of clinically important difference, and here again we used the definition above (see imprecision). In quality of life NMA, incoherence testing was not statistically possible due to absence of closed loops. In this case, all ratings were set to “some concern”.

#### Overall Confidence rating

We set the overall confidence ratings according to the ruleset below:

We added the 6 domains to achieve a total (overall) confidence score. Each domain contributed by simple addition according to this rule:

Low risk/No concerns = 0; Some concerns = 1; Major Concerns = 3

Overall score: 0-2 = High; 3-4 = Moderate; 5-6 = Low; 7+ = Very Low

We used the semi-automated methods of the contribution matrix to pool results in our sample from https://cinema.ispm.unibe.ch/ ^10^.

### §S5 Characteristics of studies included in the review

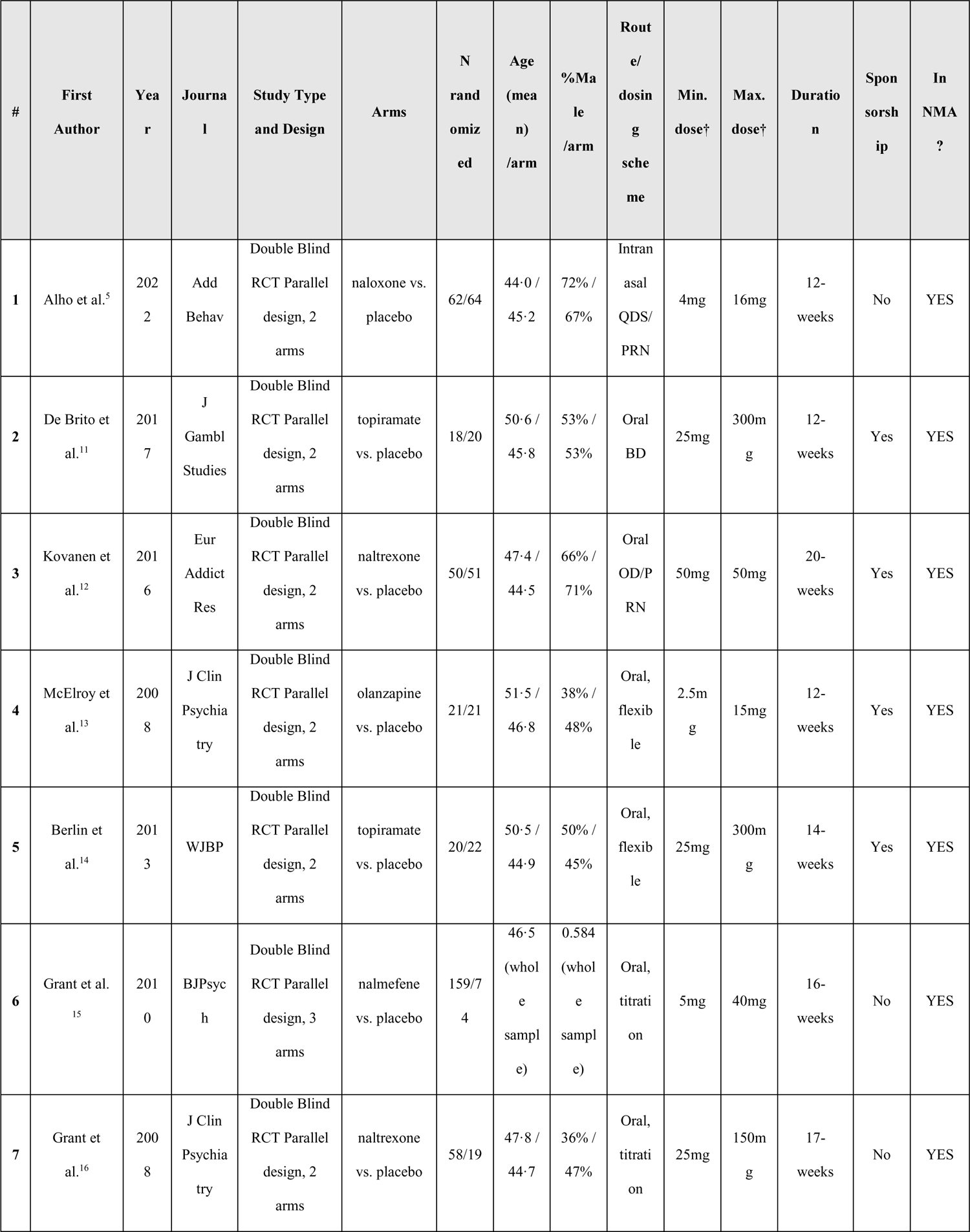
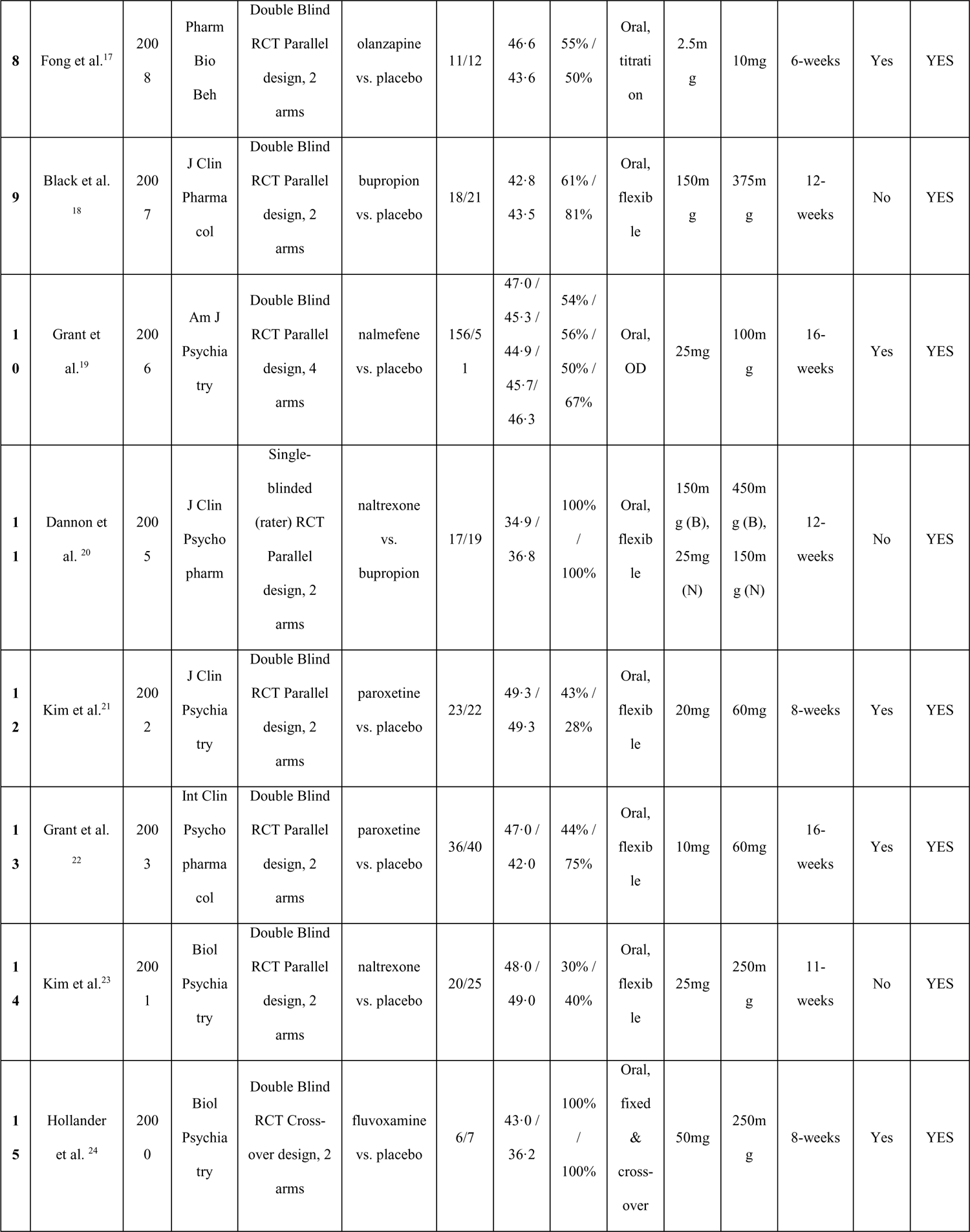
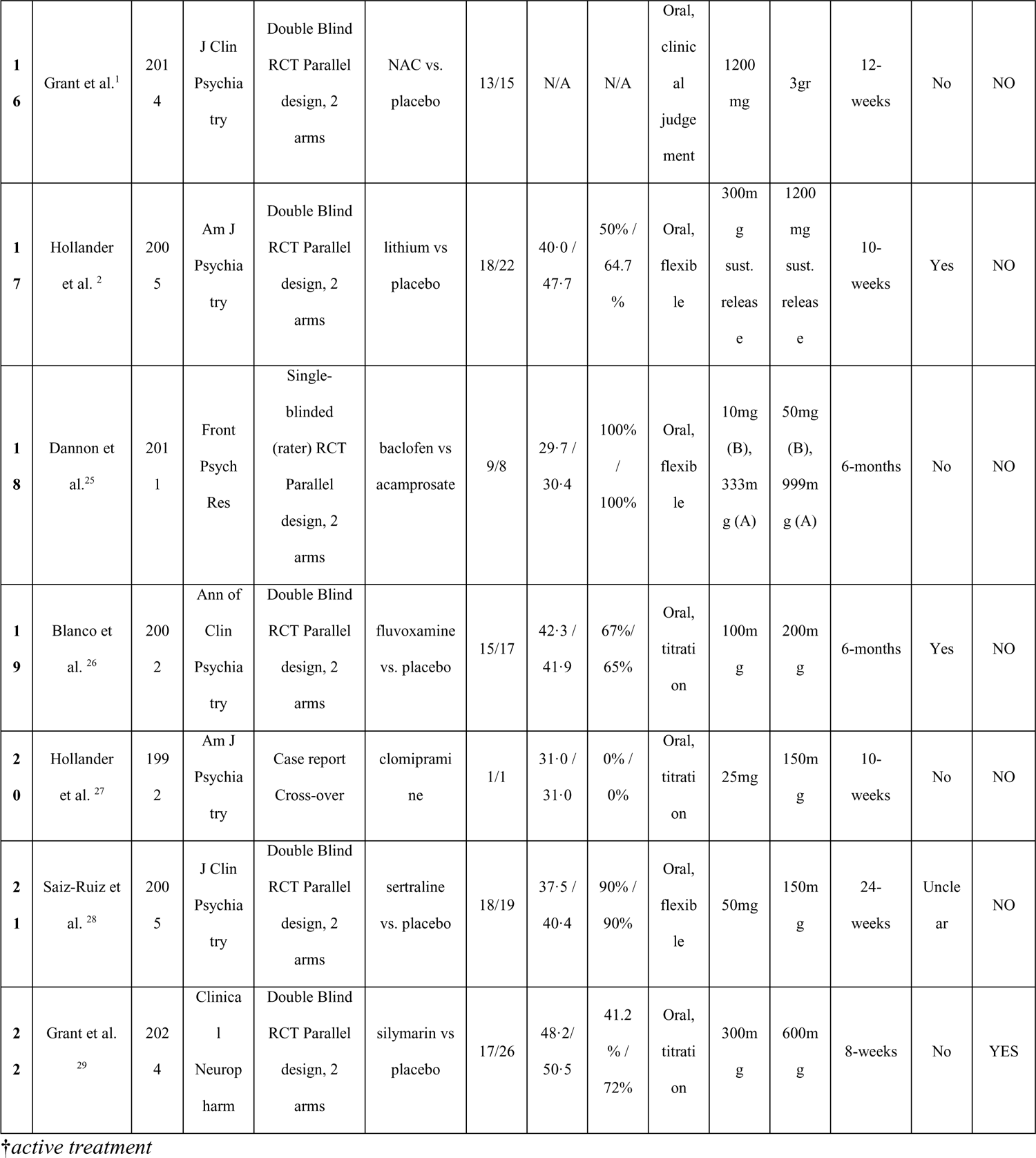

#### Risk of Bias 2 assignments

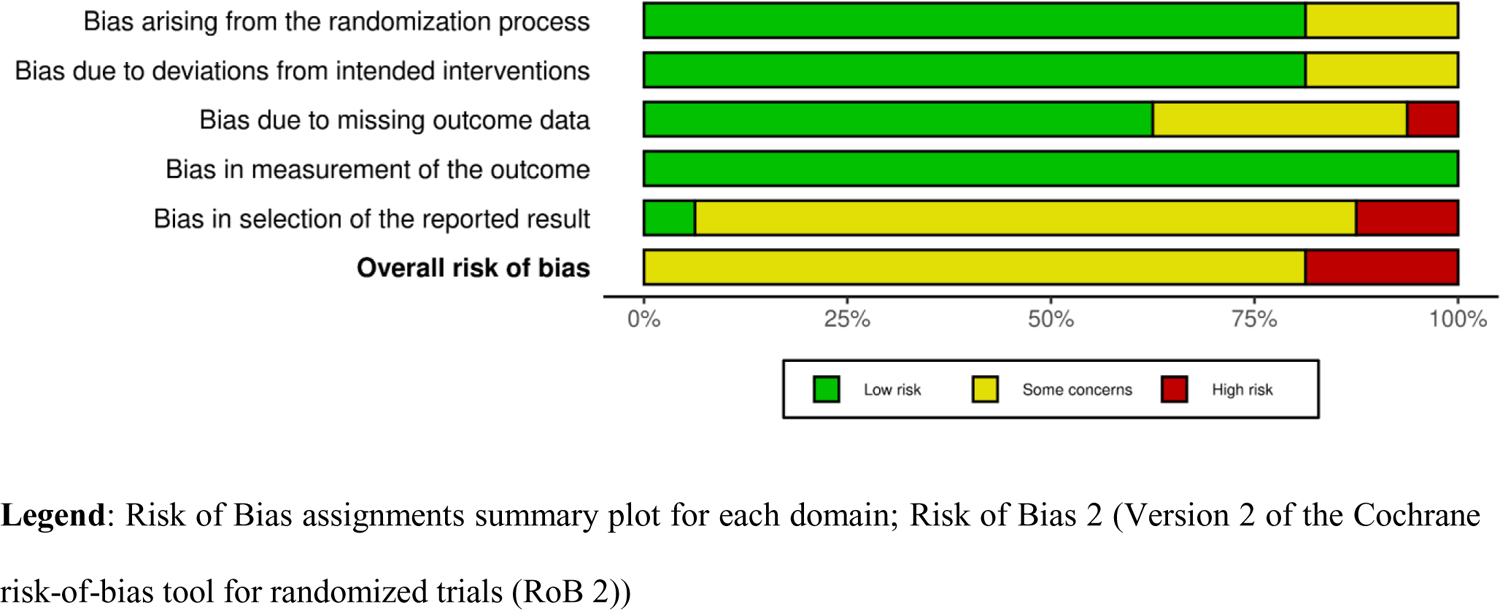

For within-study bias, each individual RCT included in our Network Meta-Analysis was assessed using the Cochrane Risk of Bias tool version 2 (RoB-2), as recommended in The Cochrane Handbook of Systematic Reviews of Interventions.^30^ The tool includes five domains on different sources of bias. For RCTs (including cross-over trials), these include: 1) bias arising from the randomisation process; 2) bias due to deviations from intended interventions; 3) bias due to missing outcome data; 4) bias in measurement of the outcome; and 5) bias in selection of the reported result.

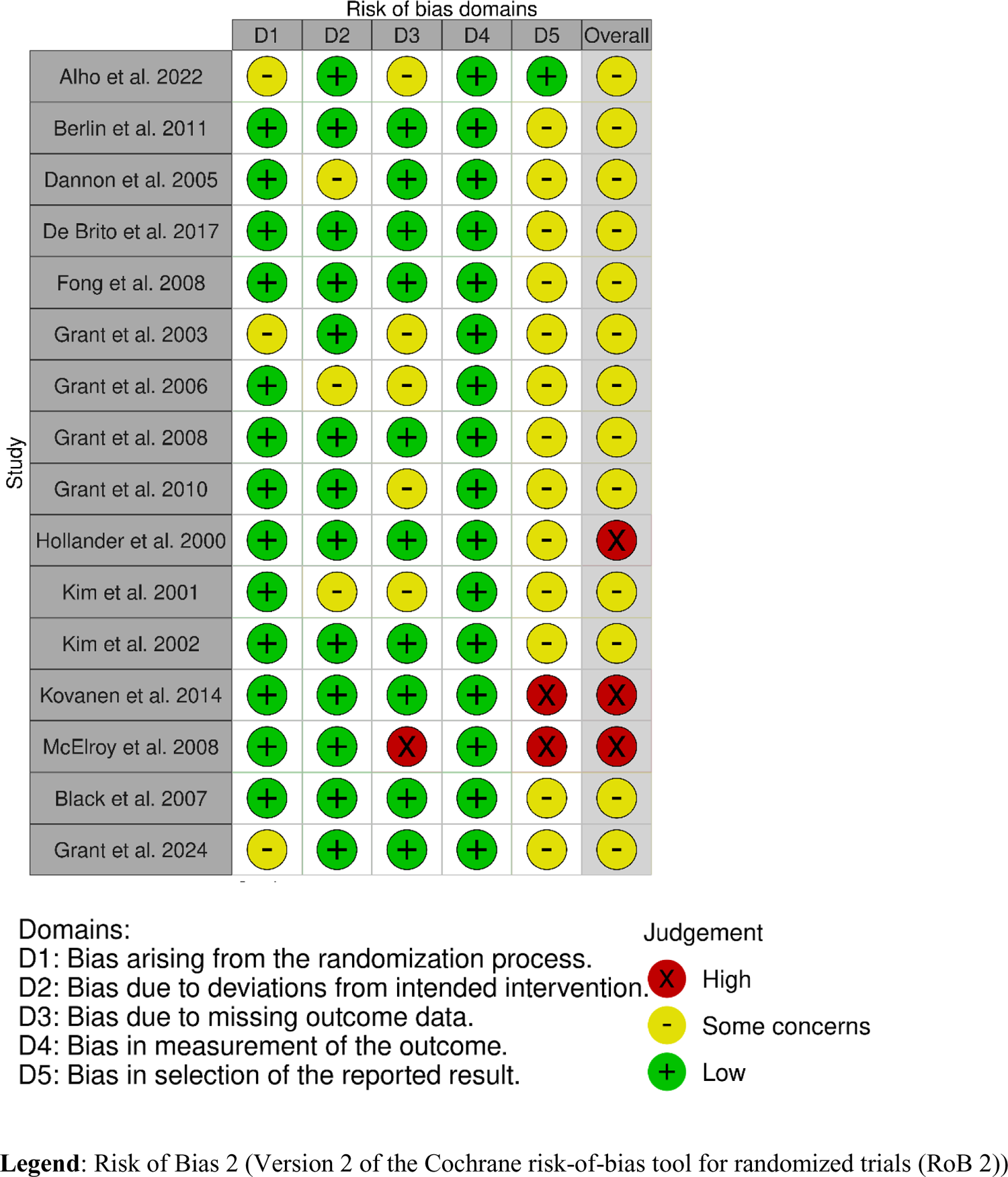

#### Outcome Reporting Bias in Trials (ORBIT) classifications

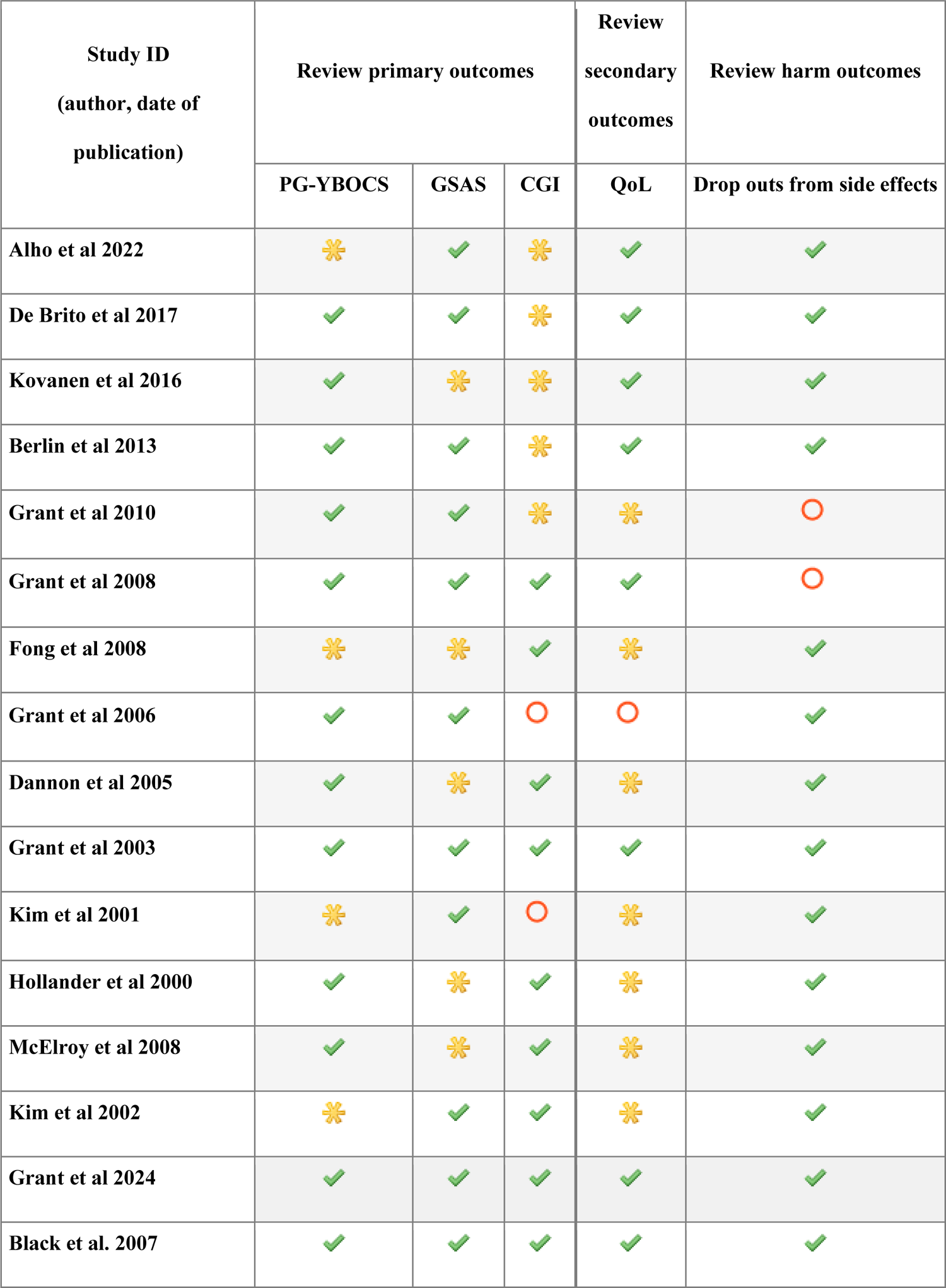
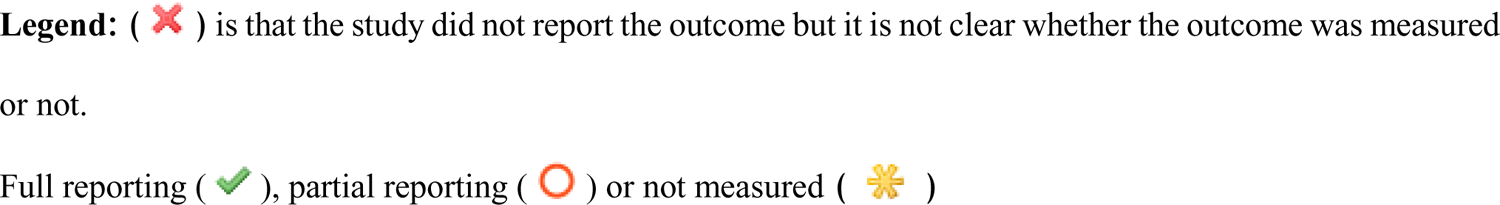

### §S6 Pairwise meta-analysis results

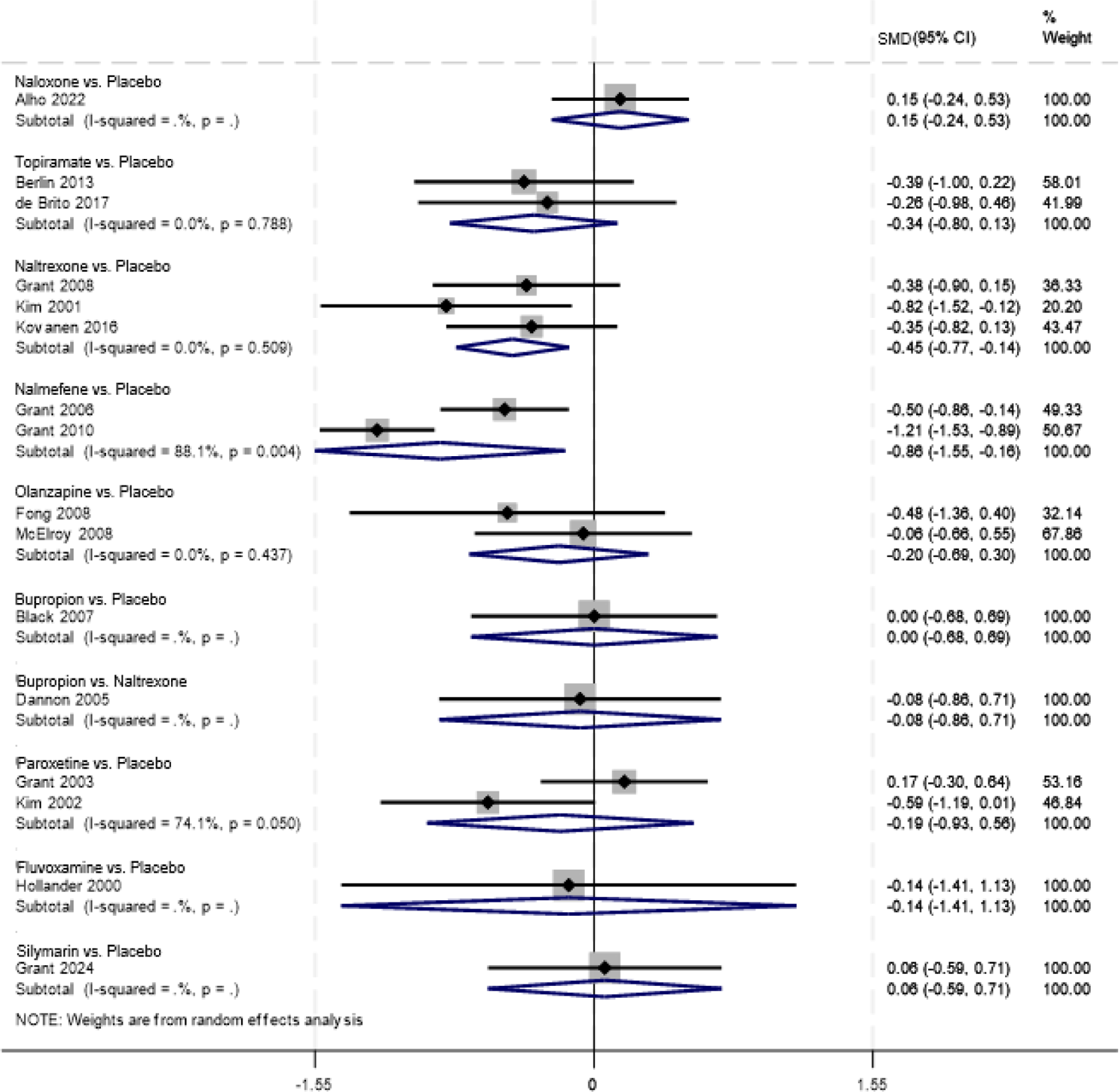

#### Gambling symptom severity

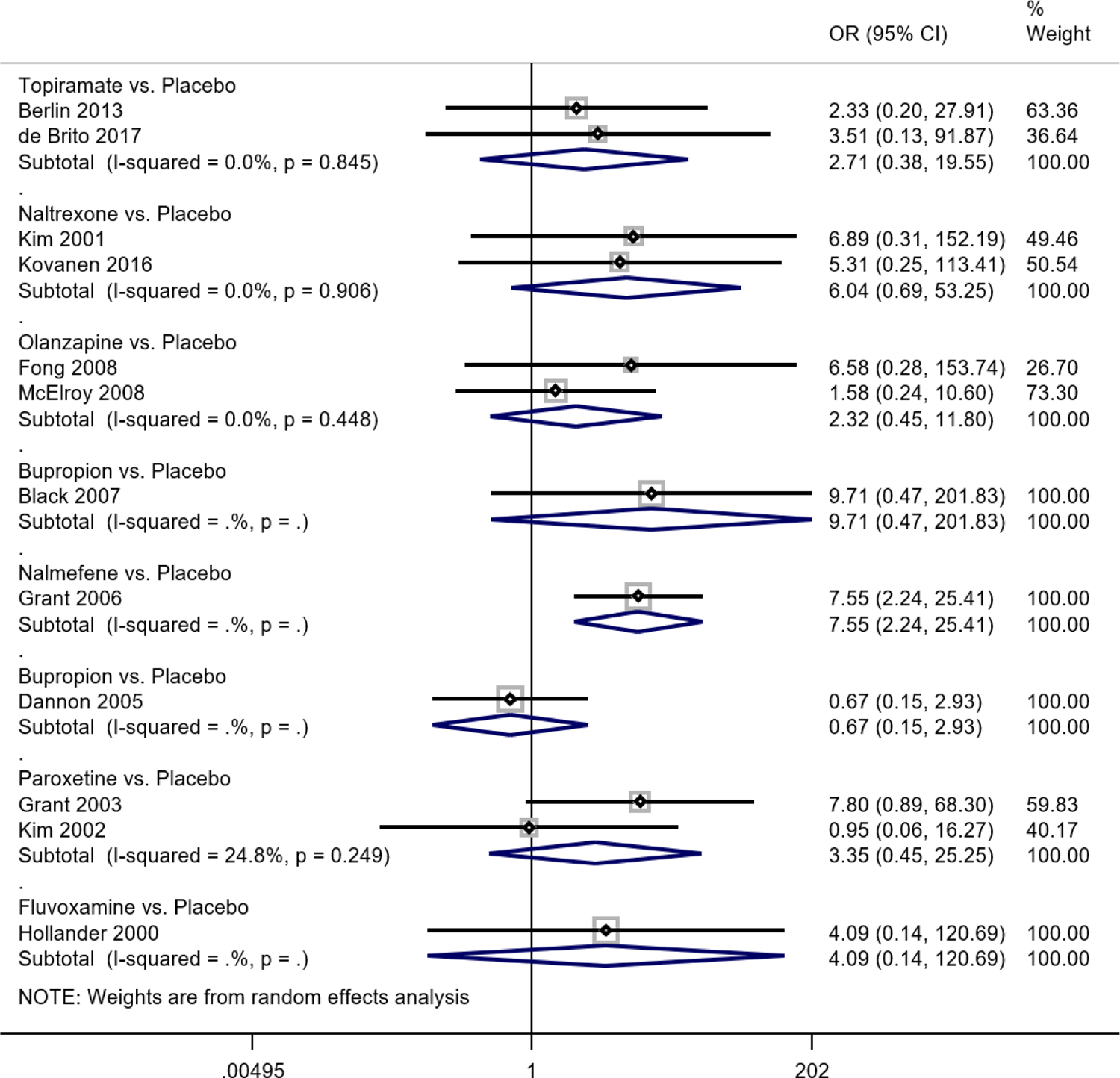

#### Tolerability

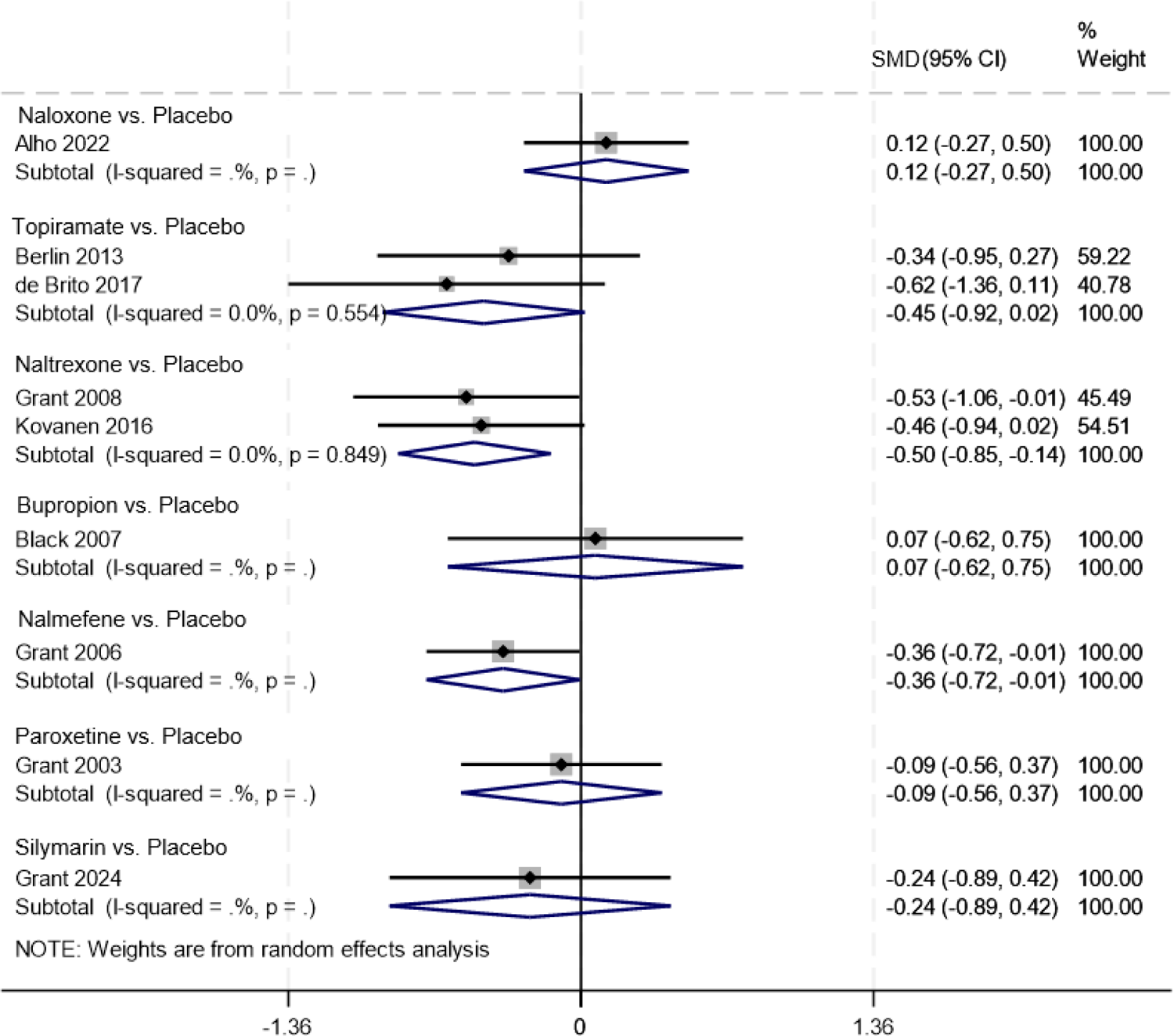

#### Quality of life

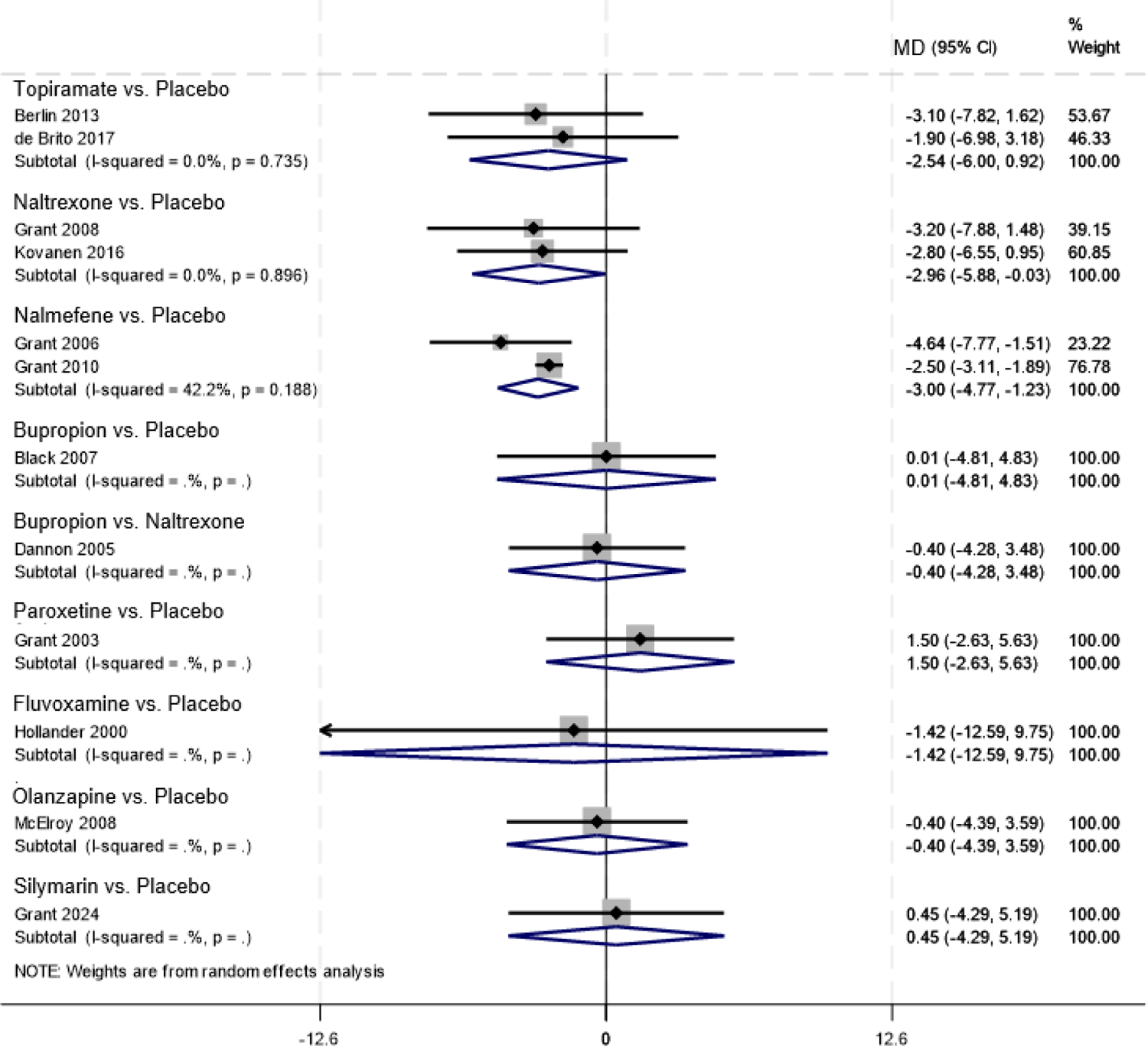

#### PG-YBOCS

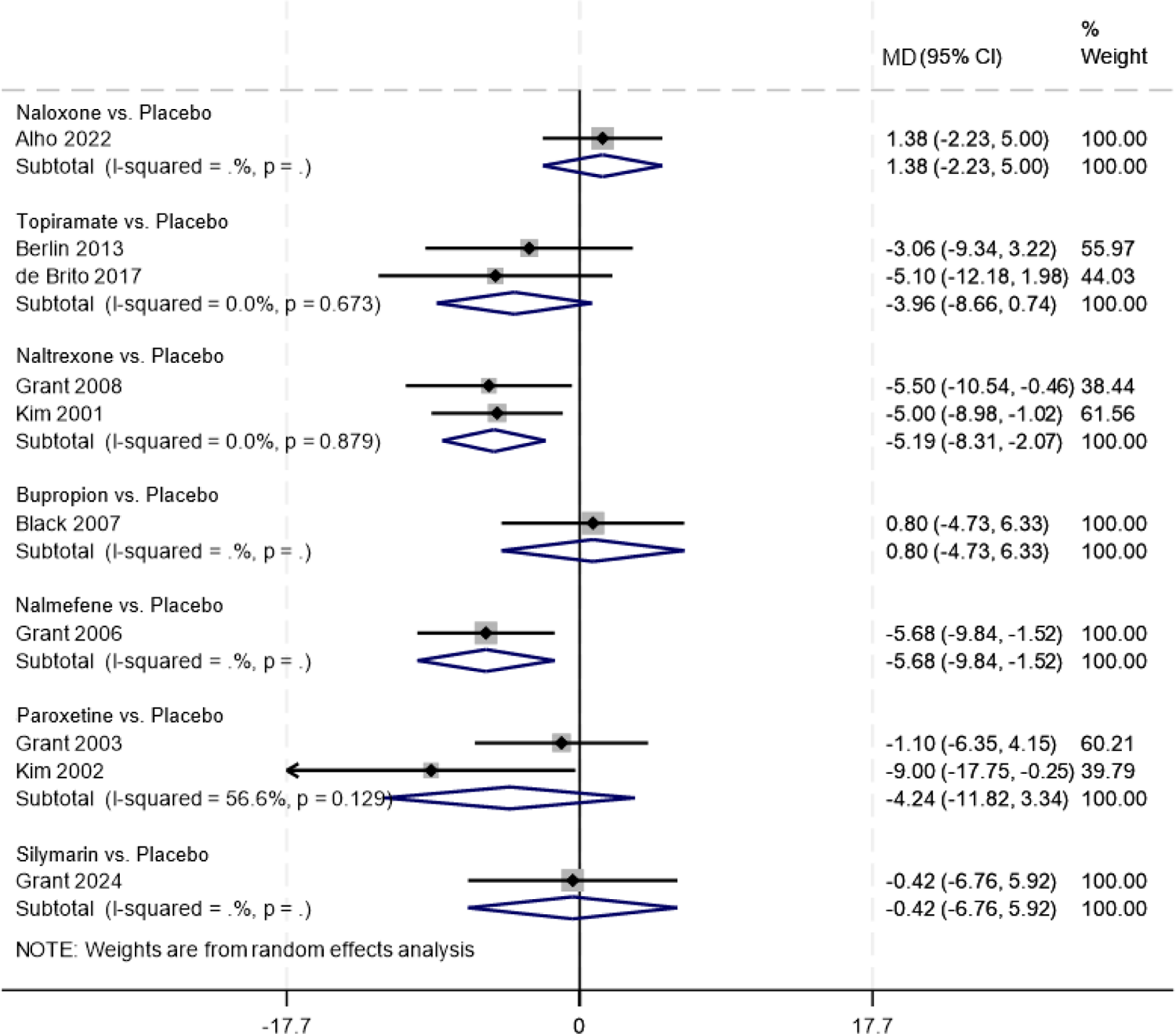

#### GSAS only (gambling severity secondary analysis)

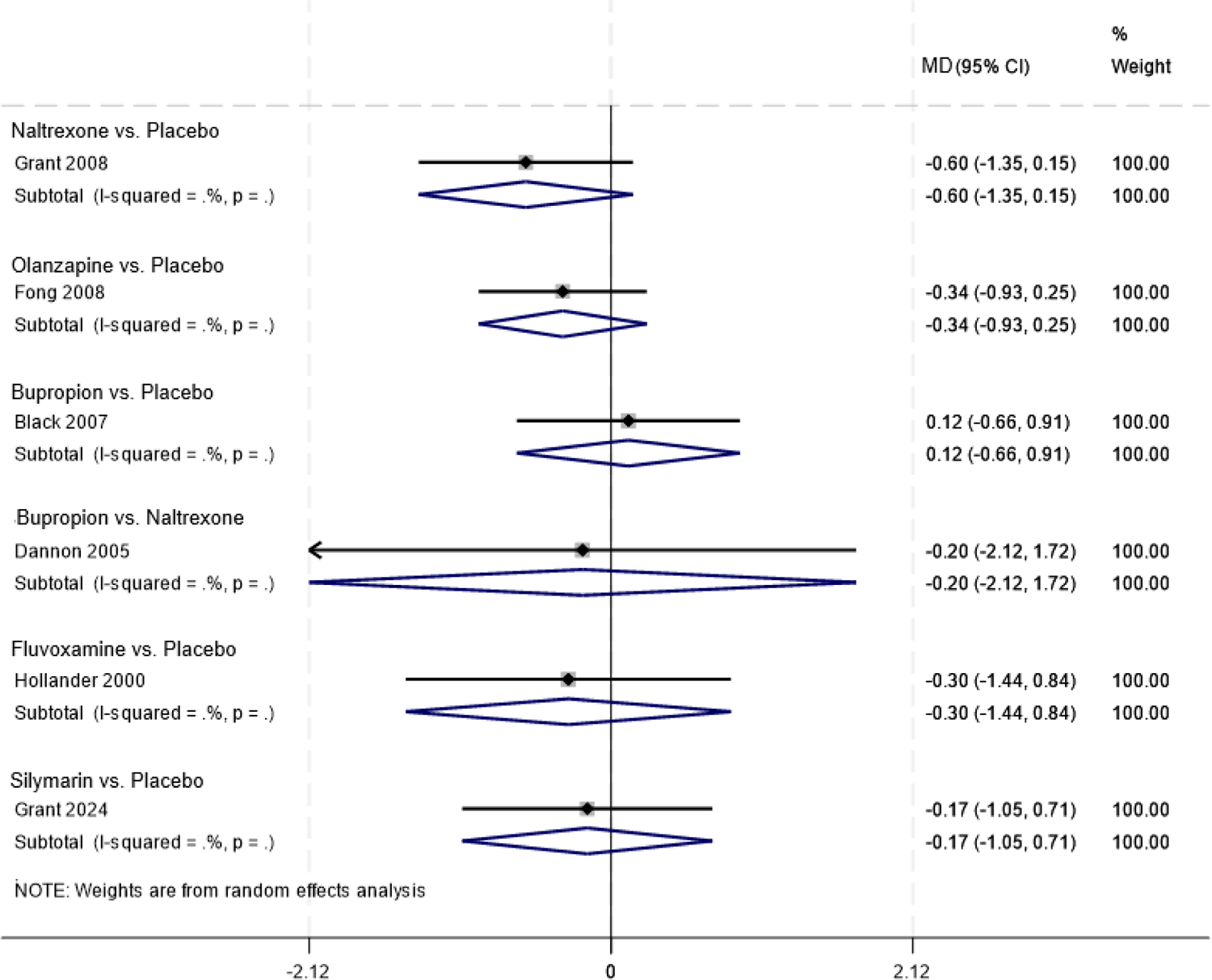

#### CGI-I

| Main Treatment effect |  |  |  | Tolerability |  |  |  | Quality of life |  |  |  |
| --- | --- | --- | --- | --- | --- | --- | --- | --- | --- | --- | --- |
| Treatment | SUCRA | PrBest | MeanRank | Treatment | SUCRA | PrBest | MeanRank | Treatment | SUCRA | PrBest | MeanRank |
| Nalmefene | 94.3 | 66.0 | 1.5 | Placebo | 90.9 | 50.4 | 1.6 | Naltrexone | 83.7 | 36.8 | 2.1 |
| Naltrexone | 69.2 | 4.1 | 3.8 | Olanzapine | 63.4 | 11.2 | 3.6 | Topiramate | 77.9 | 30.2 | 2.5 |
| Topiramate | 60.5 | 5.6 | 4.6 | Topiramate | 57.3 | 12.0 | 4.0 | Nalmefene | 70.4 | 12.8 | 3.1 |
| Bupropion | 49.7 | 3.6 | 5.5 | Paroxetine | 50.7 | 5.3 | 4.4 | Silymarin | 55.1 | 15.2 | 4.1 |
| Olanzapine | 51.2 | 3.2 | 5.4 | Bupropion | 36.7 | 2.5 | 5.4 | Paroxetine | 40.3 | 2.2 | 5.2 |
| Fluvoxamine | 46.0 | 14.6 | 5.9 | Fluvoxamine | 47.3 | 18.1 | 4.7 | Placebo | 28.3 | 0.0 | 6.0 |
| Paroxetine | 46.1 | 1.0 | 5.8 | Nalmefene | 27.1 | 0.0 | 6.1 | Bupropion | 26.9 | 2.8 | 6.1 |
| Silymarin | 31.3 | 1.8 | 7.2 | Naltrexone | 26.5 | 0.5 | 6.1 | Naloxone | 17.4 | 0.1 | 6.8 |
| Placebo | 28.8 | 0.0 | 7.4 | Naloxone | NA | NA | NA | Olanzapine | NA | NA | NA |
| Naloxone | 22.9 | 0.2 | 7.9 | Silymarin | NA | NA | NA | Fluvoxamine | NA | NA | NA |

**Legend:** Treatment rankings from the main network meta-analysis. SUCRA = surface under the cumulative ranking curve; PrBest = Probability of treatment to produce the best effect out of all comparators (Note: PrBest is biased towards treatments with high uncertainty of evidence); Mean Rank = Probabilistic ranking metric, i.e. the probability that a treatment achieves a specific rank.<sup>32</sup>

### §S7 Results from NMA for each gambling severity scale

#### PG-YBOCS (gambling severity sensitivity analysis)

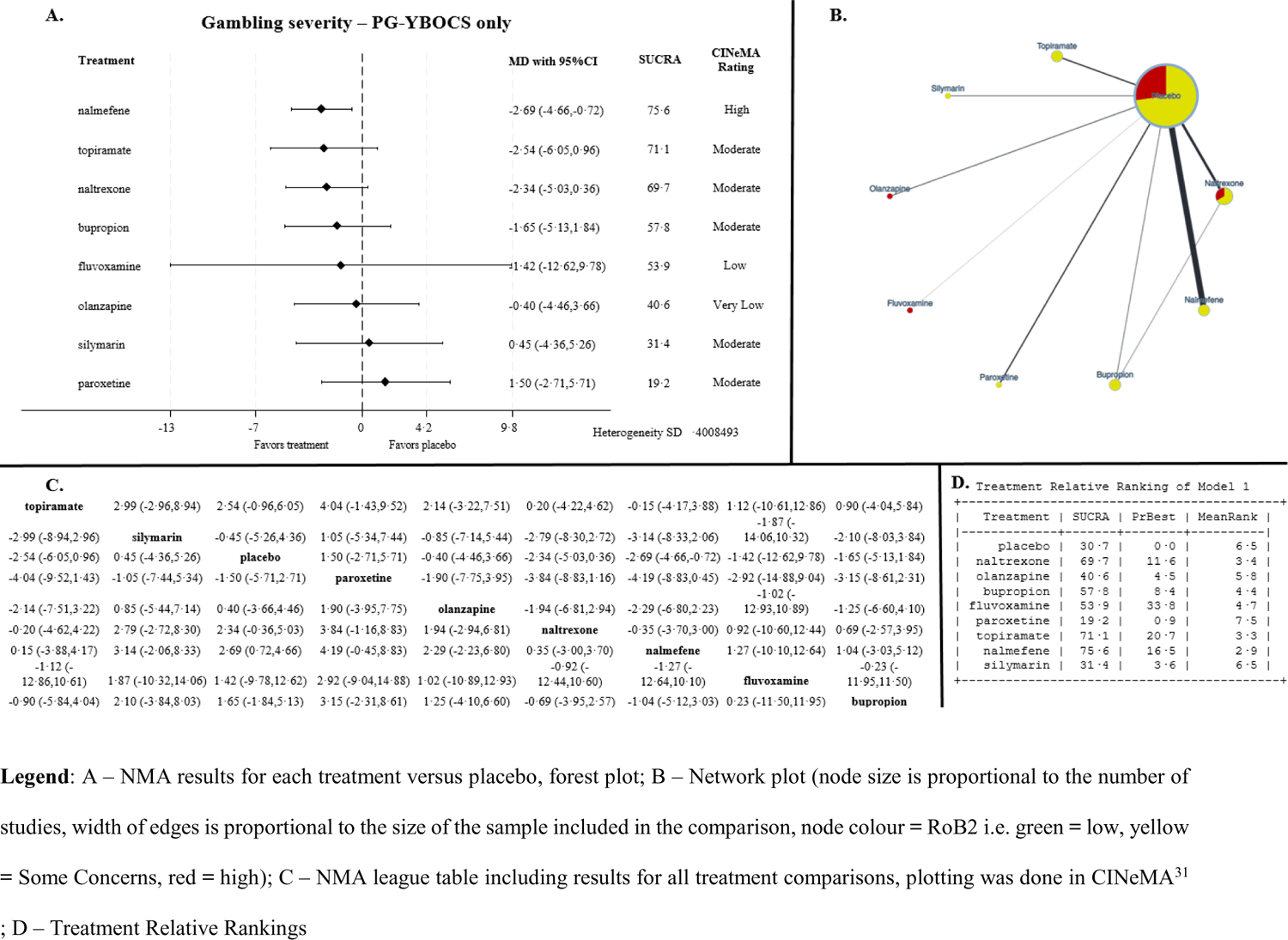

#### GSAS only (gambling severity sensitivity analysis)

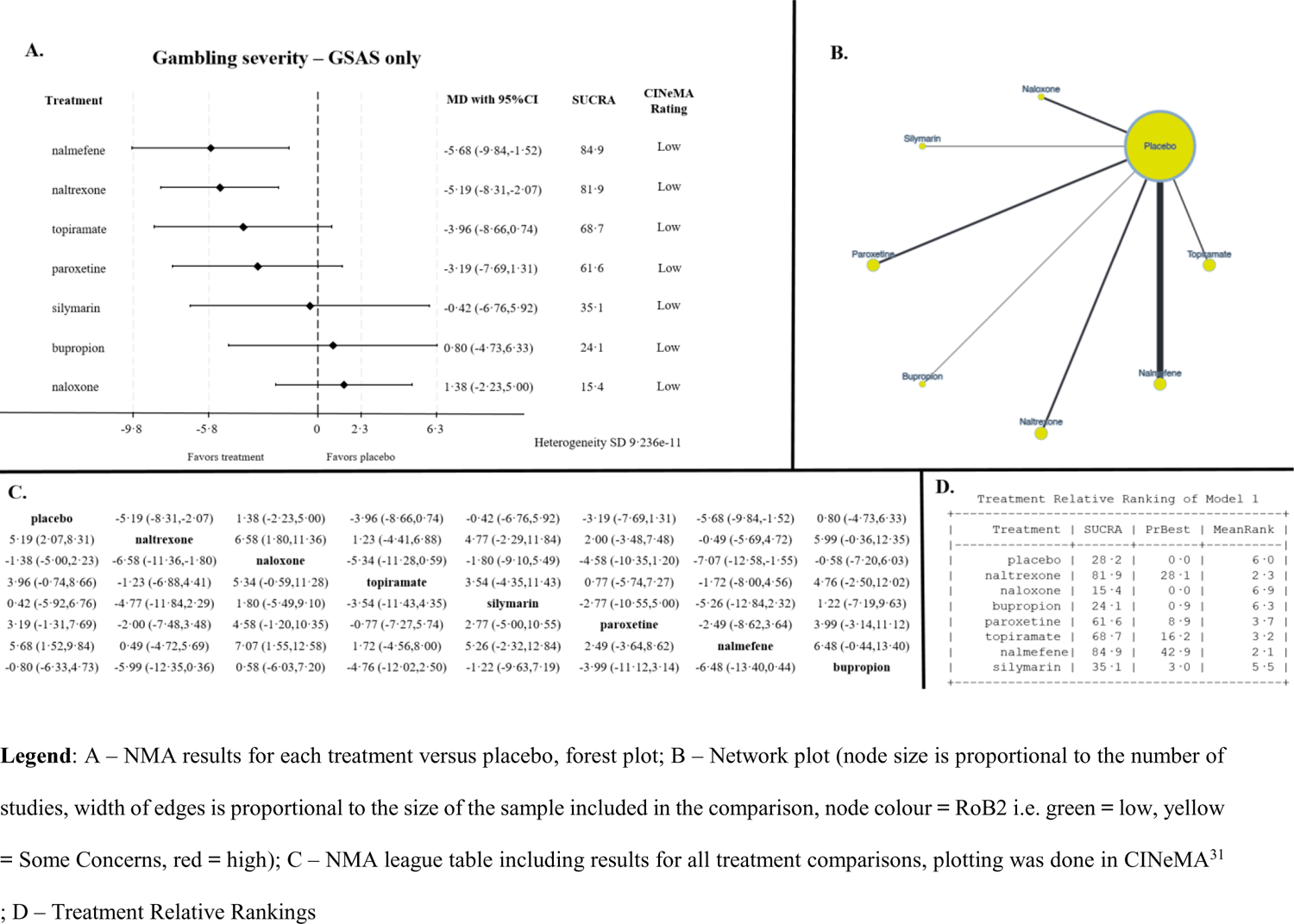

#### CGI-I only (gambling severity sensitivity analysis)

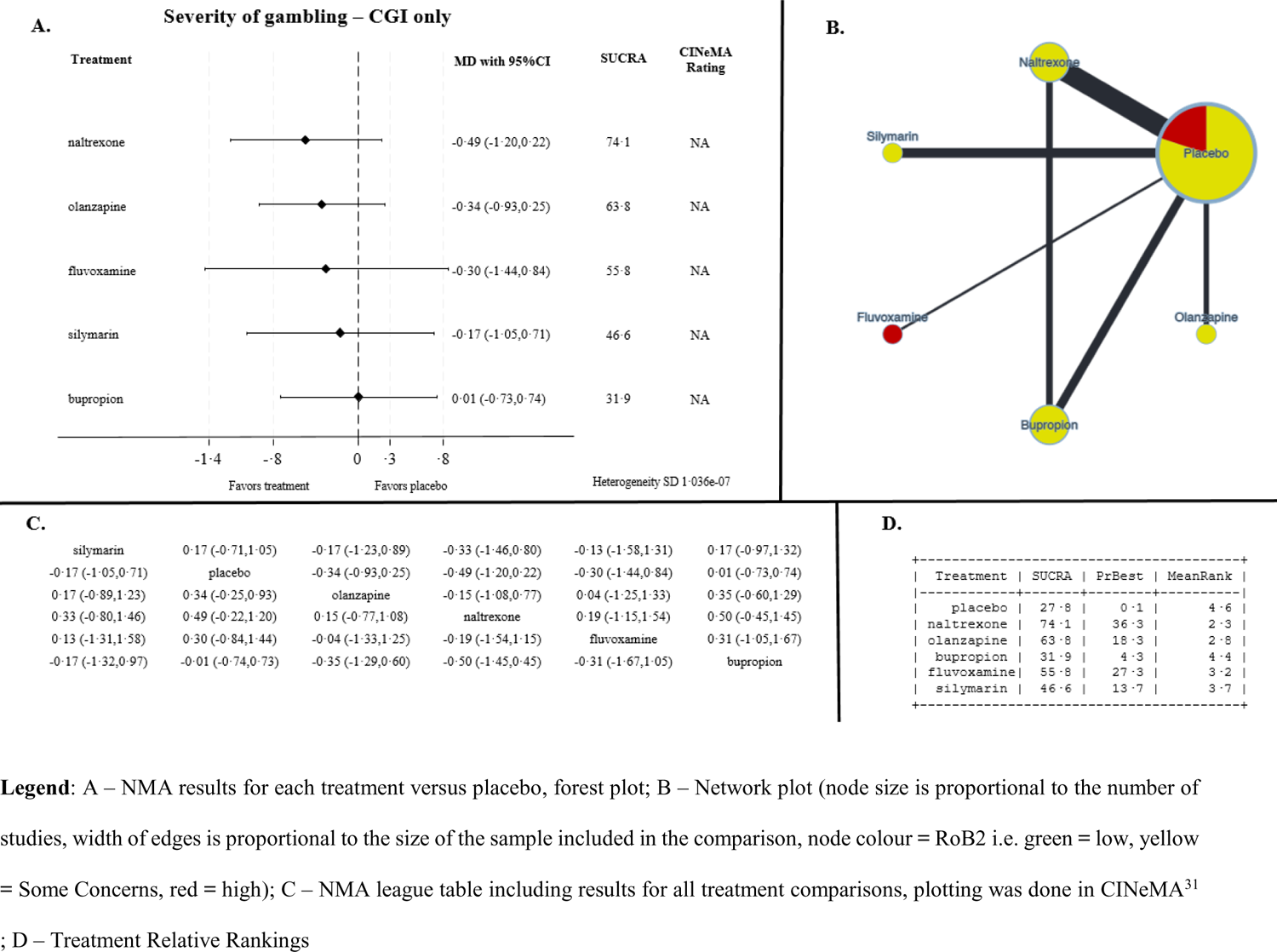

## §S8 Heterogeneity measures within NMA and results from incoherence assessment for all outcomes

### Gambling symptom severity

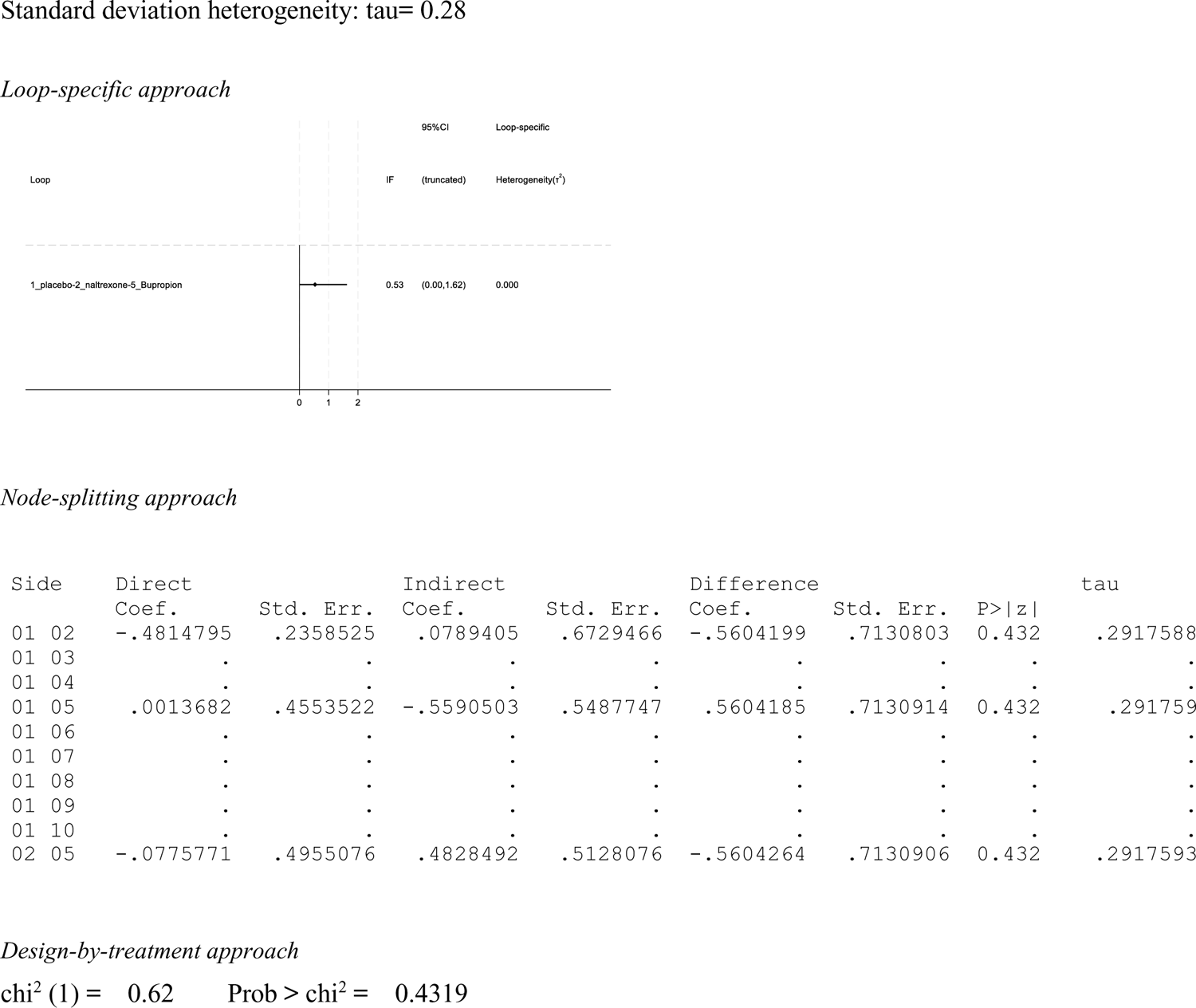

### Tolerability

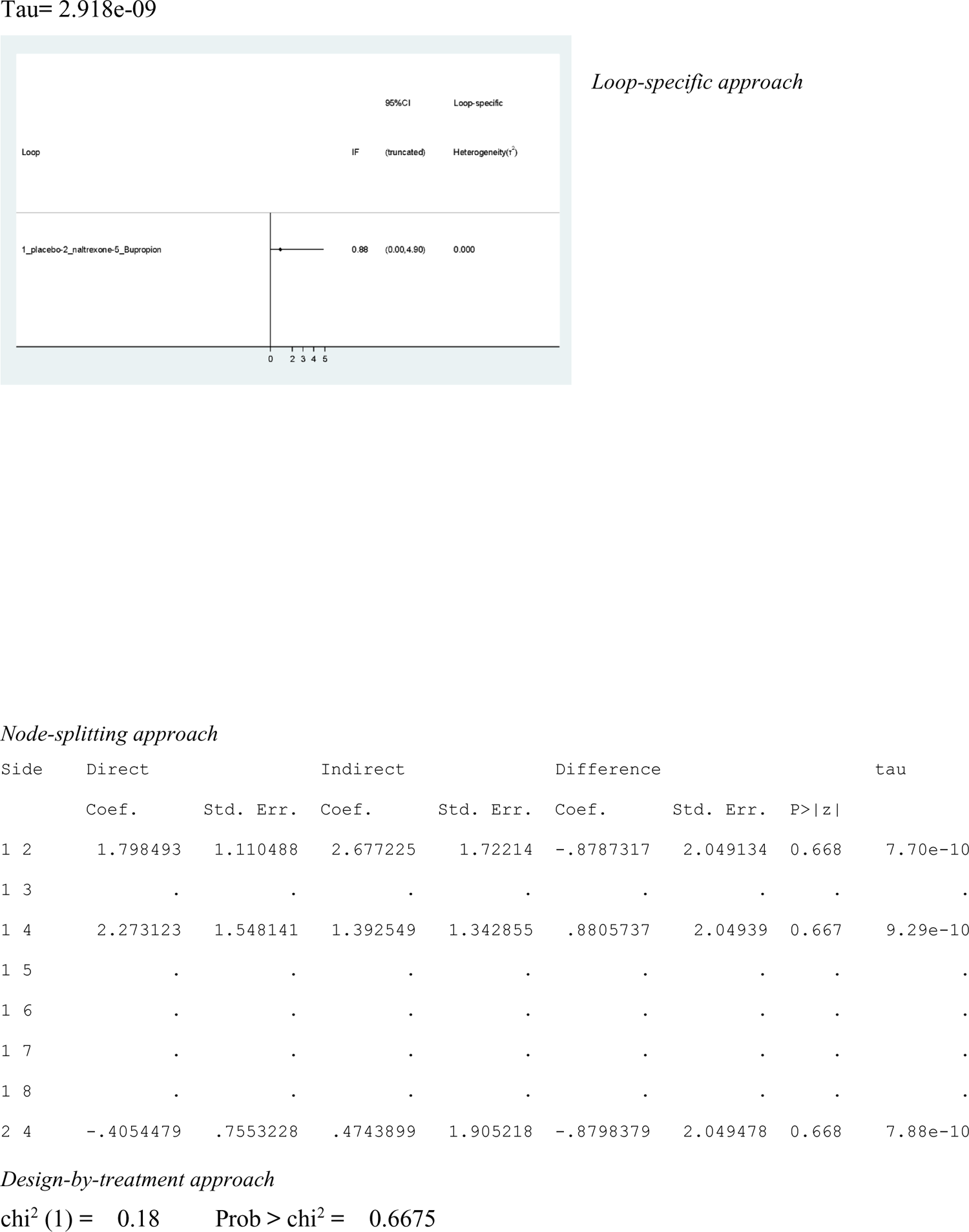

### Standard deviation heterogeneity

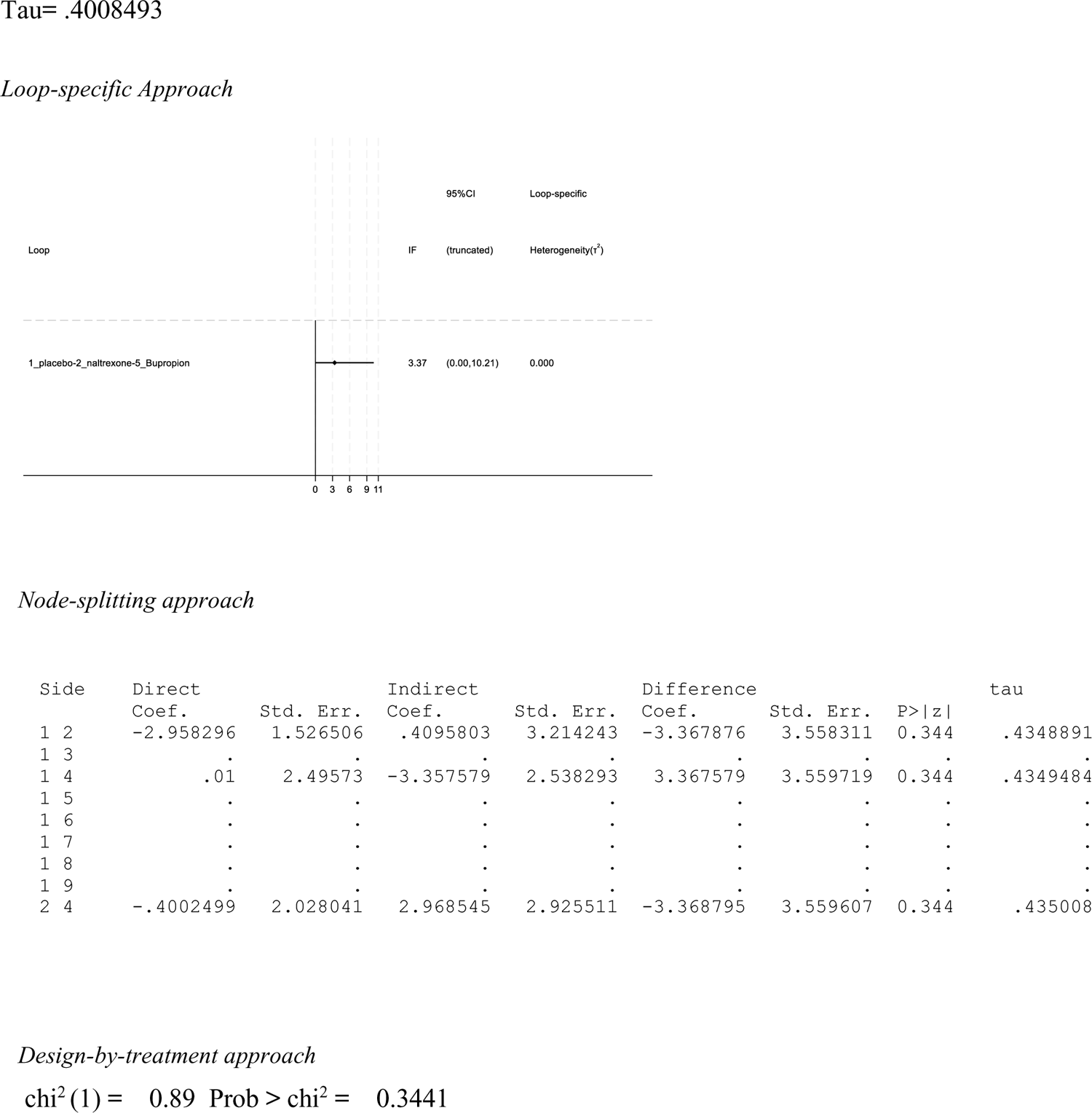

### PG-YBOCS only (gambling severity sensitivity analysis)

#### Standard deviation heterogeneity

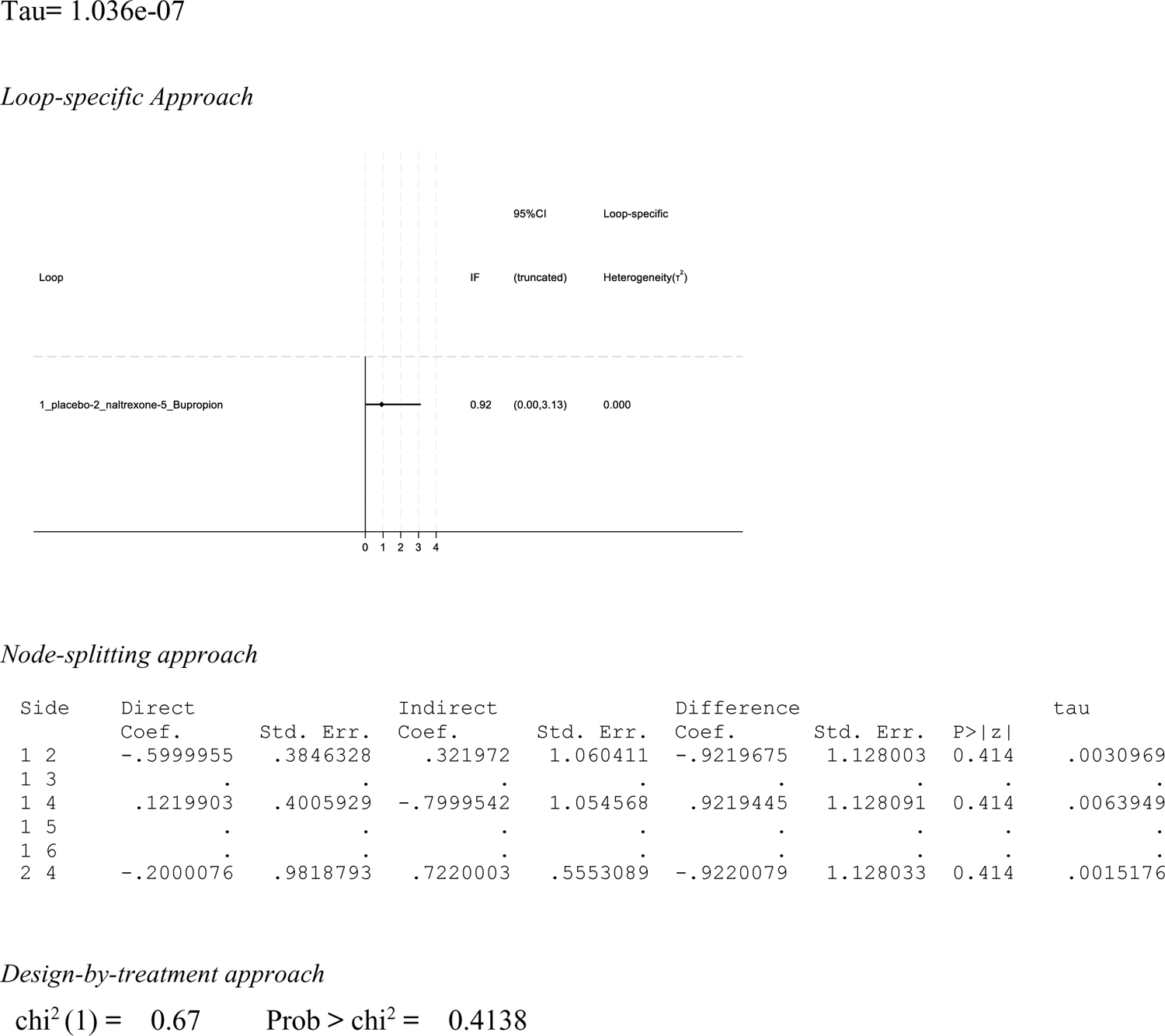

CGI-I only (gambling severity sensitivity analysis)

Standard deviation heterogeneity

### §S9 Treatment rankings from main NMA, treatment effect, tolerability and quality of life

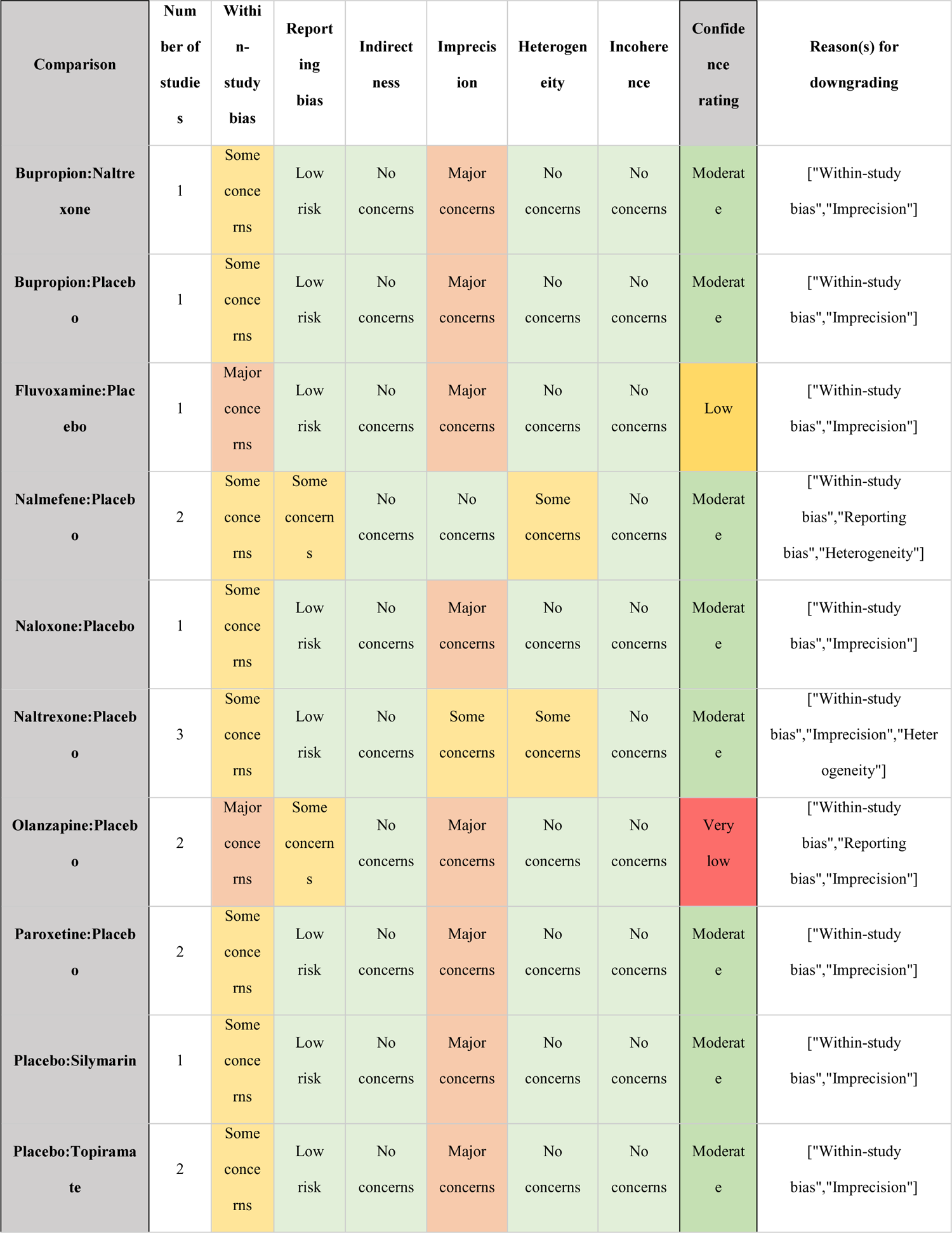
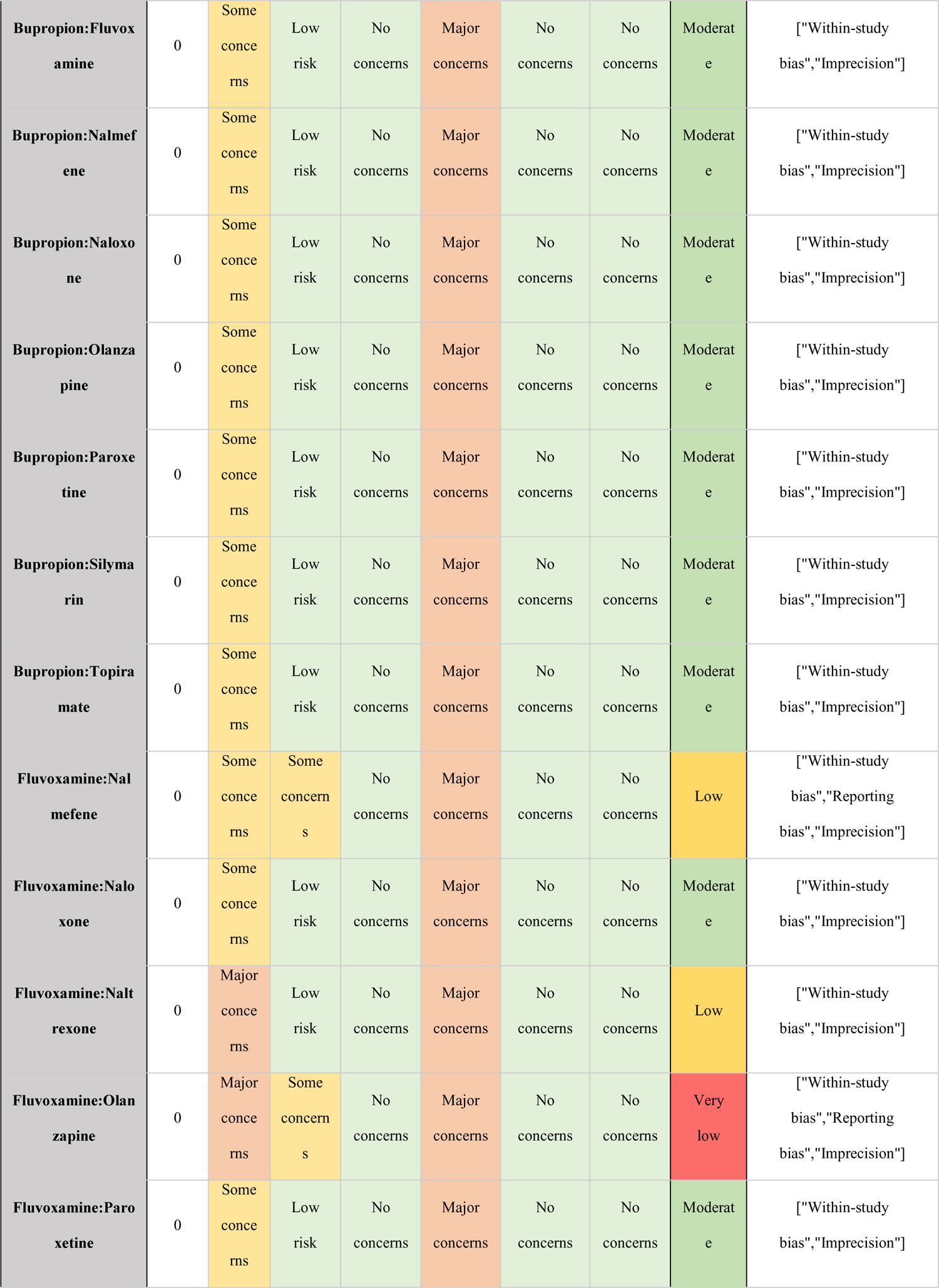
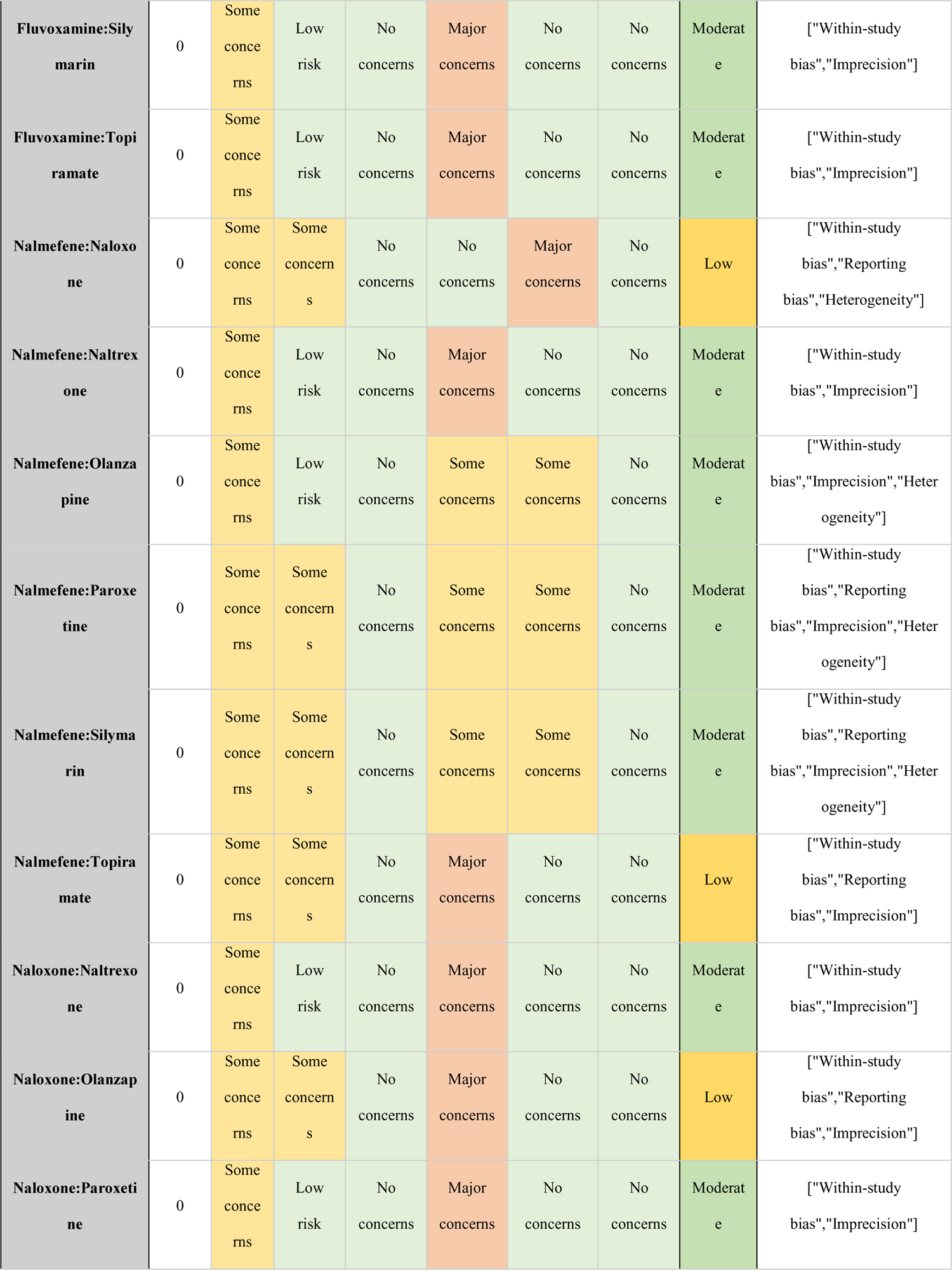
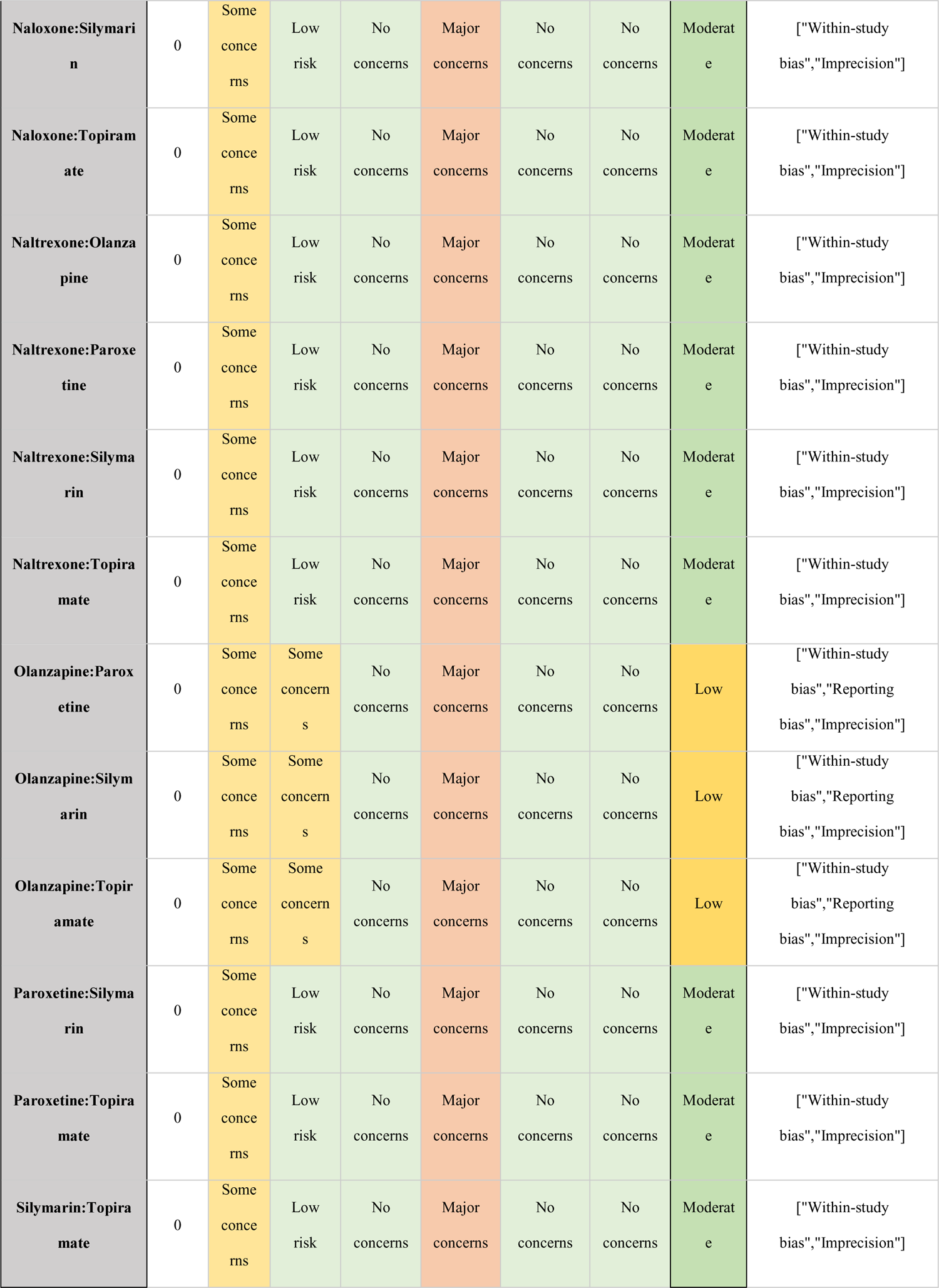

| Comparison | Number of studies | Within-study bias | Reporting bias | Indirectness | Imprecision | Heterogeneity | Incoherence | Confidence rating | Reason(s) for downgrading |
| --- | --- | --- | --- | --- | --- | --- | --- | --- | --- |
| Bupropion:Naltrexone | 1 | Some concerns | Low risk | No concerns | Major concerns | No concerns | No concerns | Moderate | ["Within-study bias", "Imprecision"] |
| Bupropion:Placebo | 1 | Some concerns | Low risk | No concerns | Major concerns | No concerns | No concerns | Moderate | ["Within-study bias", "Imprecision"] |
| Fluvoxamine:Placebo | 1 | Major concerns | Low risk | No concerns | Major concerns | No concerns | No concerns | Low | ["Within-study bias", "Imprecision"] |
| Nalmefene:Placebo | 2 | Some concerns | Some concerns | No concerns | No concerns | Some concerns | No concerns | Moderate | ["Within-study bias", "Reporting bias", "Heterogeneity"] |
| Naloxone:Placebo | 1 | Some concerns | Low risk | No concerns | Major concerns | No concerns | No concerns | Moderate | ["Within-study bias", "Imprecision"] |
| Naltrexone:Placebo | 3 | Some concerns | Low risk | No concerns | Some concerns | Some concerns | No concerns | Moderate | ["Within-study bias", "Imprecision", "Heterogeneity"] |
| Olanzapine:Placebo | 2 | Major concerns | Some concerns | No concerns | Major concerns | No concerns | No concerns | Very low | ["Within-study bias", "Reporting bias", "Imprecision"] |
| Paroxetine:Placebo | 2 | Some concerns | Low risk | No concerns | Major concerns | No concerns | No concerns | Moderate | ["Within-study bias", "Imprecision"] |
| Placebo:Silymarin | 1 | Some concerns | Low risk | No concerns | Major concerns | No concerns | No concerns | Moderate | ["Within-study bias", "Imprecision"] |
| Placebo:Topiramate | 2 | Some concerns | Low risk | No concerns | Major concerns | No concerns | No concerns | Moderate | ["Within-study bias", "Imprecision"] |
| **Bupropion:Fluvoxamine** | 0 | Some concerns | Low risk | No concerns | Major concerns | No concerns | No concerns | Moderate | ["Within-study bias", "Imprecision"] |
| **Bupropion:Nalmefene** | 0 | Some concerns | Low risk | No concerns | Major concerns | No concerns | No concerns | Moderate | ["Within-study bias", "Imprecision"] |
| **Bupropion:Naloxone** | 0 | Some concerns | Low risk | No concerns | Major concerns | No concerns | No concerns | Moderate | ["Within-study bias", "Imprecision"] |
| **Bupropion:Olanzapine** | 0 | Some concerns | Low risk | No concerns | Major concerns | No concerns | No concerns | Moderate | ["Within-study bias", "Imprecision"] |
| **Bupropion:Paroxetine** | 0 | Some concerns | Low risk | No concerns | Major concerns | No concerns | No concerns | Moderate | ["Within-study bias", "Imprecision"] |
| **Bupropion:Silymarin** | 0 | Some concerns | Low risk | No concerns | Major concerns | No concerns | No concerns | Moderate | ["Within-study bias", "Imprecision"] |
| **Bupropion:Topiramate** | 0 | Some concerns | Low risk | No concerns | Major concerns | No concerns | No concerns | Moderate | ["Within-study bias", "Imprecision"] |
| **Fluvoxamine:Nalmefene** | 0 | Some concerns | Some concerns | No concerns | Major concerns | No concerns | No concerns | Low | ["Within-study bias", "Reporting bias", "Imprecision"] |
| **Fluvoxamine:Naloxone** | 0 | Some concerns | Low risk | No concerns | Major concerns | No concerns | No concerns | Moderate | ["Within-study bias", "Imprecision"] |
| **Fluvoxamine:Naltrexone** | 0 | Major concerns | Low risk | No concerns | Major concerns | No concerns | No concerns | Low | ["Within-study bias", "Imprecision"] |
| **Fluvoxamine:Olanzapine** | 0 | Major concerns | Some concerns | No concerns | Major concerns | No concerns | No concerns | Very low | ["Within-study bias", "Reporting bias", "Imprecision"] |
| **Fluvoxamine:Paroxetine** | 0 | Some concerns | Low risk | No concerns | Major concerns | No concerns | No concerns | Moderate | ["Within-study bias", "Imprecision"] |
| **Fluvoxamine:Silymarin** | 0 | Some concerns | Low risk | No concerns | Major concerns | No concerns | No concerns | Moderate | ["Within-study bias", "Imprecision"] |
| **Fluvoxamine:Topiramate** | 0 | Some concerns | Low risk | No concerns | Major concerns | No concerns | No concerns | Moderate | ["Within-study bias", "Imprecision"] |
| **Nalmefene:Naloxone** | 0 | Some concerns | Some concerns | No concerns | No concerns | Major concerns | No concerns | Low | ["Within-study bias", "Reporting bias", "Heterogeneity"] |
| **Nalmefene:Naltrexone** | 0 | Some concerns | Low risk | No concerns | Major concerns | No concerns | No concerns | Moderate | ["Within-study bias", "Imprecision"] |
| **Nalmefene:Olanzapine** | 0 | Some concerns | Low risk | No concerns | Some concerns | Some concerns | No concerns | Moderate | ["Within-study bias", "Imprecision", "Heterogeneity"] |
| **Nalmefene:Paroxetine** | 0 | Some concerns | Some concerns | No concerns | Some concerns | Some concerns | No concerns | Moderate | ["Within-study bias", "Reporting bias", "Imprecision", "Heterogeneity"] |
| **Nalmefene:Silymarin** | 0 | Some concerns | Some concerns | No concerns | Some concerns | Some concerns | No concerns | Moderate | ["Within-study bias", "Reporting bias", "Imprecision", "Heterogeneity"] |
| **Nalmefene:Topiramate** | 0 | Some concerns | Some concerns | No concerns | Major concerns | No concerns | No concerns | Low | ["Within-study bias", "Reporting bias", "Imprecision"] |
| **Naloxone:Naltrexone** | 0 | Some concerns | Low risk | No concerns | Major concerns | No concerns | No concerns | Moderate | ["Within-study bias", "Imprecision"] |
| **Naloxone:Olanzapine** | 0 | Some concerns | Some concerns | No concerns | Major concerns | No concerns | No concerns | Low | ["Within-study bias", "Reporting bias", "Imprecision"] |
| **Naloxone:Paroxetine** | 0 | Some concerns | Low risk | No concerns | Major concerns | No concerns | No concerns | Moderate | ["Within-study bias", "Imprecision"] |
| **Naloxone:Silymarin** | 0 | Some concerns | Low risk | No concerns | Major concerns | No concerns | No concerns | Moderate | ["Within-study bias", "Imprecision"] |
| **Naloxone:Topiramate** | 0 | Some concerns | Low risk | No concerns | Major concerns | No concerns | No concerns | Moderate | ["Within-study bias", "Imprecision"] |
| **Naltrexone:Olanzapine** | 0 | Some concerns | Low risk | No concerns | Major concerns | No concerns | No concerns | Moderate | ["Within-study bias", "Imprecision"] |
| **Naltrexone:Paroxetine** | 0 | Some concerns | Low risk | No concerns | Major concerns | No concerns | No concerns | Moderate | ["Within-study bias", "Imprecision"] |
| **Naltrexone:Silymarin** | 0 | Some concerns | Low risk | No concerns | Major concerns | No concerns | No concerns | Moderate | ["Within-study bias", "Imprecision"] |
| **Naltrexone:Topiramate** | 0 | Some concerns | Low risk | No concerns | Major concerns | No concerns | No concerns | Moderate | ["Within-study bias", "Imprecision"] |
| **Olanzapine:Paroxetine** | 0 | Some concerns | Some concerns | No concerns | Major concerns | No concerns | No concerns | Low | ["Within-study bias", "Reporting bias", "Imprecision"] |
| **Olanzapine:Silymarin** | 0 | Some concerns | Some concerns | No concerns | Major concerns | No concerns | No concerns | Low | ["Within-study bias", "Reporting bias", "Imprecision"] |
| **Olanzapine:Topiramate** | 0 | Some concerns | Some concerns | No concerns | Major concerns | No concerns | No concerns | Low | ["Within-study bias", "Reporting bias", "Imprecision"] |
| **Paroxetine:Silymarin** | 0 | Some concerns | Low risk | No concerns | Major concerns | No concerns | No concerns | Moderate | ["Within-study bias", "Imprecision"] |
| **Paroxetine:Topiramate** | 0 | Some concerns | Low risk | No concerns | Major concerns | No concerns | No concerns | Moderate | ["Within-study bias", "Imprecision"] |
| **Silymarin:Topiramate** | 0 | Some concerns | Low risk | No concerns | Major concerns | No concerns | No concerns | Moderate | ["Within-study bias", "Imprecision"] |

### §S10 CINeMA full reports

#### Gambling symptoms severity

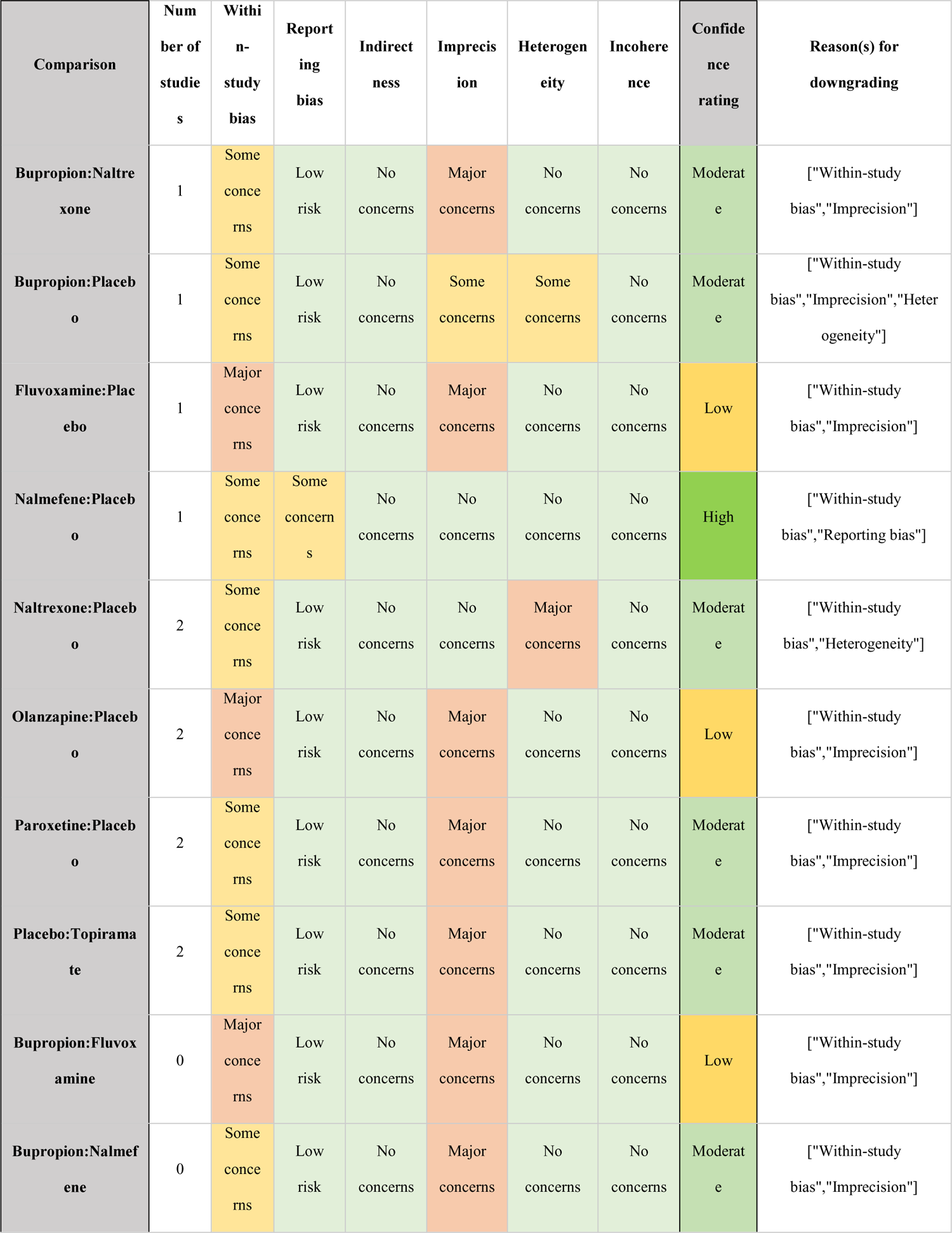
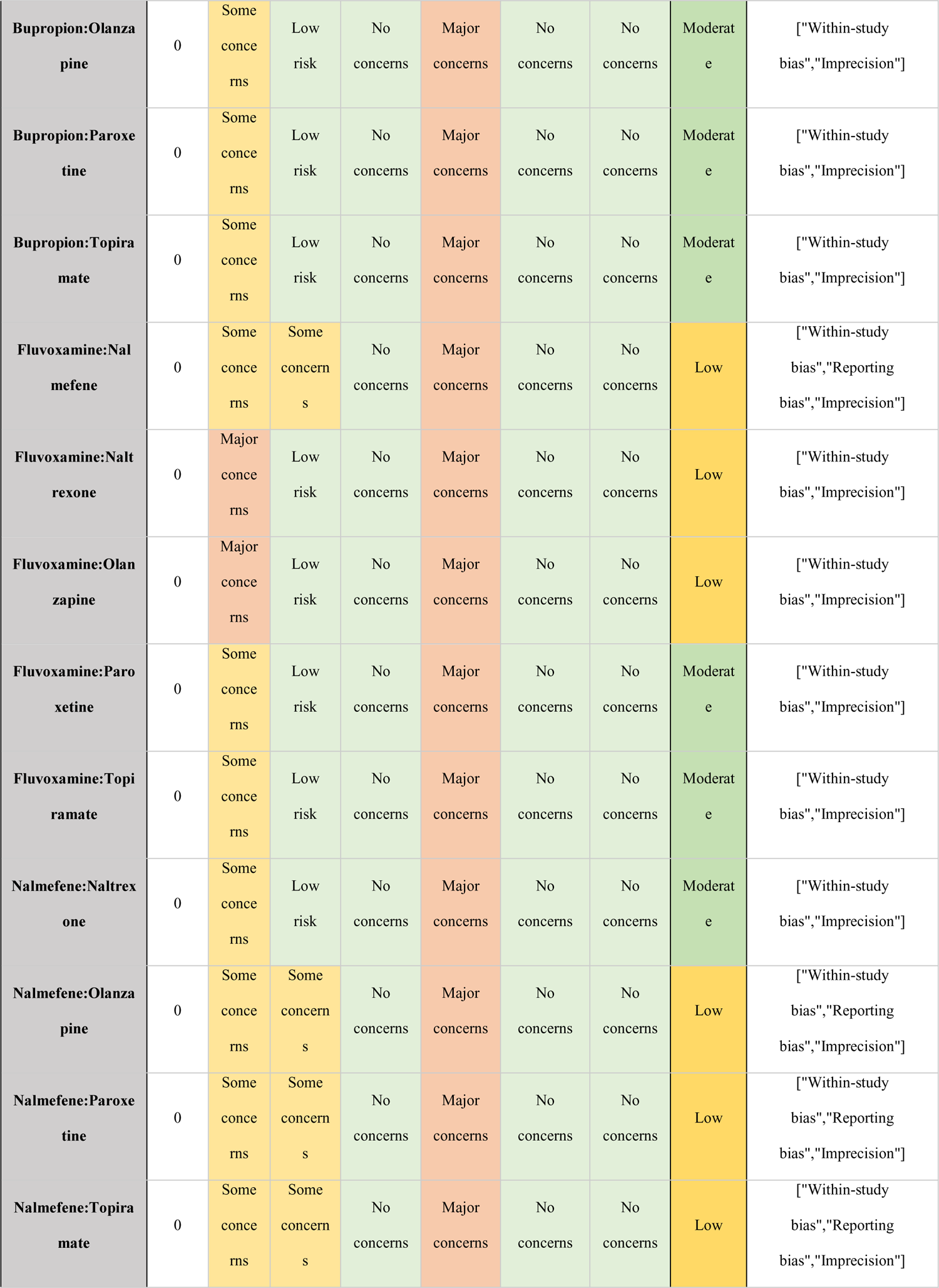
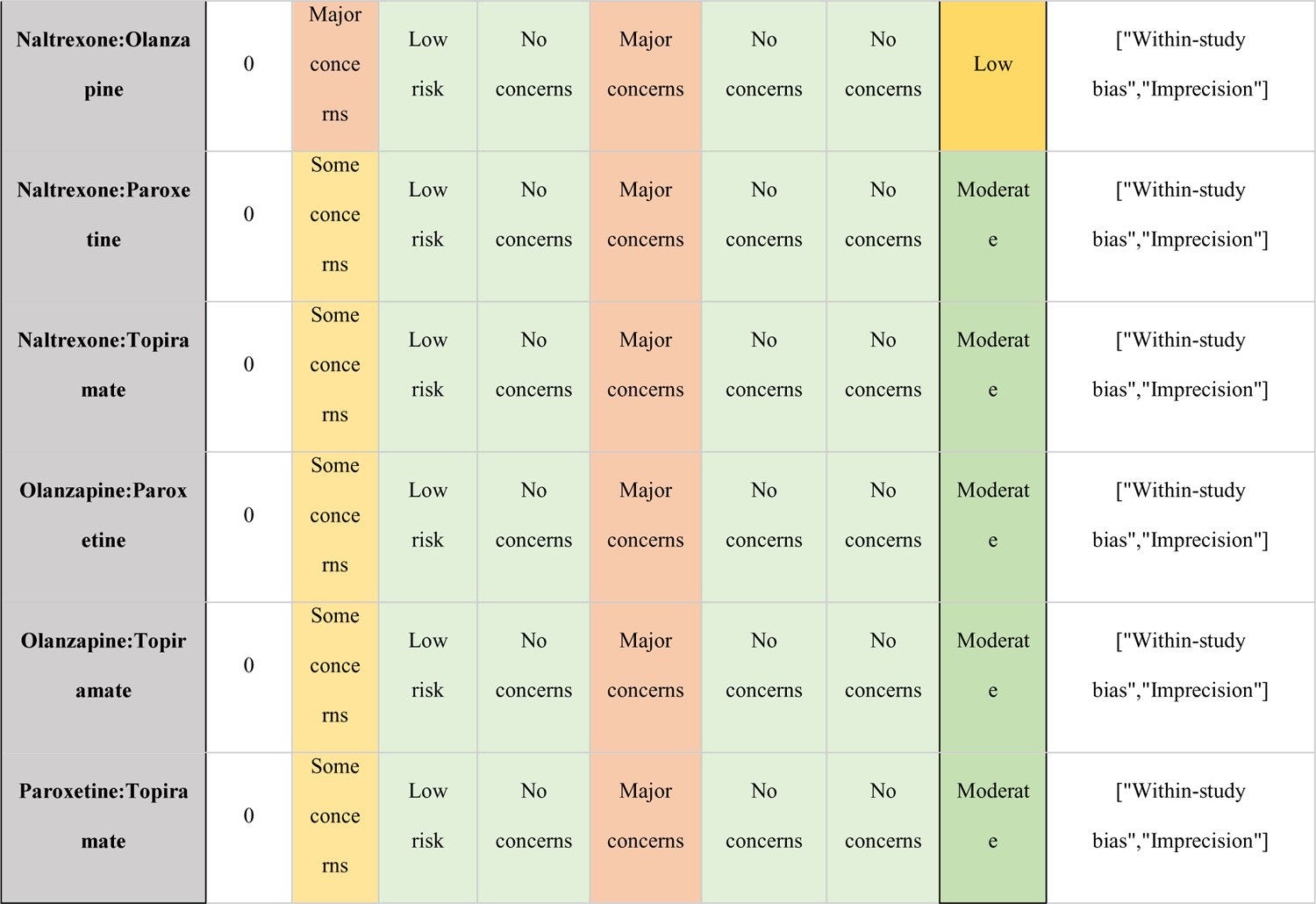

*Tolerability*
| Comparison | Number of studies | Within-study bias | Reporting bias | Indirectness | Imprecision | Heterogeneity | Incoherence | Confidence rating | Reason(s) for downgrading |
| --- | --- | --- | --- | --- | --- | --- | --- | --- | --- |
| Bupropion:Naltrexone | 1 | Some concerns | Low risk | No concerns | Major concerns | No concerns | No concerns | Moderate | ["Within-study bias", "Imprecision"] |
| Bupropion:Placebo | 1 | Some concerns | Low risk | No concerns | Some concerns | Some concerns | No concerns | Moderate | ["Within-study bias", "Imprecision", "Heterogeneity"] |
| Fluvoxamine:Placebo | 1 | Major concerns | Low risk | No concerns | Major concerns | No concerns | No concerns | Low | ["Within-study bias", "Imprecision"] |
| Nalmefene:Placebo | 1 | Some concerns | Some concerns | No concerns | No concerns | No concerns | No concerns | High | ["Within-study bias", "Reporting bias"] |
| Naltrexone:Placebo | 2 | Some concerns | Low risk | No concerns | No concerns | Major concerns | No concerns | Moderate | ["Within-study bias", "Heterogeneity"] |
| Olanzapine:Placebo | 2 | Major concerns | Low risk | No concerns | Major concerns | No concerns | No concerns | Low | ["Within-study bias", "Imprecision"] |
| Paroxetine:Placebo | 2 | Some concerns | Low risk | No concerns | Major concerns | No concerns | No concerns | Moderate | ["Within-study bias", "Imprecision"] |
| Placebo:Topiramate | 2 | Some concerns | Low risk | No concerns | Major concerns | No concerns | No concerns | Moderate | ["Within-study bias", "Imprecision"] |
| Bupropion:Fluvoxamine | 0 | Major concerns | Low risk | No concerns | Major concerns | No concerns | No concerns | Low | ["Within-study bias", "Imprecision"] |
| Bupropion:Nalmefene | 0 | Some concerns | Low risk | No concerns | Major concerns | No concerns | No concerns | Moderate | ["Within-study bias", "Imprecision"] |
| **Bupropion:Olanzapine** | 0 | Some concerns | Low risk | No concerns | Major concerns | No concerns | No concerns | Moderate | ["Within-study bias", "Imprecision"] |
| **Bupropion:Paroxetine** | 0 | Some concerns | Low risk | No concerns | Major concerns | No concerns | No concerns | Moderate | ["Within-study bias", "Imprecision"] |
| **Bupropion:Topiramate** | 0 | Some concerns | Low risk | No concerns | Major concerns | No concerns | No concerns | Moderate | ["Within-study bias", "Imprecision"] |
| **Fluvoxamine:Nalmefene** | 0 | Some concerns | Some concerns | No concerns | Major concerns | No concerns | No concerns | Low | ["Within-study bias", "Reporting bias", "Imprecision"] |
| **Fluvoxamine:Naltrexone** | 0 | Major concerns | Low risk | No concerns | Major concerns | No concerns | No concerns | Low | ["Within-study bias", "Imprecision"] |
| **Fluvoxamine:Olanzapine** | 0 | Major concerns | Low risk | No concerns | Major concerns | No concerns | No concerns | Low | ["Within-study bias", "Imprecision"] |
| **Fluvoxamine:Paroxetine** | 0 | Some concerns | Low risk | No concerns | Major concerns | No concerns | No concerns | Moderate | ["Within-study bias", "Imprecision"] |
| **Fluvoxamine:Topiramate** | 0 | Some concerns | Low risk | No concerns | Major concerns | No concerns | No concerns | Moderate | ["Within-study bias", "Imprecision"] |
| **Nalmefene:Naltrexone** | 0 | Some concerns | Low risk | No concerns | Major concerns | No concerns | No concerns | Moderate | ["Within-study bias", "Imprecision"] |
| **Nalmefene:Olanzapine** | 0 | Some concerns | Some concerns | No concerns | Major concerns | No concerns | No concerns | Low | ["Within-study bias", "Reporting bias", "Imprecision"] |
| **Nalmefene:Paroxetine** | 0 | Some concerns | Some concerns | No concerns | Major concerns | No concerns | No concerns | Low | ["Within-study bias", "Reporting bias", "Imprecision"] |
| **Nalmefene:Topiramate** | 0 | Some concerns | Some concerns | No concerns | Major concerns | No concerns | No concerns | Low | ["Within-study bias", "Reporting bias", "Imprecision"] |
| Naltrexone:Olanzapine | 0 | Major concerns | Low risk | No concerns | Major concerns | No concerns | No concerns | Low | ["Within-study bias", "Imprecision"] |
| Naltrexone:Paroxetine | 0 | Some concerns | Low risk | No concerns | Major concerns | No concerns | No concerns | Moderate | ["Within-study bias", "Imprecision"] |
| Naltrexone:Topiramate | 0 | Some concerns | Low risk | No concerns | Major concerns | No concerns | No concerns | Moderate | ["Within-study bias", "Imprecision"] |
| Olanzapine:Paroxetine | 0 | Some concerns | Low risk | No concerns | Major concerns | No concerns | No concerns | Moderate | ["Within-study bias", "Imprecision"] |
| Olanzapine:Topiramate | 0 | Some concerns | Low risk | No concerns | Major concerns | No concerns | No concerns | Moderate | ["Within-study bias", "Imprecision"] |
| Paroxetine:Topiramate | 0 | Some concerns | Low risk | No concerns | Major concerns | No concerns | No concerns | Moderate | ["Within-study bias", "Imprecision"] |

#### Tolerability

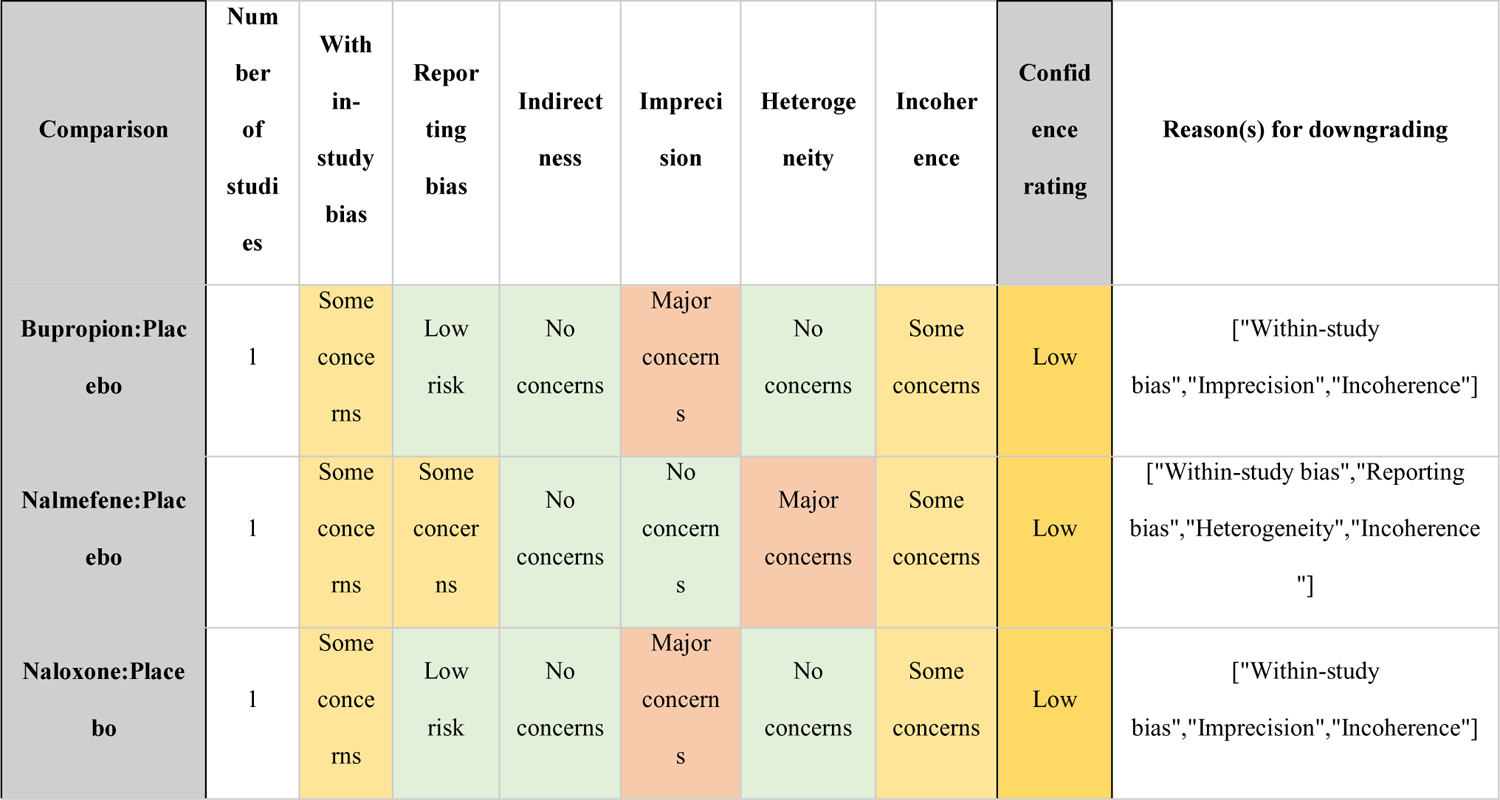
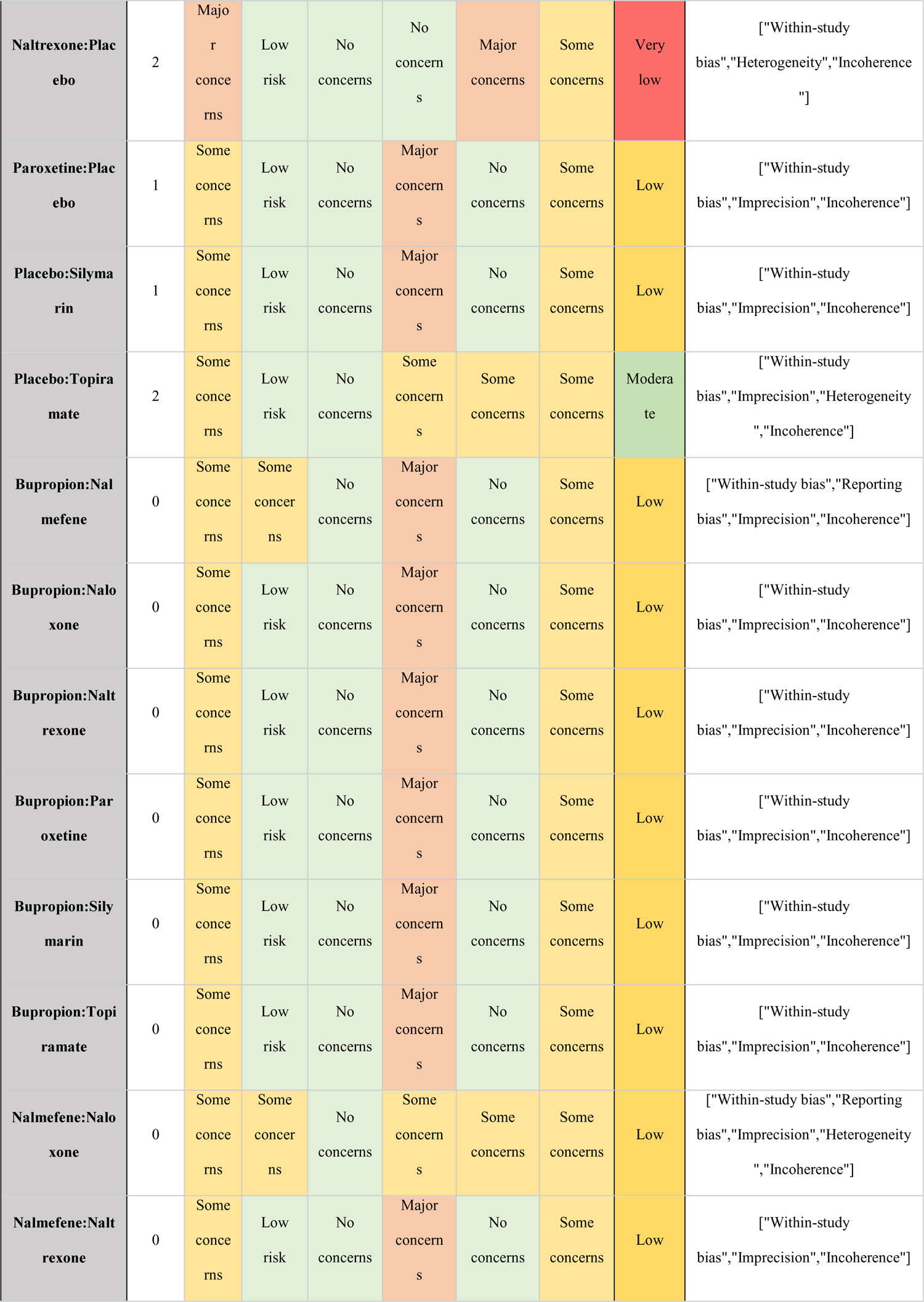
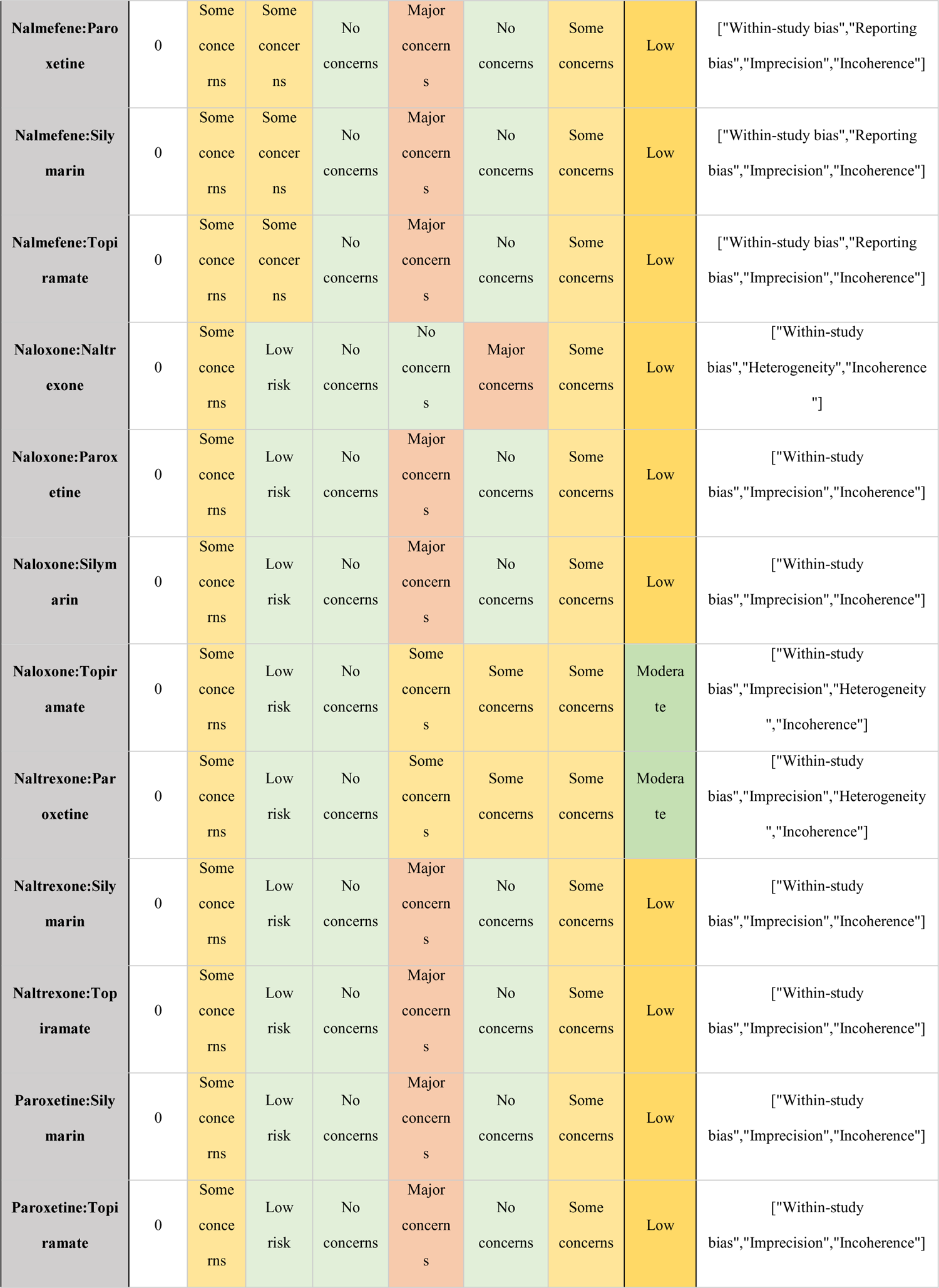
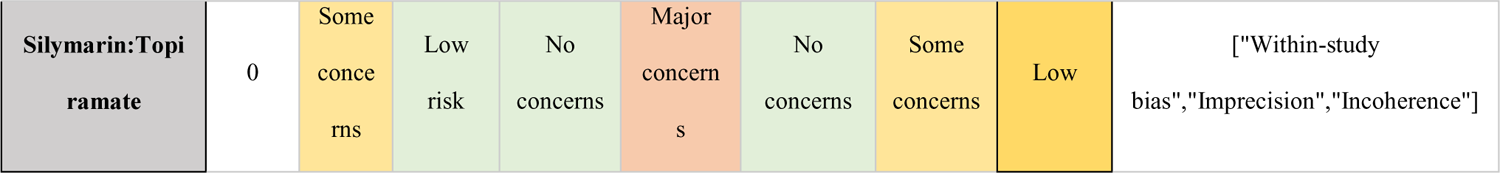

Quality of Life
| Comparison | Number of studies | With in-study bias | Reporting bias | Indirectness | Imprecision | Heterogeneity | Incoherence | Confidence rating | Reason(s) for downgrading |
| --- | --- | --- | --- | --- | --- | --- | --- | --- | --- |
| Bupropion:Placebo | 1 | Some concerns | Low risk | No concerns | Major concerns | No concerns | Some concerns | Low | ["Within-study bias", "Imprecision", "Incoherence"] |
| Nalmefene:Placebo | 1 | Some concerns | Some concerns | No concerns | No concerns | Major concerns | Some concerns | Low | ["Within-study bias", "Reporting bias", "Heterogeneity", "Incoherence"] |
| Naloxone:Placebo | 1 | Some concerns | Low risk | No concerns | Major concerns | No concerns | Some concerns | Low | ["Within-study bias", "Imprecision", "Incoherence"] |
| **Naltrexone:Placebo** | 2 | Major concerns | Low risk | No concerns | No concerns | Major concerns | Some concerns | Very low | ["Within-study bias", "Heterogeneity", "Incoherence"] |
| **Paroxetine:Placebo** | 1 | Some concerns | Low risk | No concerns | Major concerns | No concerns | Some concerns | Low | ["Within-study bias", "Imprecision", "Incoherence"] |
| **Placebo:Silymarin** | 1 | Some concerns | Low risk | No concerns | Major concerns | No concerns | Some concerns | Low | ["Within-study bias", "Imprecision", "Incoherence"] |
| **Placebo:Topiramate** | 2 | Some concerns | Low risk | No concerns | Some concerns | Some concerns | Some concerns | Moderate | ["Within-study bias", "Imprecision", "Heterogeneity", "Incoherence"] |
| **Bupropion:Nalmefene** | 0 | Some concerns | Some concerns | No concerns | Major concerns | No concerns | Some concerns | Low | ["Within-study bias", "Reporting bias", "Imprecision", "Incoherence"] |
| **Bupropion:Naloxone** | 0 | Some concerns | Low risk | No concerns | Major concerns | No concerns | Some concerns | Low | ["Within-study bias", "Imprecision", "Incoherence"] |
| **Bupropion:Naltrexone** | 0 | Some concerns | Low risk | No concerns | Major concerns | No concerns | Some concerns | Low | ["Within-study bias", "Imprecision", "Incoherence"] |
| **Bupropion:Paroxetine** | 0 | Some concerns | Low risk | No concerns | Major concerns | No concerns | Some concerns | Low | ["Within-study bias", "Imprecision", "Incoherence"] |
| **Bupropion:Silymarin** | 0 | Some concerns | Low risk | No concerns | Major concerns | No concerns | Some concerns | Low | ["Within-study bias", "Imprecision", "Incoherence"] |
| **Bupropion:Topiramate** | 0 | Some concerns | Low risk | No concerns | Major concerns | No concerns | Some concerns | Low | ["Within-study bias", "Imprecision", "Incoherence"] |
| **Nalmefene:Naloxone** | 0 | Some concerns | Some concerns | No concerns | Some concerns | Some concerns | Some concerns | Low | ["Within-study bias", "Reporting bias", "Imprecision", "Heterogeneity", "Incoherence"] |
| **Nalmefene:Naltrexone** | 0 | Some concerns | Low risk | No concerns | Major concerns | No concerns | Some concerns | Low | ["Within-study bias", "Imprecision", "Incoherence"] |
| **Nalmefene:Paroxetine** | 0 | Some concerns | Some concerns | No concerns | Major concerns | No concerns | Some concerns | Low | ["Within-study bias", "Reporting bias", "Imprecision", "Incoherence"] |
| **Nalmefene:Silymarin** | 0 | Some concerns | Some concerns | No concerns | Major concerns | No concerns | Some concerns | Low | ["Within-study bias", "Reporting bias", "Imprecision", "Incoherence"] |
| **Nalmefene:Topiramate** | 0 | Some concerns | Some concerns | No concerns | Major concerns | No concerns | Some concerns | Low | ["Within-study bias", "Reporting bias", "Imprecision", "Incoherence"] |
| **Naloxone:Naltrexone** | 0 | Some concerns | Low risk | No concerns | No concerns | Major concerns | Some concerns | Low | ["Within-study bias", "Heterogeneity", "Incoherence"] |
| **Naloxone:Paroxetine** | 0 | Some concerns | Low risk | No concerns | Major concerns | No concerns | Some concerns | Low | ["Within-study bias", "Imprecision", "Incoherence"] |
| **Naloxone:Silymarin** | 0 | Some concerns | Low risk | No concerns | Major concerns | No concerns | Some concerns | Low | ["Within-study bias", "Imprecision", "Incoherence"] |
| **Naloxone:Topiramate** | 0 | Some concerns | Low risk | No concerns | Some concerns | Some concerns | Some concerns | Moderate | ["Within-study bias", "Imprecision", "Heterogeneity", "Incoherence"] |
| **Naltrexone:Paroxetine** | 0 | Some concerns | Low risk | No concerns | Some concerns | Some concerns | Some concerns | Moderate | ["Within-study bias", "Imprecision", "Heterogeneity", "Incoherence"] |
| **Naltrexone:Silymarin** | 0 | Some concerns | Low risk | No concerns | Major concerns | No concerns | Some concerns | Low | ["Within-study bias", "Imprecision", "Incoherence"] |
| **Naltrexone:Topiramate** | 0 | Some concerns | Low risk | No concerns | Major concerns | No concerns | Some concerns | Low | ["Within-study bias", "Imprecision", "Incoherence"] |
| **Paroxetine:Silymarin** | 0 | Some concerns | Low risk | No concerns | Major concerns | No concerns | Some concerns | Low | ["Within-study bias", "Imprecision", "Incoherence"] |
| **Paroxetine:Topiramate** | 0 | Some concerns | Low risk | No concerns | Major concerns | No concerns | Some concerns | Low | ["Within-study bias", "Imprecision", "Incoherence"] |
| Silymarin:Topiramate | 0 | Some concerns | Low risk | No concerns | Major concerns | No concerns | Some concerns | Low | ["Within-study bias", "Imprecision", "Incoherence"] |

#### Quality of Life

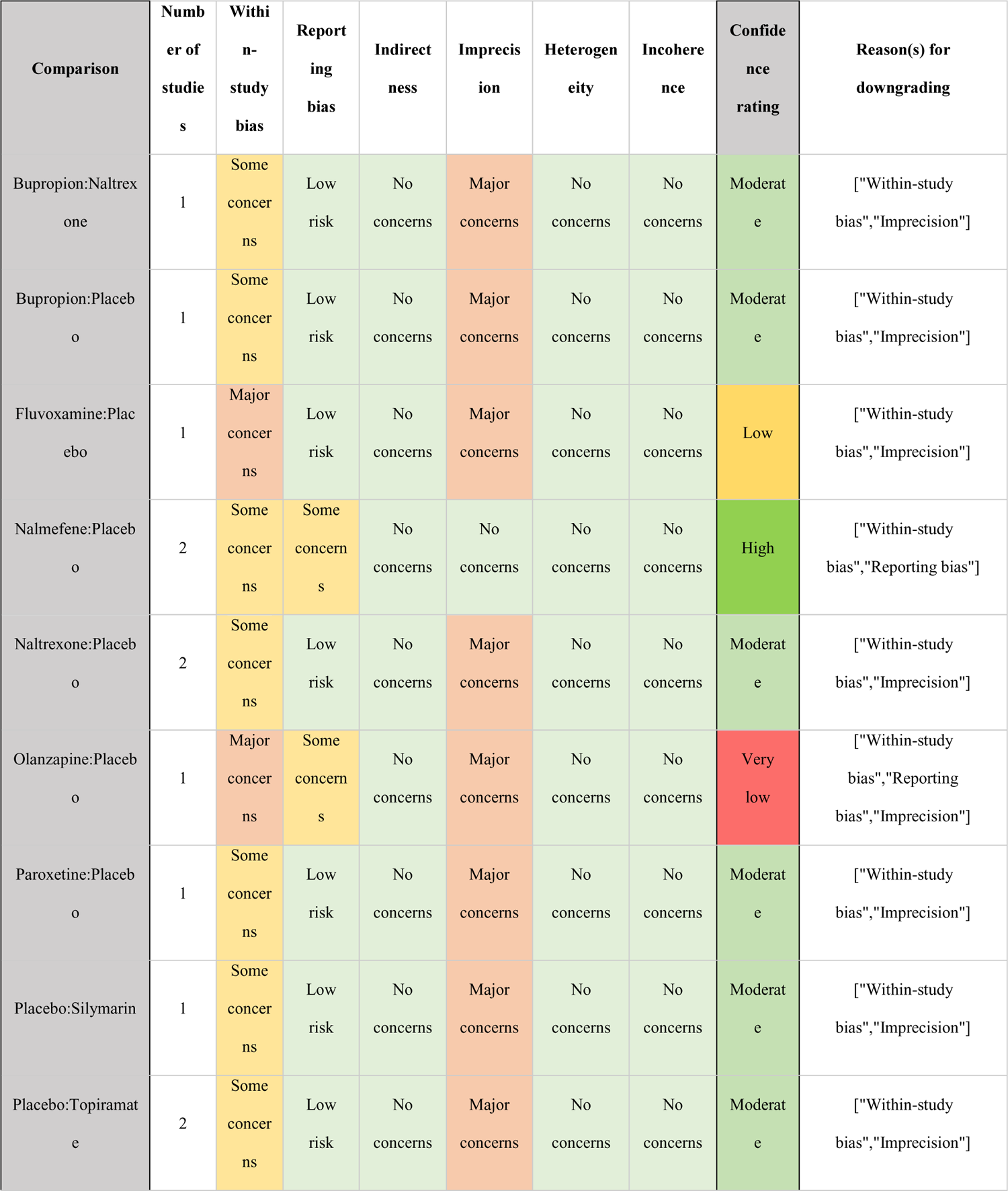
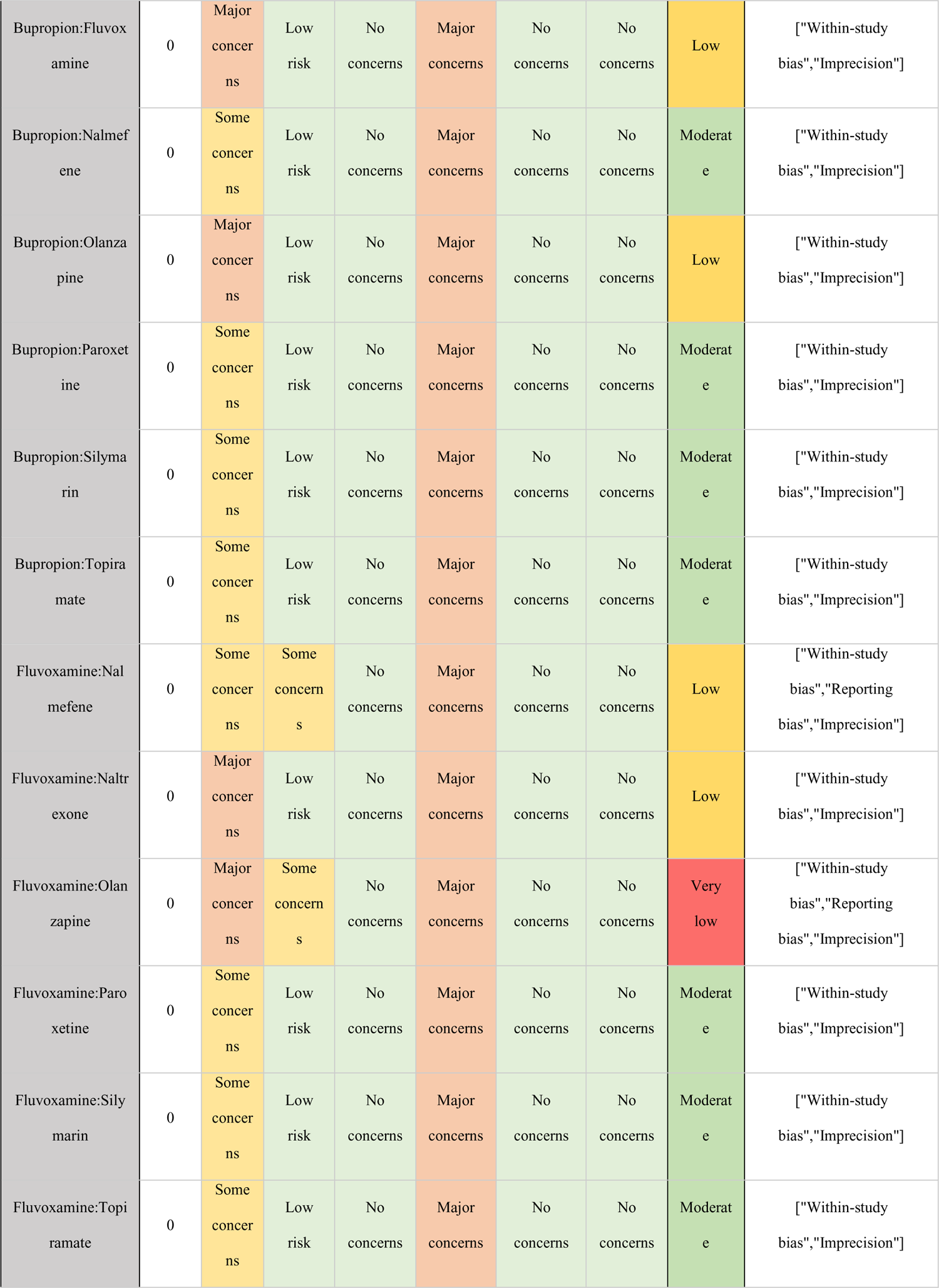
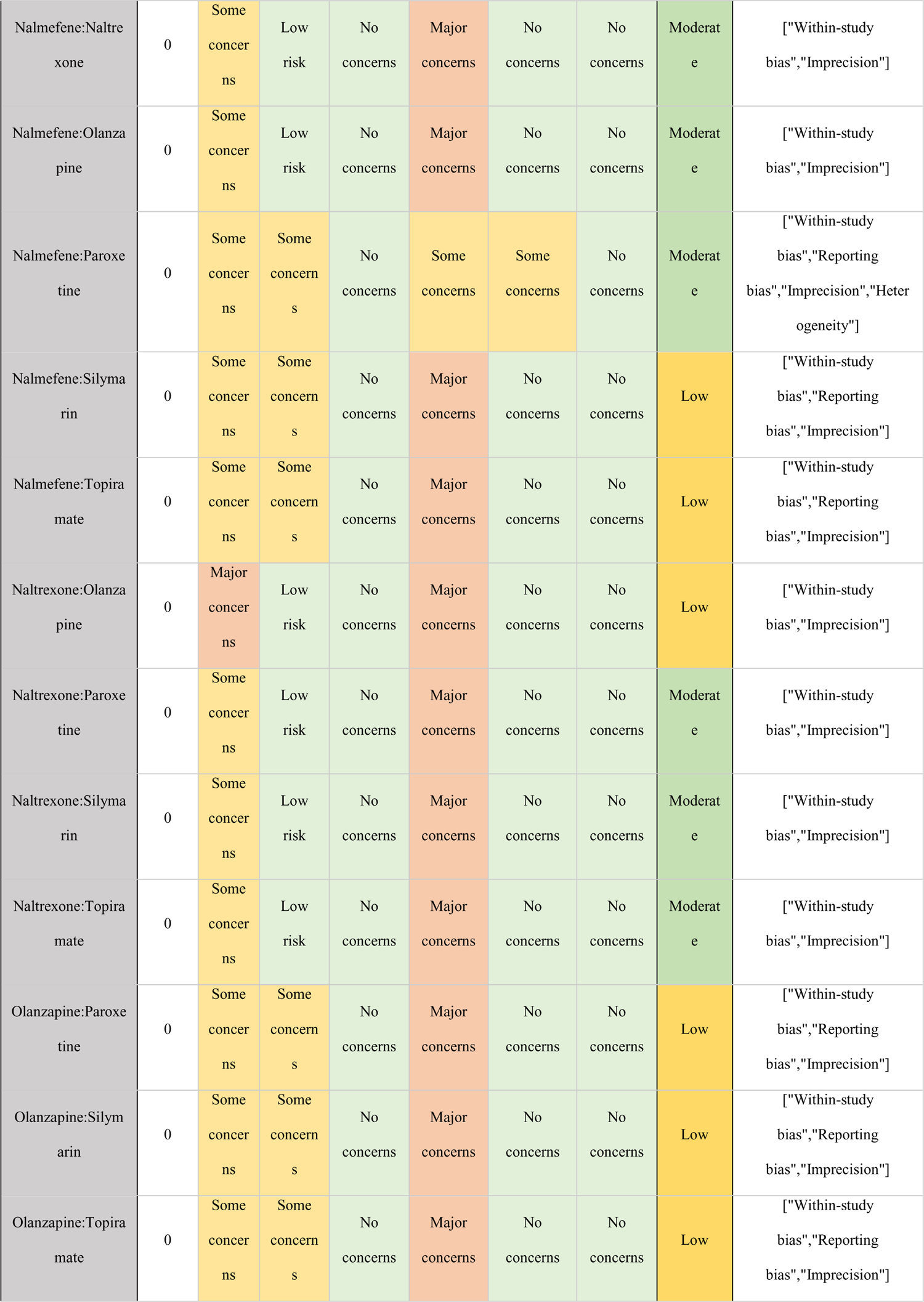
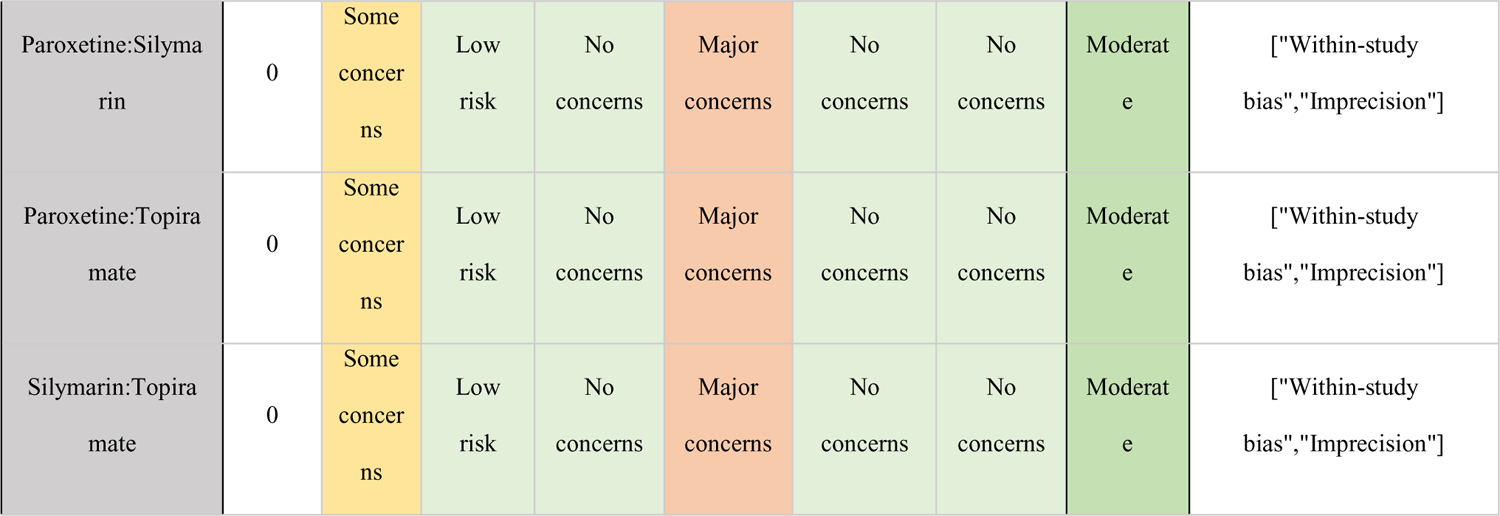

Gambling Severity – PGYBOCS
| Comparison | Number of studies | Within-study bias | Reporting bias | Indirectness | Imprecision | Heterogeneity | Incoherence | Confidence rating | Reason(s) for downgrading |
| --- | --- | --- | --- | --- | --- | --- | --- | --- | --- |
| Bupropion:Naltrexone | 1 | Some concerns | Low risk | No concerns | Major concerns | No concerns | No concerns | Moderate | ["Within-study bias", "Imprecision"] |
| Bupropion:Placebo | 1 | Some concerns | Low risk | No concerns | Major concerns | No concerns | No concerns | Moderate | ["Within-study bias", "Imprecision"] |
| Fluvoxamine:Placebo | 1 | Major concerns | Low risk | No concerns | Major concerns | No concerns | No concerns | Low | ["Within-study bias", "Imprecision"] |
| Nalmefene:Placebo | 2 | Some concerns | Some concerns | No concerns | No concerns | No concerns | No concerns | High | ["Within-study bias", "Reporting bias"] |
| Naltrexone:Placebo | 2 | Some concerns | Low risk | No concerns | Major concerns | No concerns | No concerns | Moderate | ["Within-study bias", "Imprecision"] |
| Olanzapine:Placebo | 1 | Major concerns | Some concerns | No concerns | Major concerns | No concerns | No concerns | Very low | ["Within-study bias", "Reporting bias", "Imprecision"] |
| Paroxetine:Placebo | 1 | Some concerns | Low risk | No concerns | Major concerns | No concerns | No concerns | Moderate | ["Within-study bias", "Imprecision"] |
| Placebo:Silymarin | 1 | Some concerns | Low risk | No concerns | Major concerns | No concerns | No concerns | Moderate | ["Within-study bias", "Imprecision"] |
| Placebo:Topiramate | 2 | Some concerns | Low risk | No concerns | Major concerns | No concerns | No concerns | Moderate | ["Within-study bias", "Imprecision"] |
| Bupropion:Fluvoxamine | 0 | Major concerns | Low risk | No concerns | Major concerns | No concerns | No concerns | Low | ["Within-study bias", "Imprecision"] |
| Bupropion:Nalmefene | 0 | Some concerns | Low risk | No concerns | Major concerns | No concerns | No concerns | Moderate | ["Within-study bias", "Imprecision"] |
| Bupropion:Olanzapine | 0 | Major concerns | Low risk | No concerns | Major concerns | No concerns | No concerns | Low | ["Within-study bias", "Imprecision"] |
| Bupropion:Paroxetine | 0 | Some concerns | Low risk | No concerns | Major concerns | No concerns | No concerns | Moderate | ["Within-study bias", "Imprecision"] |
| Bupropion:Silymarin | 0 | Some concerns | Low risk | No concerns | Major concerns | No concerns | No concerns | Moderate | ["Within-study bias", "Imprecision"] |
| Bupropion:Topiramate | 0 | Some concerns | Low risk | No concerns | Major concerns | No concerns | No concerns | Moderate | ["Within-study bias", "Imprecision"] |
| Fluvoxamine:Nalmefene | 0 | Some concerns | Some concerns | No concerns | Major concerns | No concerns | No concerns | Low | ["Within-study bias", "Reporting bias", "Imprecision"] |
| Fluvoxamine:Naltrexone | 0 | Major concerns | Low risk | No concerns | Major concerns | No concerns | No concerns | Low | ["Within-study bias", "Imprecision"] |
| Fluvoxamine:Olanzapine | 0 | Major concerns | Some concerns | No concerns | Major concerns | No concerns | No concerns | Very low | ["Within-study bias", "Reporting bias", "Imprecision"] |
| Fluvoxamine:Paroxetine | 0 | Some concerns | Low risk | No concerns | Major concerns | No concerns | No concerns | Moderate | ["Within-study bias", "Imprecision"] |
| Fluvoxamine:Silymarin | 0 | Some concerns | Low risk | No concerns | Major concerns | No concerns | No concerns | Moderate | ["Within-study bias", "Imprecision"] |
| Fluvoxamine:Topiramate | 0 | Some concerns | Low risk | No concerns | Major concerns | No concerns | No concerns | Moderate | ["Within-study bias", "Imprecision"] |
| Nalmefene:Naltrexone | 0 | Some concerns | Low risk | No concerns | Major concerns | No concerns | No concerns | Moderate | ["Within-study bias", "Imprecision"] |
| Nalmefene:Olanzapine | 0 | Some concerns | Low risk | No concerns | Major concerns | No concerns | No concerns | Moderate | ["Within-study bias", "Imprecision"] |
| Nalmefene:Paroxetine | 0 | Some concerns | Some concerns | No concerns | Some concerns | Some concerns | No concerns | Moderate | ["Within-study bias", "Reporting bias", "Imprecision", "Heterogeneity"] |
| Nalmefene:Silymarin | 0 | Some concerns | Some concerns | No concerns | Major concerns | No concerns | No concerns | Low | ["Within-study bias", "Reporting bias", "Imprecision"] |
| Nalmefene:Topiramate | 0 | Some concerns | Some concerns | No concerns | Major concerns | No concerns | No concerns | Low | ["Within-study bias", "Reporting bias", "Imprecision"] |
| Naltrexone:Olanzapine | 0 | Major concerns | Low risk | No concerns | Major concerns | No concerns | No concerns | Low | ["Within-study bias", "Imprecision"] |
| Naltrexone:Paroxetine | 0 | Some concerns | Low risk | No concerns | Major concerns | No concerns | No concerns | Moderate | ["Within-study bias", "Imprecision"] |
| Naltrexone:Silymarin | 0 | Some concerns | Low risk | No concerns | Major concerns | No concerns | No concerns | Moderate | ["Within-study bias", "Imprecision"] |
| Naltrexone:Topiramate | 0 | Some concerns | Low risk | No concerns | Major concerns | No concerns | No concerns | Moderate | ["Within-study bias", "Imprecision"] |
| Olanzapine:Paroxetine | 0 | Some concerns | Some concerns | No concerns | Major concerns | No concerns | No concerns | Low | ["Within-study bias", "Reporting bias", "Imprecision"] |
| Olanzapine:Silymarin | 0 | Some concerns | Some concerns | No concerns | Major concerns | No concerns | No concerns | Low | ["Within-study bias", "Reporting bias", "Imprecision"] |
| Olanzapine:Topiramate | 0 | Some concerns | Some concerns | No concerns | Major concerns | No concerns | No concerns | Low | ["Within-study bias", "Reporting bias", "Imprecision"] |
| Paroxetine:Silymarin | 0 | Some concerns | Low risk | No concerns | Major concerns | No concerns | No concerns | Moderate | ["Within-study bias", "Imprecision"] |
| Paroxetine:Topiramate | 0 | Some concerns | Low risk | No concerns | Major concerns | No concerns | No concerns | Moderate | ["Within-study bias", "Imprecision"] |
| Silymarin:Topiramate | 0 | Some concerns | Low risk | No concerns | Major concerns | No concerns | No concerns | Moderate | ["Within-study bias", "Imprecision"] |

#### Gambling Severity – PGYBOCS

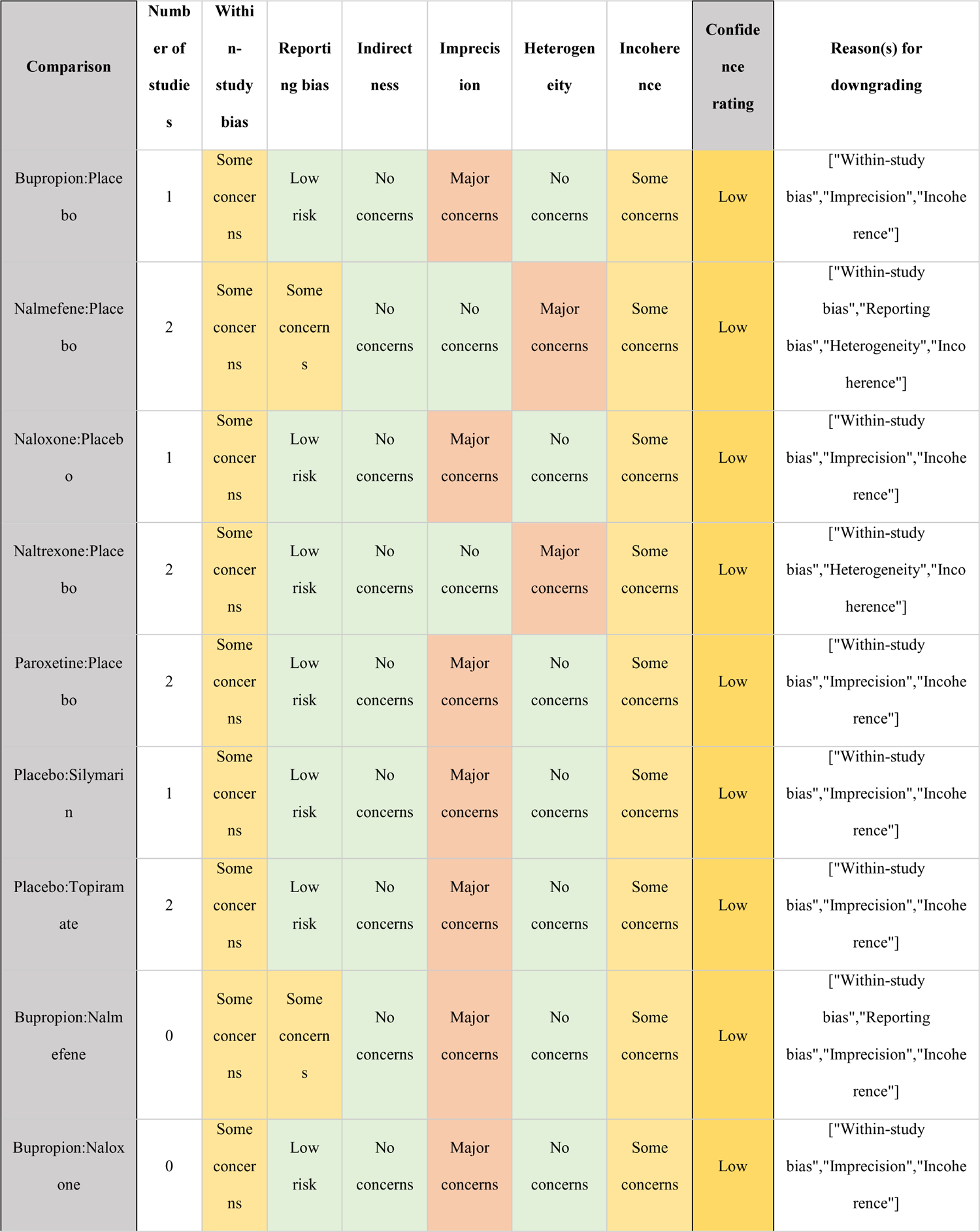
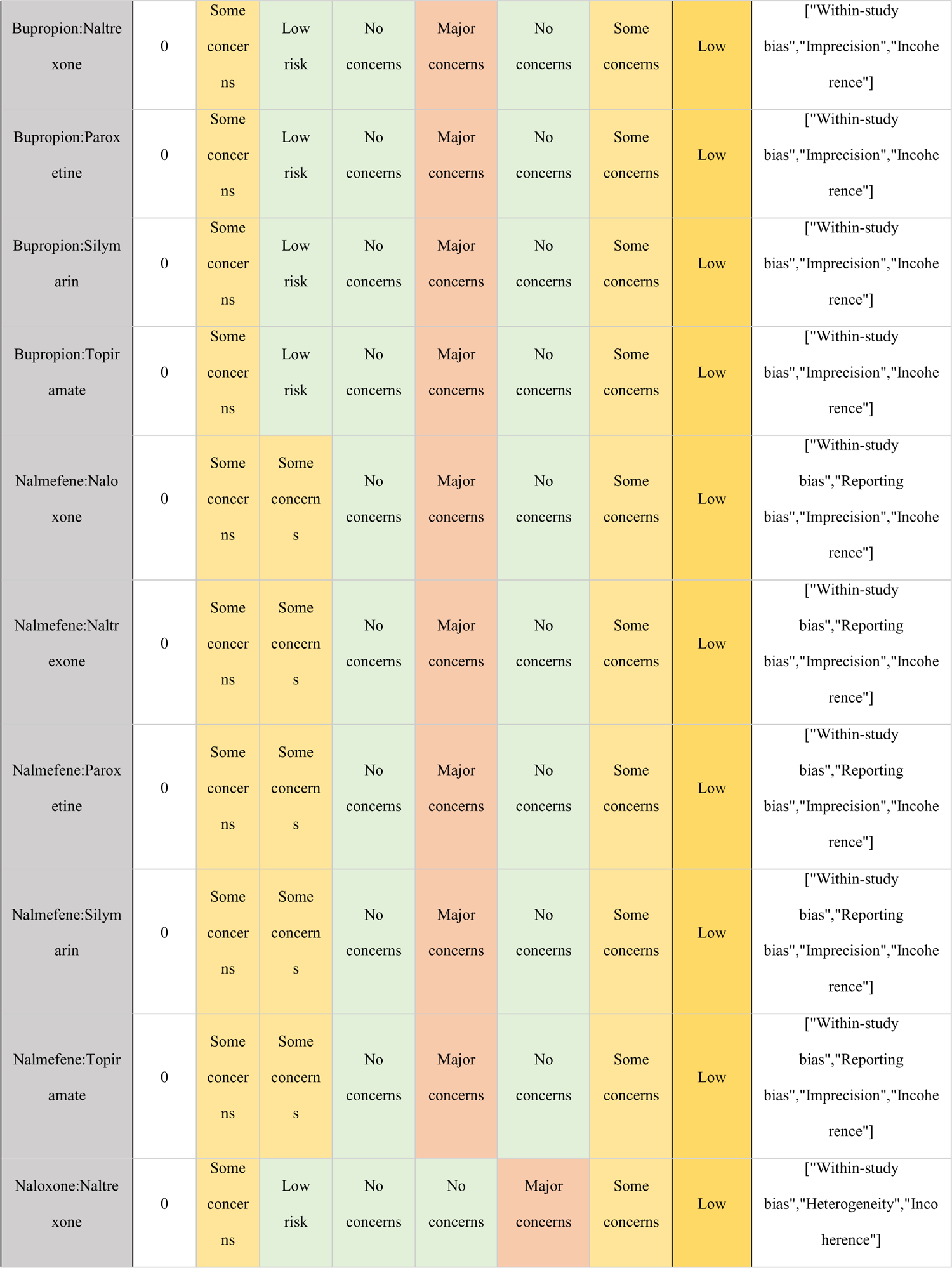
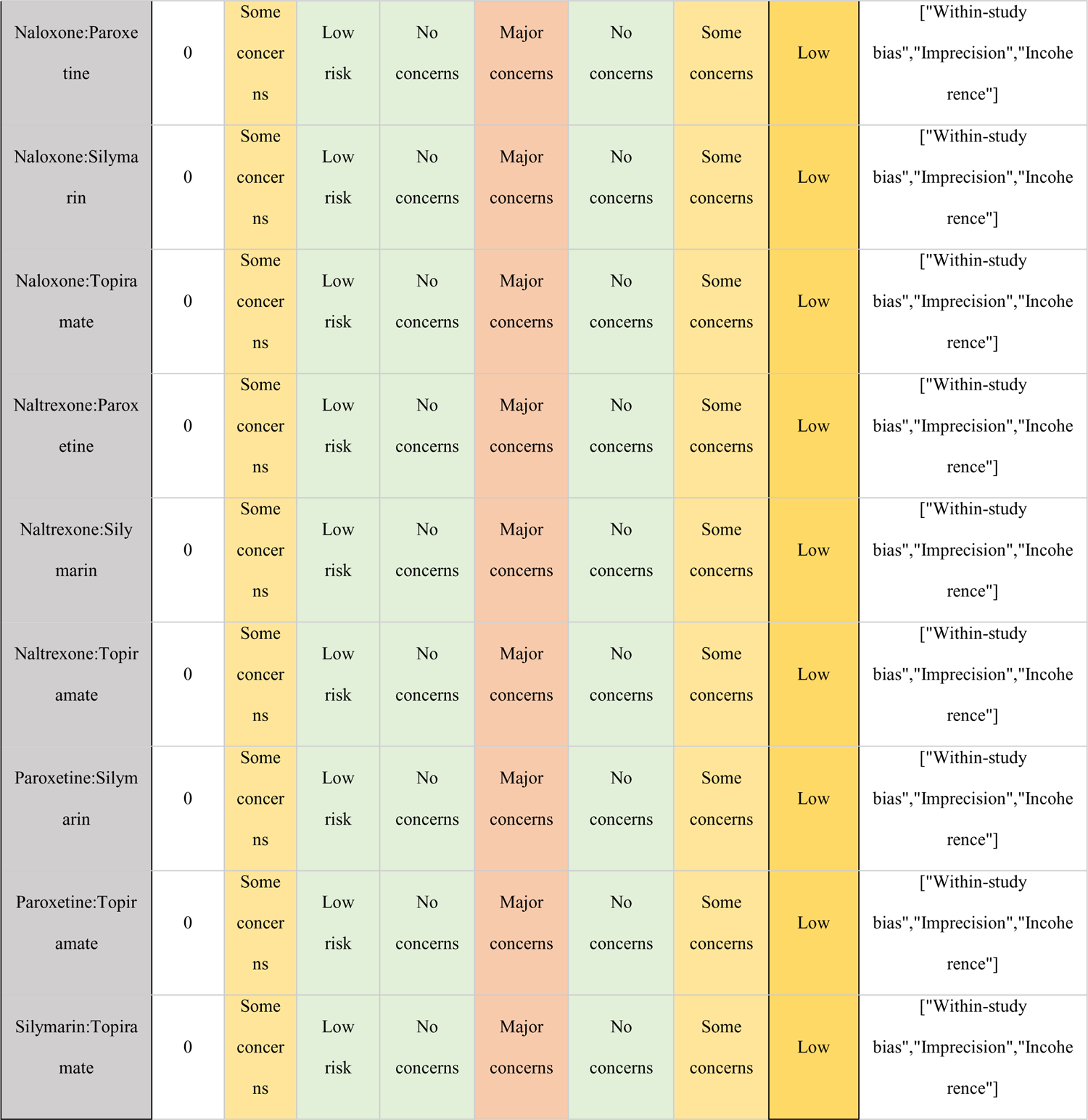

#### Gambling Severity – GSAS

## §S11 Example of STATA code for network meta-analysis

We used STATA®/IC 18.1 (StataCorp LLC, College Station, TX, USA) to perform all analyses. In particular, we have used the commands within the network package to derive forest plot showing the network estimates versus a common comparator, the netleague tables showing the network estimates for each treatment comparison and results for incoherence.

Network meta-analysis (NMA) is a statistical technique used to combine direct and indirect evidence from multiple studies that compare multiple treatments. Here’s a simplified example of how you might perform a network meta-analysis using Stata. Please note that you would need to adapt this code to your specific dataset and research question. ssc install netmeta #Load the required packages for network meta-analysis use “your_dataset.dta”, clear #* Load your dataset (replace “your_dataset.dta” with your actual dataset file) netmeta id study_id treatment outcome effect se_lower se_upper #* Specify the variables for the analysis netmegen id study_id treatment outcome effect se_lower se_upper, graph #* Perform the network meta-analysis netmeprev random #* Run the network meta-analysis model (you might need to adjust the model specification) netmesum #* Summarize the results netmetaforest #* Generate forest plots netmetaleague # * Generate league tables netmetasucra # * Generate surface under the cumulative ranking (SUCRA) plots netmetarankogram #* Generate rankograms netmetasave “results_output.dta” #* Save the results

